# The impact of alternative delivery strategies for novel tuberculosis vaccines in low- and middle-income countries: a modelling study

**DOI:** 10.1101/2022.04.16.22273762

**Authors:** Rebecca A. Clark, Christinah Mukandavire, Allison Portnoy, Chathika K. Weerasuriya, Arminder Deol, Danny Scarponi, Andrew Iskauskas, Roel Bakker, Matthew Quaife, Shelly Malhotra, Nebiat Gebreselassie, Matteo Zignol, Raymond C.W. Hutubessy, Birgitte Giersing, Mark Jit, Rebecca C. Harris, Nicolas A. Menzies, Richard G. White

## Abstract

**Background:** Tuberculosis is a leading infectious cause of death worldwide. Novel vaccines will be required to reach global targets and reverse setbacks from the COVID-19 pandemic. We estimated the impact of novel tuberculosis vaccines in low- and middle-income countries (LMICs) under alternative delivery scenarios.

**Methods:** We calibrated a tuberculosis model to 105 LMICs (93% of global tuberculosis incidence). Vaccine scenarios were implemented as *Basecase*: routine vaccination of 9-year-olds and a one-time vaccination campaign for ages ≥10 with country-specific introduction between 2028–2047 and 5-year scale-up to target coverage; *Accelerated Scale-up*: as *Basecase*, but all countries introducing in 2025 with instant scale-up; and *Routine Only*: as *Basecase*, but routine vaccination only. Vaccines protected against disease for 10-years, with 50% efficacy.

**Findings:** The *Basecase* scenario prevented 44.0 (95% uncertainty range=37.2–51.6) million tuberculosis cases, and 5.0 (4.6–5.4) million tuberculosis deaths before 2050, including 2.2 million in the WHO South-East Asian region. The *Accelerated Scale-up* scenario prevented 65.5 (55.6–76.0) million cases and 7.9 (7.3–8.5) million deaths before 2050. The *Routine Only* scenario prevented 8.8 (7.6–10.1) million cases and 1.1 (0.9–1.2) million deaths before 2050.

**Interpretations:** Novel tuberculosis vaccines could have substantial impact, which will vary depending on delivery strategy. Including a campaign will be crucial for rapid impact. Accelerated introduction, more similar to the pace of COVID-19 vaccines, could increase lives saved by around 60%. Investment is required to support vaccine development, manufacturing, prompt introduction and scale-up.

**Funding:** WHO (2020/985800-0)

## Background

Tuberculosis is one of the leading causes of infectious disease death worldwide, second only to COVID-19 in 2020.^1^ The negative impact of COVID-19 on tuberculosis-related health services, such as delays in diagnosis, treatment, and neonatal vaccination has paused and reversed previous slowly declining trends in mortality.^1,2^

The WHO established the “End TB Strategy” in 2015, with the goal of reducing disease incidence, deaths, and costs worldwide from tuberculosis.^3^ Targets for 2025 and 2035 include reductions in the absolute number of tuberculosis deaths by 75% and 95% and the tuberculosis incidence rate by 50% and 90%, respectively, compared to 2015 levels.^3^ However, the majority of countries are not on track to achieve these targets.^1,4^

The 2035 End TB targets explicitly assumed the introduction of new tools, including a novel tuberculosis vaccine, in 2025.^3^ The WHO has proposed Preferred Product Characteristics for New Tuberculosis Vaccines (WHO PPCs) developed through a highly consultative process, including regulators and policy makers from high burden countries.^5^ While progress has been made, it is unlikely that the 2025 target for novel tuberculosis vaccine introduction will be achieved.

A phase 2b trial of the M72/AS01E candidate vaccine demonstrated an efficacy of 49.7% (95% confidence interval: 2.1–74.2) for preventing disease in adults positive by interferon-gamma release assay (IGRA+) from South Africa, Zambia, and Kenya after three years follow-up,^6^ and a trial of BCG-revaccination appeared efficacious at preventing sustained infection in a cohort of IGRA negative (IGRA-) adolescents in South Africa with an efficacy of 45.4% (6.4–68.1).^7^ Unfortunately, the phase 3 trial of M72/AS01E has not started, and therefore the realistic licensure date, should a positive result be found, may not be for a number of years. Policy changes on BCG-revaccination in adolescents could happen sooner in settings such as South Africa, as the trial is likely to be completed in 2024/2025, but BCG-revaccination has not been tested in infected individuals–a population shown previously to be epidemiologically important for rapid population-level vaccine impact.^8^

This raises critical questions for global and country decision-makers, including: How many lives will be lost if we fail to roll out a novel tuberculosis vaccine by 2025? What is the potential impact if instead vaccines are introduced and rolled out following more traditional timelines, and how do we best prepare for that? And how would these impacts vary by WHO region, income level, and TB burden?

We estimated the potential impact of vaccines meeting the technical specifications of the WHO Preferred Product Characteristics for New Tuberculosis Vaccines in low- and middle-income countries (LMICs), in a range of introduction and scale-up scenarios.

## Methods

### Model Development and Calibration

To estimate the impact of novel tuberculosis vaccines, we developed a compartmental age-stratified dynamic *Mycobacterium tuberculosis* (*Mtb)* transmission model (Figure 1), by adapting features of earlier models.^8,9^ In our model, tuberculosis natural history is represented using eight compartments, allowing for *Mtb* infection along a spectrum from uninfected to active clinical disease.^10,11^ A detailed description of the structure can be found in Supplementary Material section 1, with parameterisation in Supplementary Material section 2.

**Figure 1:**
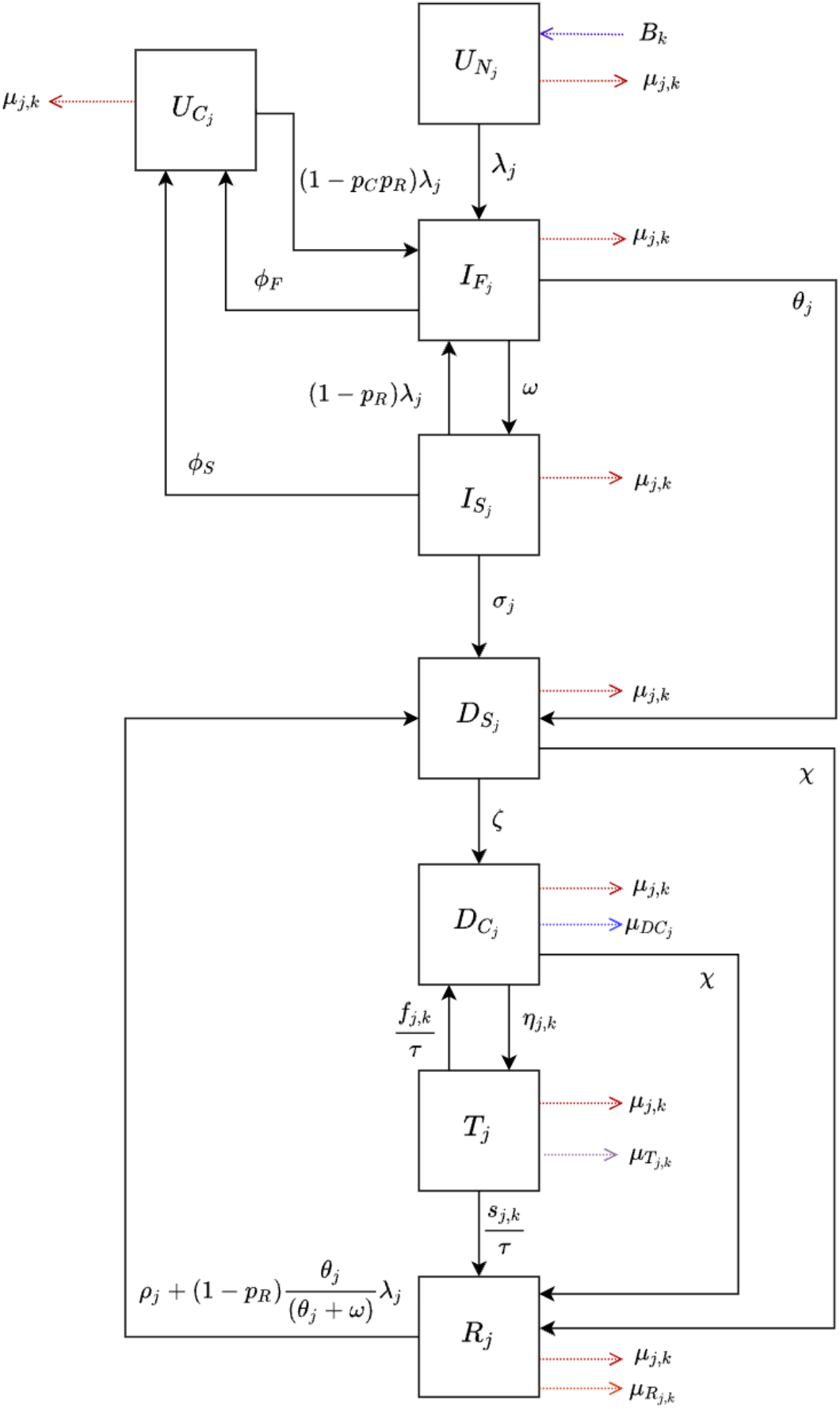
Tuberculosis natural history model structure. DC = Clinical Disease, DS = Subclinical Disease; IF = Infection-Fast, IS = Infection-Slow, R = Resolved, T = On-Treatment, UC = Uninfected-Cleared, UN = Uninfected-Naive. See Supplementary Material section 1 for further details.

We incorporated an access-to-care structure to represent the systematic differences in tuberculosis burden and healthcare access by income.^12^ The access-to-care structure contains high-access-to-care, representing the top three income quintiles (60% of the population) and low-access-to-care, representing the bottom two income quintiles (40% of the population). We assumed no transition between the high- and low-access-to-care classes, and random mixing between them.

To account for the influences of human immunodeficiency virus (HIV) and antiretroviral therapy (ART) on the risk of infection and progression to disease,^13,14^ we included an HIV structure for countries if the proportion of tuberculosis cases among people living with HIV (PLHIV) was at least 15%, and the HIV prevalence was greater than 1% (countries listed in the Supplementary Material Table S5.3). The HIV structure included HIV uninfected, HIV infected and not on ART, and HIV infected and on ART. The tuberculosis mortality rate and risk of progression is increased in both HIV compartments, with greater increases in those not on ART compared to on ART.

We calibrated the model to epidemiologic data in each country separately using history matching with emulation through the *hmer* R package,^15^ generating at least 1000 fitted parameter sets per country. We used the distribution of results produced by these parameter sets to quantify estimation uncertainty.^16^ The model for each country was fit to nine calibration targets in 2019: the tuberculosis incidence rate (overall and by age), tuberculosis case notification rate (overall and by age), tuberculosis mortality rate (overall), the fraction of subclinical tuberculosis among active tuberculosis, and the risk ratio of active tuberculosis in the low-access-to-care group relative to high-access-to-care. Models for countries with the HIV structure were fit to four additional all age HIV targets in 2019: HIV prevalence, ART coverage, tuberculosis incidence rate in PLHIV, and tuberculosis mortality rate in PLHIV.

### Policy Scenarios

#### i. Status Quo No-New-Vaccine Baseline

For each country, separately, a primary baseline with no novel vaccine introduction was simulated, assuming that non-vaccine tuberculosis interventions continue at current levels (‘*Status Quo No-New-Vaccine’* baseline). As reported country-level data includes the high coverage of neonatal BCG vaccination,^17^ this was not explicitly modelled. We assumed that BCG vaccination would not be discontinued over the model time horizon.

#### ii Novel Vaccine Scenarios

Aligning with the product characteristics described in the WHO PPC, we evaluated a novel adolescent/adult and a novel infant vaccine.^5^ Vaccines were assumed to prevent progression to disease and confer 10-years duration of protection on average, with exponential waning. We assumed the adolescent/adult vaccine would “take” in individuals in any infection state at the time of vaccination aside from active tuberculosis disease (i.e., a “pre- and post-infection” vaccine), with 50% vaccine efficacy. We assumed the infant vaccine would “take” in individuals who were uninfected at the time of vaccination (i.e., a “pre-infection” vaccine), with 80% efficacy. Further details are in Supplementary Material section 7.1.

The infant vaccine was implemented in two scenarios, and, separately, the adolescent/adult vaccine was implemented in three scenarios, with assumptions confirmed through consultation with a range of global tuberculosis vaccine stakeholders. The *Basecase* and *Accelerated Scale-up* scenarios included routine neonatal vaccination for the infant vaccine (85% coverage), and routine vaccination of 9-year-olds (80% coverage) with a one-time vaccination campaign for ages ten and older (70% coverage) for the adolescent/adult vaccine. The *Routine Only* scenario (adolescent/adult vaccine only) was introduced through routine vaccination of 9-year-olds (i.e., no campaign).

We evaluated vaccine delivery scenarios by varying the introduction year and scale-up trends between scenarios and countries (Table 1). In the *Basecase* and *Routine Only* scenarios, based on data from historical vaccine introduction, vaccines were introduced in country-specific years and linearly scaled-up to coverage targets over five years once introduced. To estimate introduction years, countries were divided into ‘procuring with Gavi support’ and ‘self-procuring’. Factors influencing the likely timing of vaccine introduction were identified through expert consultation, and included disease burden, prior early adopter status, timelines for Gavi processes, capacity for immunisation, country-specific registration timelines, and commercial prioritisation. A scoring system was applied to each factor, and countries were assigned an aggregate score ranking their likely introduction position. The number of countries introducing the vaccine per year was informed by pneumococcal vaccine scale-up.^18^ See Supplementary Material section 7.2 for details. In the *Accelerated Scale-up* scenarios, to more resemble the pace of COVID-19 vaccine introduction, vaccines were introduced in 2025 in all countries with vaccine coverage targets reached instantly.

**Table 1:**
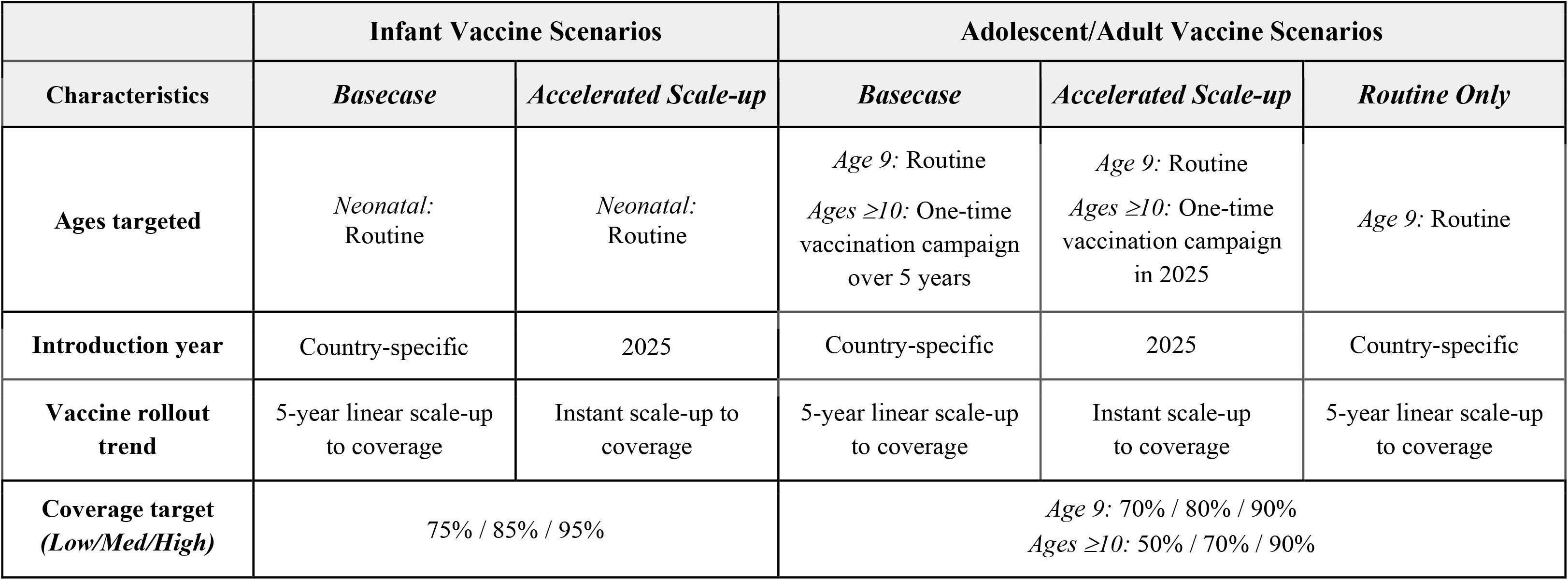
Characteristics of modelled vaccine delivery scenarios

|  | Infant Vaccine Scenarios |  | Adolescent/Adult Vaccine Scenarios |  |  |
| --- | --- | --- | --- | --- | --- |
| Characteristics | *Basecase* | *Accelerated Scale-up* | *Basecase* | *Accelerated Scale-up* | *Routine Only* |
| Ages targeted | *Neonatal:*<br>Routine | *Neonatal:*<br>Routine | *Age 9:* Routine<br>*Ages $\geq 10$:* One-time<br>vaccination campaign<br>over 5 years | *Age 9:* Routine<br>*Ages $\geq 10$:* One-time<br>vaccination campaign<br>in 2025 | *Age 9:* Routine |
| Introduction year | Country-specific | 2025 | Country-specific | 2025 | Country-specific |
| Vaccine rollout trend | 5-year linear scale-up<br>to coverage | Instant scale-up to<br>coverage | 5-year linear scale-up<br>to coverage | Instant scale-up<br>to coverage | 5-year linear scale-up<br>to coverage |
| Coverage target<br>(*Low/Med/High*) | 75% / 85% / 95% |  | *Age 9:* 70% / 80% / 90%<br>*Ages $\geq 10$:* 50% / 70% / 90% |  |  |

#### Health impact indicators

We calculated the cumulative number of tuberculosis treatments, deaths, and cases averted between vaccine introduction and 2050 and calculated tuberculosis incidence and mortality rate reductions in 2050 for each vaccine scenario compared to the *Status Quo No-New-Vaccine* baseline. Incidence rates in 2035 for each vaccine scenario were also estimated to investigate the feasibility of meeting the 2035 End TB target. Results are presented as the median and 95% uncertainty range for all countries modelled, WHO region, World Bank income group,^19^ and WHO tuberculosis burden level.^22^

#### Additional scenario analyses

We conducted scenario analyses to evaluate alternative assumptions regarding the *Status Quo No-New-Vaccine* baseline, vaccine characteristics, and delivery. We simulated vaccine scenarios with lifelong duration of protection for both vaccines, as well as scenarios with adolescent/adult vaccine efficacy increased to 75%. For each scenario, low- and high-coverage targets for five years post-introduction were compared to the medium-coverage targets used for the main analyses. We also explored an alternative baseline: the *2025 End TB No-New-Vaccine* baseline, which assumed strengthening of non-vaccine tuberculosis interventions to meet the 2025 End TB incidence target,^3^ providing an alternative estimate of impact assuming existing measures would be deployed more effectively (Supplementary Material section 6.2).

## Results

### Model calibration and vaccine introduction year

Epidemiologic and demographic data were available to model 115 of 135 LMICs. We successfully calibrated 105 of 115 countries, accounting for 93.3% of global tuberculosis cases and 93.6% of deaths in 2019. Calibrated model incidence and mortality rate trends for WHO regions, WHO TB burden levels, and World Bank income groups are given in Supplementary Material section 9.3. Country-specific vaccine introduction years (used in *Basecase* and *Routine Only* scenarios) ranged between 2028 and 2047 (Supplementary Material Table S8.1), with the earliest Gavi-supported country introducing following a two-year delay compared to the earliest self-procuring. Figure 2 demonstrates the cumulative number of countries introducing per year between 2028 and 2047, with 50% of countries introducing the vaccine by 2034.

**Figure 2:**
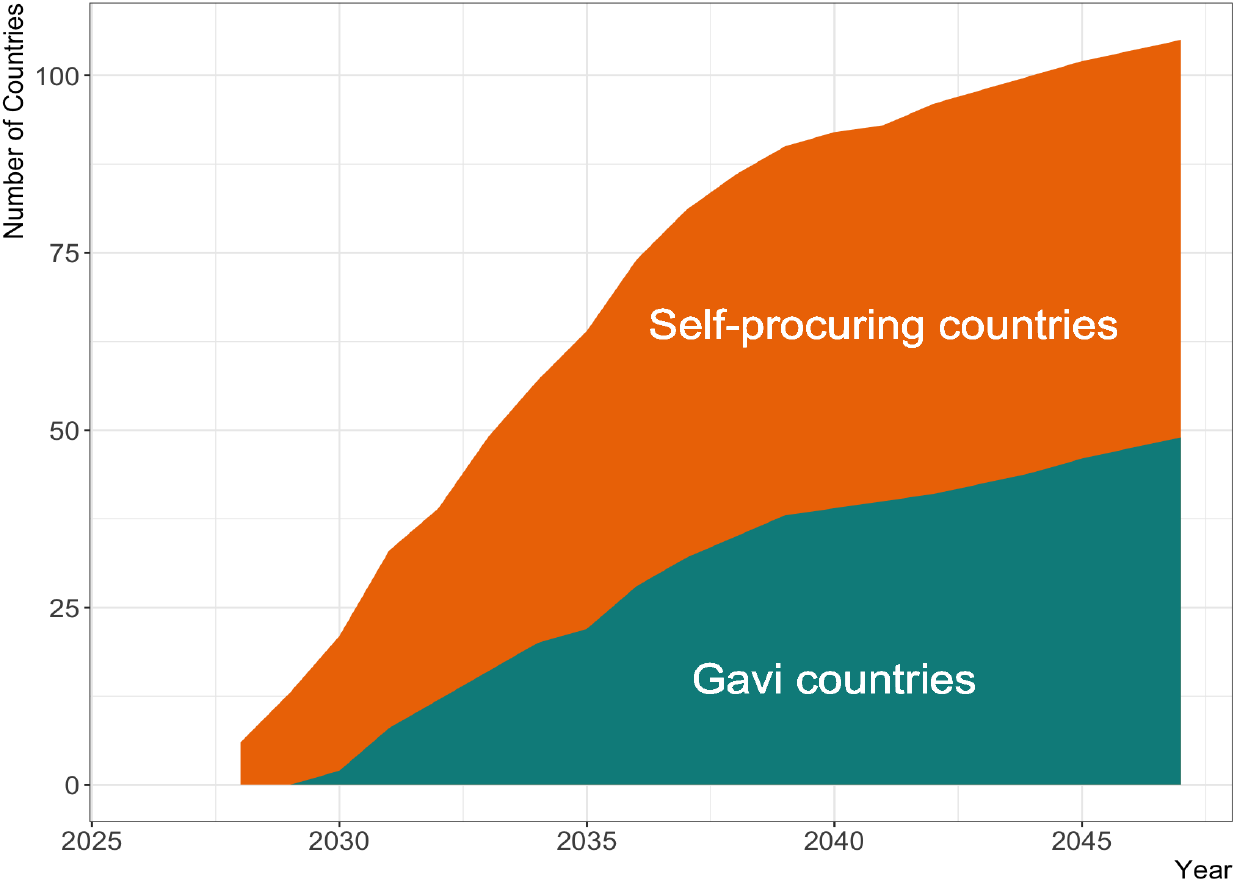
Assumed cumulative number of countries introducing the novel vaccine by year for the *Basecase* and *Routine Only* scenarios. The earliest vaccine introduction occurs in 2028 and the latest in 2047. See Supplementary Material section 7 for full details.

### Health impact for the adolescent/adult vaccine

Our findings suggest that introducing a 50% efficacy adolescent/adult vaccine with 10 years protection in the *Basecase* scenario could avert approximately 44.0 (95% uncertainty range=37.2– 51.6) million cases for all countries compared to the *Status Quo No-New-Vaccine* baseline by 2050, including 34.3 (28.6–40.3) million cases in lower middle-income countries (Table 2, Figure 3). High numbers of cases could be averted in both AFR and the WHO South-East Asian (SEAR) regions, which contribute the highest number to the global total. By 2050, 5.0 (4.6–5.4) million deaths could be averted for all countries, including 2.2 million in SEAR, 2.1 million in AFR, and 4.1 million in lower middle-income countries (Table 2, Figure 3). By 2050, 24.9 (21.9–27.3) million treatments could be averted, with 11.7 (10.1–13.4) million averted treatments in SEAR alone. In the 27 countries categorised by WHO as high-TB-burden of the 105 modelled, 39.8 (33.7–46.7) million cases, 22.6 (19.9–24.8) million treatments, and 4.5 (4.2–4.9) million deaths could be averted by 2050; around ten times higher than those averted in all other countries combined (Table 2, Figure 3).

**Table 2:**
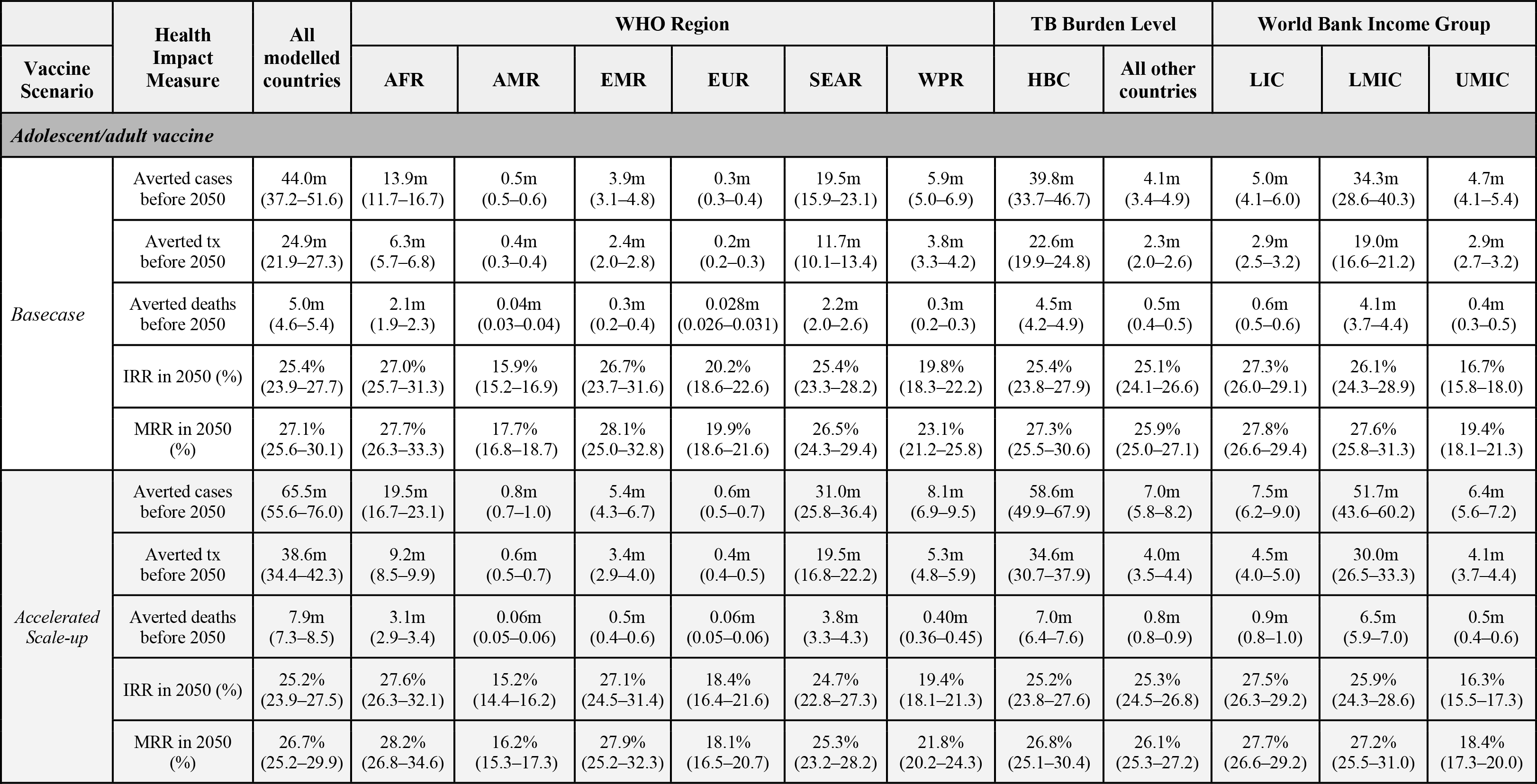
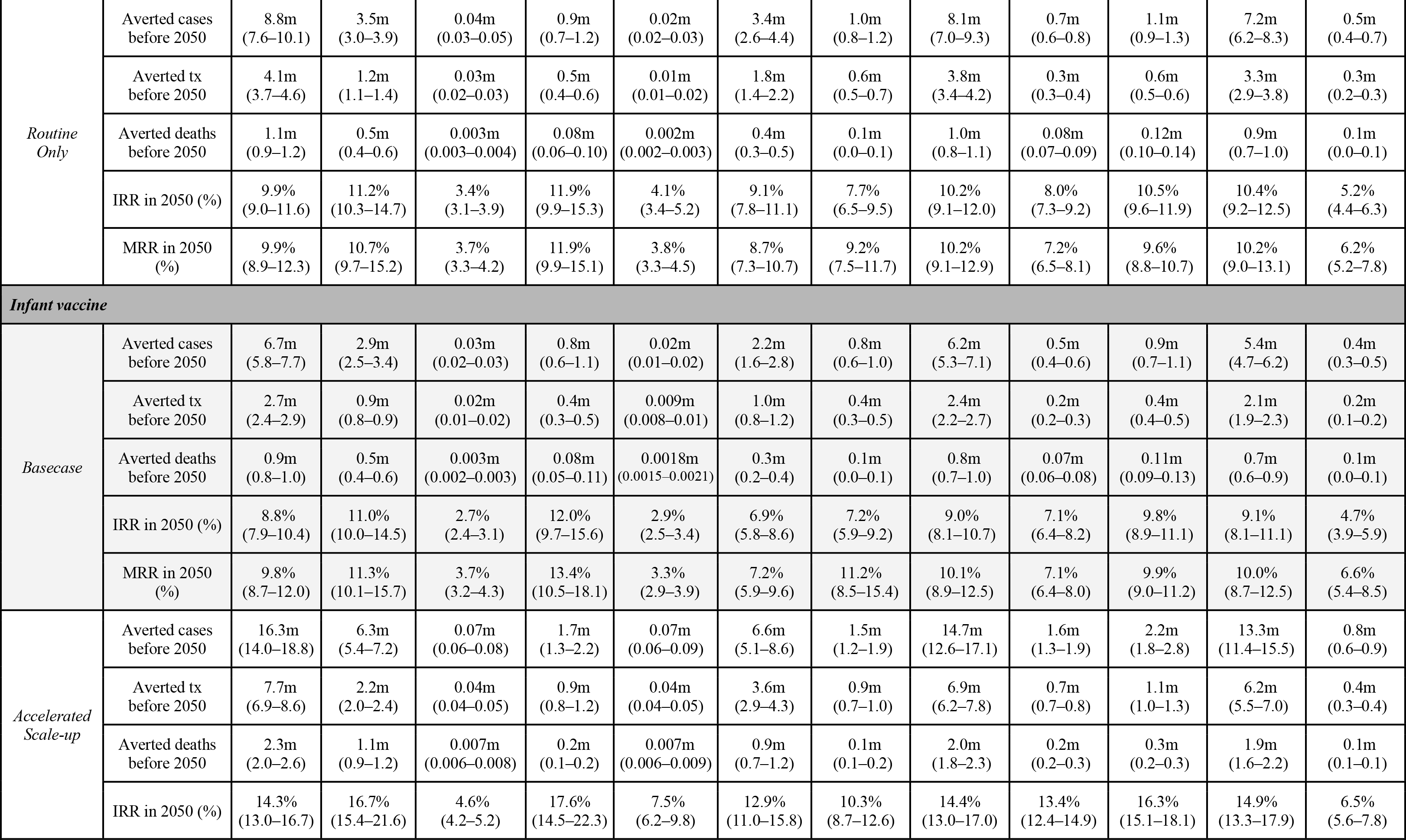
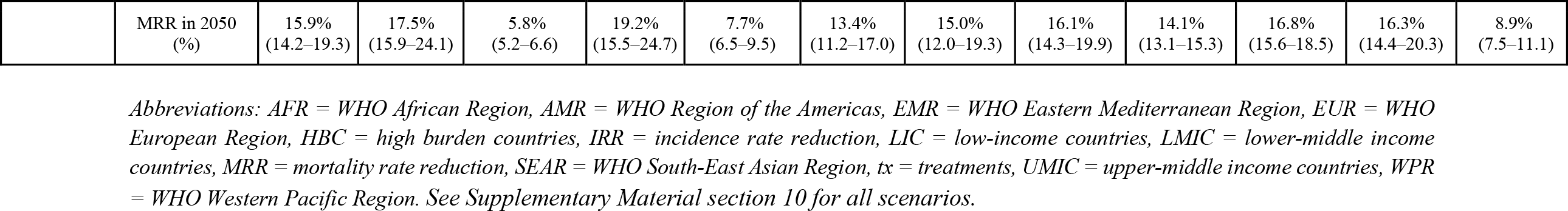
Cumulative cases, treatments, and deaths averted between vaccine introduction and 2050 and incidence and mortality rate reductions in 2050 by WHO region, income level, and tuberculosis burden level for select vaccine scenarios (all 10-years duration of protection and medium coverage targets). Value in cell is the median estimate and 95% uncertainty range.

**Figure 3:**
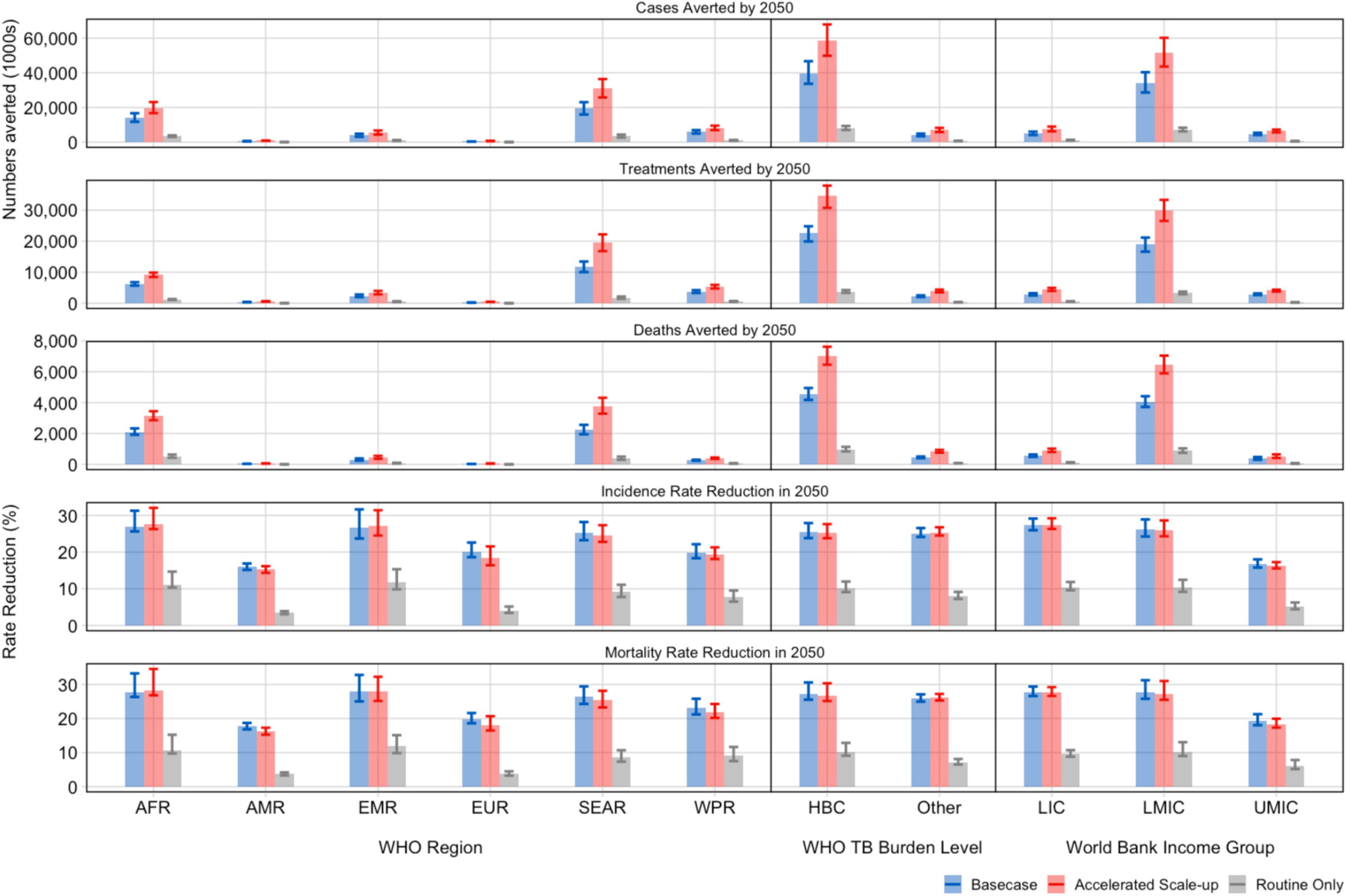
Cumulative cases, treatments, and deaths averted between vaccine introduction and 2050 and incidence and mortality rate reductions in 2050 for the adolescent/adult vaccine with varying delivery scenarios (50% efficacy vaccine, medium coverage, 10-years duration of protection), by WHO region, WHO TB burden level, and World Bank income group. Abbreviations: AFR = WHO African Region, AMR = WHO Region of the Americas, EMR = WHO Eastern Mediterranean Region, EUR = WHO European Region, HBC = high-burden countries, LIC = low-income countries, LMIC = lower-middle income countries, SEAR = WHO South-East Asian Region, UMIC = upper-middle income countries, WPR = WHO Western Pacific Region

Introducing the adolescent/adult vaccine in the *Basecase* scenario was predicted to reduce tuberculosis incidence and mortality rates in 2050 by 25.4% (23.9–27.7%) and 27.1% (25.6– 30.1%), respectively, compared to the *Status Quo No-New-Vaccine* baseline (Table 2). The incidence reduction ranged from 15.9% in the WHO Region of the Americas (AMR) to 27.0% in the African (AFR) region (Table 2, Figure 3). The tuberculosis mortality rate was reduced by 17.7% in AMR to 28.1% in EMR. By income group, relative impact was higher in low-income and lower middle-income countries than in the upper middle-income countries (Table 2, Figure 3).

### Health impact for the infant vaccine

For both the *Basecase* and *Accelerated Scale-up* scenarios, lower impact was estimated for the infant vaccine compared to the adolescent/adult vaccine before 2050, including 0.4–0.6 times incidence and mortality rate reductions by 2050 and 0.1–0.3 times the number of cases, treatments, and deaths averted (Table 2).

### Health impact by delivery scenario

With the *Accelerated Scale-up* scenario, a 50% efficacy adolescent/adult vaccine could prevent 7.9 (7.3–8.5) million deaths–2.9 million more deaths than the *Basecase* delivery–and avert 65.5 (55.6–76.0) million cases and 38.6 (34.4–42.3) million treatments (Table 2, Figure 3). In contrast, by only routinely vaccinating 9-year-olds (*Routine Only* scenario), 8.8 (7.6–10.1) million cases, 4.1 (3.7–4.6) million treatments, and 1.1 (0.9–1.2) million deaths would be averted (Table 2, Figure 3).

### Comparing to the 2035 End TB target

Assuming non-vaccine interventions do not improve in the future (*Status Quo No-New-Vaccine* baseline), the *Basecase* and *Accelerated Scale-up* scenarios of the adolescent/adult vaccine suggest we would reach 34% and 41% of the 2035 global target for reduction in tuberculosis cases, respectively. Assuming the 2025 End TB targets are met before vaccine roll out (*2025 End TB No-New-Vaccine* baseline), progress is increased, with the *Basecase and Accelerated Scale-up* scenarios reaching 82% of the target.

### Additional scenario analyses

Impact results from scenarios with lifelong duration of protection, 75% efficacy, and low- and high-coverage targets are provided in the Supplementary Material section 10. Assuming lower coverage targets or the *2025 End TB No-New-Vaccine* baseline led to reduced health impact, and higher coverage targets, 75% efficacy, or lifelong duration of protection vaccines led to an increased health impact compared to the *Status Quo No-New-Vaccine* baseline, a medium-coverage target, 50% efficacy vaccine, or vaccine with only ten-years protection.

## Discussion

Our results suggest that novel tuberculosis vaccines could substantially reduce the tuberculosis burden in the coming decades. The *Basecase* scenario, in which a 50% efficacy adolescent/adult vaccine was introduced over 20 years, could prevent 44.0 million cases and 5.0 million deaths before 2050, including 2.2 million in the WHO South-East Asian region and 2.1 million in the African region. The more ambitious *Accelerated Scale-up* scenario could prevent 65.5 million cases and 7.9 million deaths (around 60% more). The less ambitious *Routine Only* scenario could prevent 8.8 million cases and 1.1 million deaths (around a fifth).

Impact estimates for vaccine introduction varied by region. While incidence and mortality rate reductions achievable by 2050 were similar between high-TB-burden countries and all other countries, the number of cases, treatments, and deaths averted were around ten times higher than those averted in all other countries, emphasising the need to focus on high-burden countries to maximise health impact. Particularly large numbers of averted cases, treatments, and deaths were predicted in Africa and South-East Asia, and in lower-middle-income countries, arguably populations in the greatest need.

Vaccination campaigns will be important to expedite health gains from vaccination. The *Basecase* and *Routine Only* scenarios offer a direct comparison of implementing with and without a campaign. The *Basecase* scenario averted up to six times as many cases, deaths, and treatments as the *Routine Only* scenario, supporting the need to include a campaign in any future delivery strategy to maximise health impact.

A new vaccine will be an important tool to accelerate progress towards 2035 End TB targets. Conservatively assuming non-vaccine intervention coverage and quality does not improve in the future (*Status Quo No-New-Vaccine* baseline) and roll out from 2028 in line with previous vaccines (before COVID-19) the *Basecase* scenario suggests we could reach around a third of the 2035 global target. More optimistic assumptions, that assume 2025 End TB targets are met before vaccine roll out (*2025 End TB No-New-Vaccine* baseline), combined with the *Accelerated Scale-up* scenario, suggests over 80% of the global 2035 target could be met.

Two recent systematic reviews have highlighted potential health impacts of novel tuberculosis vaccines.^20,21^ Our study expands on the findings of these reviews and addresses some identified gaps. We showed that the adolescent/adult vaccine would have greater and more rapid health impacts than an infant vaccine before 2050. The largest burden of pulmonary tuberculosis disease is most often found in adults,^1^ and the adolescent/adult vaccine targets the age ranges with the highest burden of tuberculosis, compared to the infant vaccine. However, as the health outcomes are measured in 2050, the maximum follow-up time between vaccine delivery and impact calculation is 25 years. Therefore, even with duration of protection increased, it is unlikely that the infant vaccine would be protecting those at highest risk of progressing to active disease in most countries during the time period of our analyses.

Meeting the End TB target to develop and licence an adolescent/adult vaccine by 2025 and introducing at a pace more like that of COVID-19 vaccines (*Accelerated Scale-up*) could save around 60% more deaths, compared to introduction at a historical pace (*Basecase*). The pace of COVID-19 vaccine introduction in LMICs, albeit slower than in high-income countries, has been much faster. At the time of writing, over 10% of the population in at least 75% (102/135) of LMICs have been fully vaccinated in the two years since COVID-19 vaccines have been available, showing that faster vaccine introduction in LMICs is possible with high political will and financial resources.^22^ This is more similar to our *Accelerated Scale-up* scenario, which averted up to 2.9 million more deaths than our *Basecase* scenario, and while the benefits of rolling out a vaccine from 2028 at pre-COVID-19 pace are predicted to be large, the increase in deaths demonstrate the health consequences from failing to rapidly introduce a vaccine. Unlike COVID-19, tuberculosis is a disease of the poor, which does not have the associated novelty, nor the same impact on high-income countries. Therefore, tuberculosis vaccines need concerted, sustained policy attention to overcome these barriers.

Our study has limitations. We successfully calibrated 105 of the 135 LMICs, representing 93% of global tuberculosis incidence, due in part to missing data from 20 countries that prevented calibration from being attempted. Excluding 30 countries will slightly underestimate the number of cases, deaths, and treatments averted, and may bias the generalizability of the relative impact results.

Aligning with the WHO PPC, we assumed the adolescent/adult vaccine would be efficacious (“take”) in both infected and uninfected individuals. However, the majority of previous and current trials have only enrolled either IGRA+ or IGRA-individuals.^6,7^ Ideally, novel vaccines will be safe and effective in both infected and uninfected individuals, as it would be costly and logistically difficult to include testing for tuberculosis infection before vaccine administration. If the realised vaccine will only be efficacious in either IGRA+ or IGRA-, our results will be overestimates of impact, as show in previous work.^8^ We also assumed that vaccine efficacy was equivalent in PLHIV and HIV-naïve. However, vaccines are not always as efficacious in immunocompromised individuals,^23,24^ which would reduce the vaccine impact in countries incorporating the HIV structure.

We evaluated three vaccine scenarios for the adolescent/adult vaccine and two vaccine scenarios for the infant vaccine to provide estimates of vaccine impact under more and less ambitious introduction years and scale-up trends. Our more ambitious scenario, (*Accelerated Scale-up*), is less realistic, as it assumes a vaccine candidate would be ready for licensure, the supply exists, and that countries are positioned to make an introduction decision resulting in immediate uptake, all within the next 3 years. No specific risk groups were vaccinated in our model, however, initial delivery of novel tuberculosis vaccines within countries is likely to be through a targeted approach–a strategy shown to have a large population impact per vaccinated individual.^25–29^ Countries may decide to initially target groups at the highest risk of developing tuberculosis disease or that contribute the most to transmission, while others may focus on vaccinating vulnerable age groups. Understanding how a new tuberculosis vaccine may be introduced in different settings is an important area for future research.

There are remaining gaps that modelling can help to address to provide evidence for investing in tuberculosis vaccine development and delivery to inform the Full Value of Vaccine Assessment.^30^ Estimates of the cost-effectiveness, budget impact, and wider benefits of specific tuberculosis vaccine candidates would support research investment decision making. Future modelling research can help to better understand potential vaccine effectiveness considering a variety of factors, such as age, gender, duration of vaccine protection, and specific risk groups. We included an access-to-care structure in the model to account for differences in tuberculosis burden and healthcare access, which could be used to investigate vaccine targeting by income. Additionally, to maximise the potential evidence available to individual countries, it would be beneficial to create more detailed individual country models to inform vaccine introduction decision making.

Novel tuberculosis vaccines could have a substantial impact, which will vary depending on vaccine and delivery characteristics. Vaccination campaigns will be crucial for rapid impact and accelerated introduction more similar to the pace of COVID-19 vaccine introduction may save around 60% more deaths before 2050. The COVID-19 pandemic has demonstrated the advantage that billions of dollars of investment can have on vaccine research and development, and this provides an illustration of what is possible to achieve with novel tuberculosis vaccines. Continued investment in tuberculosis vaccine research is required to strengthen vaccine development, trials, and manufacturing, and to support prompt introduction and scale-up.

## Data sharing statement

No individual level participant data was used for this modelling study. Epidemiologic data used are available from the *World Health Organization Global TB Report CSV files to download* (https://www.who.int/teams/global-tuberculosis-programme/data) and summarised in the Supplementary Material section 9.2.1. Population estimates and projections are available from the *United Nations Department of Economic and Social Affairs World Population Prospects 2019* (https://population.un.org/wpp/Download/Standard/Population/). Analytic code will be made available at https://doi.org/10.5281/zenodo.6421372 immediately following publication indefinitely for anyone who wishes to access the data for any purpose.

## Supporting information

Supplementary Material_CountryTimelines

## Data Availability

Analytic code will be made available at https://doi.org/10.5281/zenodo.6421372 immediately following publication indefinitely for anyone who wishes to access the data for any purpose.

## Acknowledgements

This is part of a WHO commissioned work on the Full Value Assessment of TB Vaccines (2020/985800-0). We thank all the attendees at the WHO meetings on the Full Value Assessment of TB Vaccines for insightful advice and direction. We thank Philippe Glaziou (WHO) for reviewing and providing helpful suggestions on the paper.

## Author contributions

Conception: RGW, NAM, MJ, RCH, CKW

Data acquisition and preparation: AD, RAC, CM, CKW, DS, AP, SM

Data analysis: RAC, CM, DS, CKW

Interpretation of results: RAC, RGW, NAM, CKW, CM, AP

Manuscript drafting and revisions: RAC, RGW, AP, NAM, CKW, CM, RCH, MJ, SM, DS, RB, NG, MZ, RCWH, BG, MQ, AD, AI

