## Supplementary Material for "The impact of alternative delivery strategies for novel tuberculosis vaccines in low- and middle-income countries: a modelling study"

Rebecca A. Clark et al.

**Table of Contents**

SUPPLEMENTAL METHODS: 3
1. Model structure and equations 3
1.1 Tuberculosis natural history dimension 3
1.1.1 Tuberculosis natural history structure 3
1.1.2 Tuberculosis natural history equations 5
1.1.3 Force of infection equation 6
1.2 HIV and ART structure 7
1.2.1 HIV and ART description 7
1.2.2 HIV and ART diagram 7
1.2.3 HIV and ART equations 8
1.2.4 Natural history equations incorporating HIV and ART 8
1.3 Access to Care Dimension 9
1.3.1 Access to care description 9
2. Model Parameters and Data Sources 10
2.1 Model Parameters and Data Sources 10
2.2 Operationalizing Age Varying Parameters 13
3. Tuberculosis treatment 14
3.1 Tuberculosis treatment initiation 14
3.2 Tuberculosis treatment outcomes 14
4. Calibration methodology 16
5. Low- and middle-income countries 17
5.1 Eligible countries 17
5.2 Countries excluded from attempting calibration 18
5.3 Countries incorporating the HIV structure 19
6. No-New-Vaccine baselines 20
6.1 *Status Quo No-New-Vaccine* baseline 20
6.2 *2025 End TB No-New-Vaccine* baseline 20
7. Vaccination 21
7.1 Vaccine profile 21
7.2 Vaccine delivery scenarios 21
7.2.1 Country-specific introduction years 21
7.2.2 Vaccine coverage targets 24
7.3 Vaccine implementation 26

7.3.1 Vaccine structure 26
7.3.1 Vaccine implementation in the tuberculosis natural history model 28
8. Model outcomes 29
8.1 Epidemiological impact measures 29
8.2 Groupings for reporting model outcomes 29
8.2 Calculating uncertainty 34
SUPPLEMENTAL RESULTS: 35
9. Model Calibration 35
9.1 LMIC countries calibration 35
9.2 WHO Region and World Bank Income Group for Calibrated LMICs 35
9.3 *Status Quo No-New-Vaccine* baseline calibration target values 36
9.4 Calibrated *Status-Quo No-New-Vaccine* baseline trends 45
10. Vaccine Health Impact Results 49
10.1 Incidence and mortality rate reductions and cumulative cases, treatments, and deaths averted 49
References 60

**SUPPLEMENTAL METHODS:**

### 1. Model structure and equations

We created a compartmental tuberculosis vaccine model, which includes separate structures to account for key modelling components required. The structures, or "dimensions" we incorporated into the low- and middle-income country (LMIC) modelling are age, tuberculosis natural history, HIV and ART, and access to care.

#### 1.1 Tuberculosis natural history dimension

##### 1.1.1 Tuberculosis natural history structure

The core natural history model is specified in Figure S1.1. Model parameters used in the tuberculosis natural history dimension and their definitions are provided in Table S3.1.

Those with no previous exposure or infection with *Mtb* [Uninfected-Naive ($U_N$)] could become infected at rate $\lambda_j$ and progress to an Infection-Fast ($I_F$) class following initial infection. From Infection-Fast, three possible pathways were possible: (i) Fast progression to Subclinical Disease ($D_S$), where individuals are infectious with a reduced infectiousness compared to clinical tuberculosis, but display no symptoms of tuberculosis disease;[1] (ii) self-clearance to Uninfected-Cleared ($U_C$), where individuals are no longer infected with *Mtb* and therefore are not at risk of progression to tuberculosis disease without reinfection;[2] or (iii) continue to remain latently infected with a risk of reactivation and progression to disease, albeit at a lower rate than Infection-Fast, by transitioning to the Infection-Slow ($I_S$) class. Those in the Infection-Slow class could self-clear to the Uninfected-Cleared class, be reinfected and return to the Infection-Fast class, or reactivate their infection and progress to Subclinical Disease.

Once in the Subclinical Disease class, individuals could naturally cure (*without* treatment) to the Resolved (R) class, or progress to Clinical Disease ($D_C$), where individuals are infectious and display symptoms of tuberculosis disease. Treatment initiation from Clinical Disease to On-Treatment (T) began in 1960 and increased following a sigmoid curve to 2019, with average treatment duration assumed to be 6 months.[3,4] Treatment completions transitioned to the Resolved class and treatment non-completions returned to Clinical Disease. Deaths occurring on-treatment and in clinical disease counted toward the total number of tuberculosis deaths during the year. Those with clinical disease could also naturally cure to the resolved class. Individuals in the Resolved class could be reinfected or relapse to Subclinical Disease but could *not* enter Infection-Fast or Infection-Slow directly. We assumed that the infection and resolved classes are partially protected against reinfection.[5,6] In those who have self-cleared, we assumed the level of protection against reinfection is half of the protection against reinfection for the infection and resolved classes.

Age was modelled in single years from ages 0 to 79 and aggregated into two categories for ages 80 to 89, and ages 90 to 99. Births and ageing occurred at the beginning of each year.

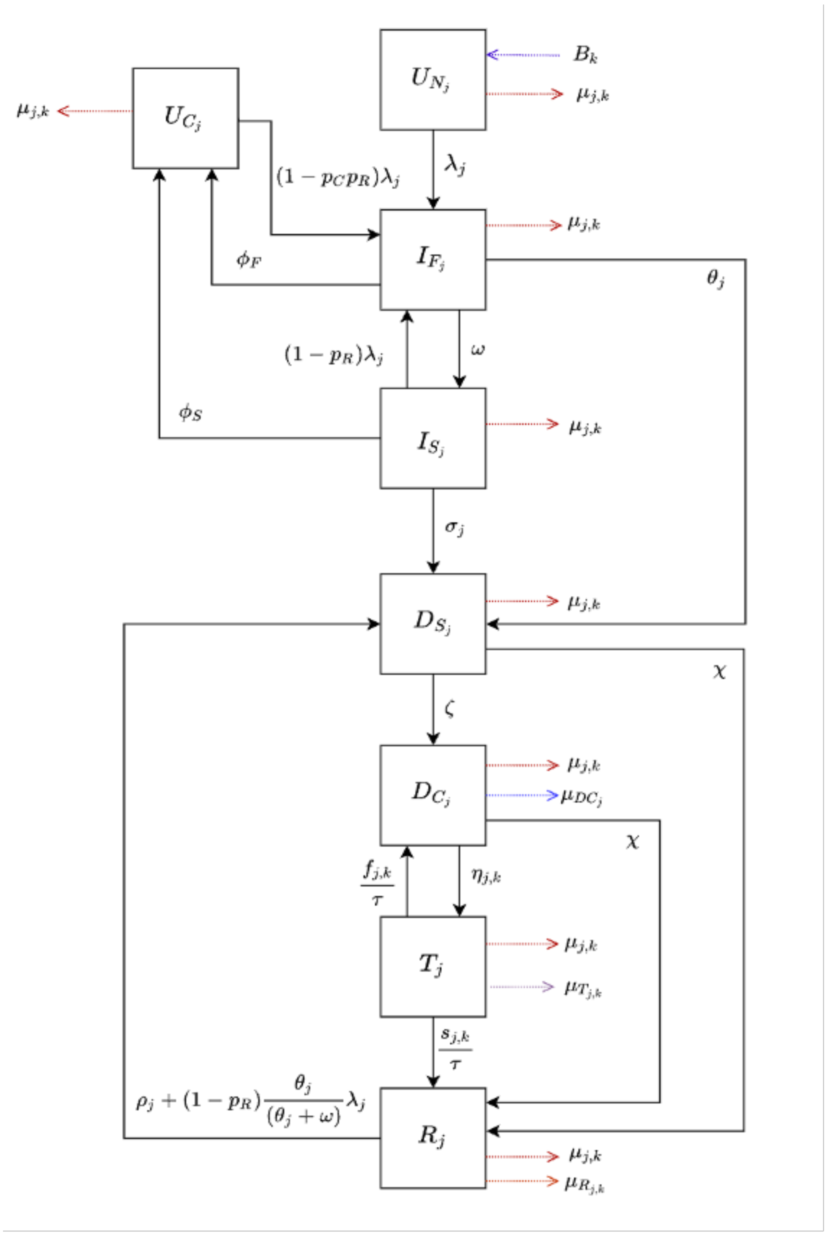


**Figure S1.1** Tuberculosis natural history model

*Abbreviations: $D_C$ = Clinical Disease; $D_S$ = Subclinical Disease; $I_F$ = Infection-Fast; $I_S$ = Infection-Slow; R = Resolved; T = On-Treatment; $U_C$ = Uninfected-Cleared; $U_N$ = Uninfected-Naive.*

#### 1.1.2 Tuberculosis natural history equations

$$Age\ j = 0$$

$$\frac{dU_{N_j}}{dt} = B_k - (\lambda_j + \mu_{j,k})U_{N_j}$$

$$Age\ j \neq 0$$

$$\frac{dU_{N_j}}{dt} = -(\lambda_j + \mu_{j,k})U_{N_j}$$

$$\frac{dU_{C_j}}{dt} = \phi_F I_{F_j} + \phi_S I_{S_j} - [(1 - p_C p_R)\lambda_j + \mu_{j,k}]U_{C_j}$$

$$\frac{dI_{F_j}}{dt} = \lambda_j U_{N_j} + (1 - p_C p_R)\lambda_j U_{C_j} + [(1 - p_R)\lambda_j]I_{S_j} - (\phi_F + \omega + \theta_j + \mu_{j,k})I_{F_j}$$

$$\frac{dI_{S_j}}{dt} = \omega I_{F_j} - (\phi_S + \sigma_j + (1 - p_R)\lambda_j + \mu_{j,k})I_{S_j}$$

$$\frac{dD_{S_j}}{dt} = \theta_j I_{F_j} + \sigma_j I_{S_j} + [\rho_j + (1 - p_R)\frac{\theta_j}{\theta_j + \omega}\lambda_j]R_j - (\chi + \zeta + \mu_{j,k})D_{S_j}$$

$$\frac{dD_{C_j}}{dt} = \zeta D_{S_j} + \frac{f_{j,k}}{\tau}T_j - (\chi + \eta_{j,k} + \mu_{DC_j} + \mu_{j,k})D_{C_j}$$

$$\frac{dT_j}{dt} = \eta_{j,k} D_{C_j} - \left(\frac{s_{j,k} + f_{j,k}}{\tau} + \mu_{T_{j,k}} + \mu_{j,k}\right)T_j$$

$$\frac{dR_j}{dt} = \frac{s_{j,k}}{\tau}T_j + (D_{S_j} + D_{C_j})\chi - [\rho_j + (1 - p_R)\frac{\theta_j}{\theta_j + \omega}\lambda_j + \mu_{R_j} + \mu_{j,k}]R_j$$

#### 1.1.3 Force of infection equation

The equation for the age-specific force of infection ($\lambda_j$), or the rate at which Uninfected-Naïve individuals acquire *Mtb* infection in the population, is given below. Clinically, infection with *Mtb* can present as pulmonary tuberculosis which impacts the lungs, and extrapulmonary tuberculosis (EPTB) which occurs in sites other than the lungs.[7,8] EPTB is not infectious, and as we are modelling *Mtb* transmission we would want to exclude it. However, the WHO tuberculosis estimates which we calibrated to include both EPTB and pulmonary tuberculosis. Therefore, instead of excluding EPTB from the model, we discounted the force of infection by the proportion of incident cases that are EPTB to account for the fact that they are not infectious and calibrated to the targets that include both EPTB and pulmonary tuberculosis. We also discounted the force of infection to account for the relative reduced infectiousness of subclinical disease compared to clinical disease.

$$\lambda_j = p_T \times \sum_{y=1}^{n_{ygroups}} C[m, y] \times \left( \frac{(1 - ep)(TD_{C_y} + rTD_{S_y})}{N_y} \right)$$

where:

$$N_y = \sum_{j=j_{min}}^{j_{max}} U_{N_j} + U_{C_j} + I_{F_j} + I_{S_j} + D_{S_j} + D_{C_j} + T_j + R_j$$

$$TD_{C_y} = \sum_{j=j_{min}}^{j_{max}} D_{C_j} \qquad TD_{S_y} = \sum_{j=j_{min}}^{j_{max}} D_{S_j}$$

| Parameter | Definition |
|---|---|
| $j$ | Age of individual (in years) |
| $y$ | Age group of contact |
| $n_{ygroups}$ | Number of contact age groups |
| $p_T$ | Accounting for the probability of transmission per infectious contact |
| $m$ | Age group of individual |
| $C[m, y]$ | Contact rate between individual of age group $m$ and contact of age group $y$ from Prem et al.[9] |
| $ep$ | Average proportion of tuberculosis cases that are extrapulmonary |
| $r$ | Proportional reduction in infectiousness from subclinical disease |
| $TD_{C_y}$ | Total population in a clinical disease class in age group $y$ |
| $TD_{S_y}$ | Total population in a subclinical disease class in age group $y$ |
| $N_y$ | Total population alive in age group $y$ |
| $j_{min}$ | Minimum age $j$ within age group $y$ |
| $j_{max}$ | Maximum age $j$ within age group $y$ |

### 1.2 HIV and ART structure

#### 1.2.1 HIV and ART description

In order to account for the influences of human immunodeficiency virus (HIV) and antiretroviral therapy (ART) on the risk of infection with *Mtb* and progression to tuberculosis disease,[10,11] we have implemented an HIV structure composed of 3 compartments: HIV uninfected [$HIV_0$], people living with HIV (PLHIV) not on ART [$HIV_1$], and PLHIV on ART [ART]. HIV uninfected individuals were diagnosed with HIV and moved from the $HIV_0$ compartment to the $HIV_1$ compartment with rate $\lambda_H$. Within the $HIV_1$ compartment, there is a higher risk of tuberculosis progression and an increased tuberculosis mortality rate compared to the $HIV_0$ compartment. PLHIV are initiated on treatment with ART from $HIV_1$ following a sigmoid trend. The increases in tuberculosis mortality rate and tuberculosis progression are reduced while in ART compared to $HIV_1$, but still higher than in $HIV_0$. ART also reduces the HIV mortality rate. Model parameters used in the HIV and ART structure and their definitions are provided in Table S3.1.

#### 1.2.2 HIV and ART diagram

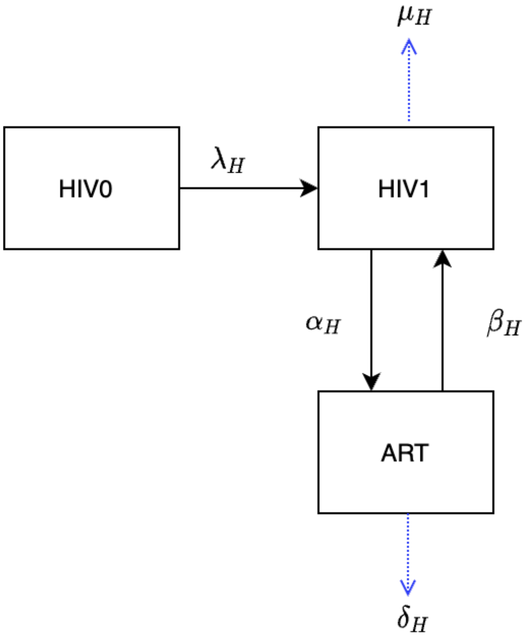


**Figure S1.2** HIV and ART structure

##### 1.2.3 HIV and ART equations

$$\frac{d\ \mathrm{HIV}_0}{dt} = -\lambda_H \mathrm{HIV}_0$$

$$\frac{d\ \mathrm{HIV}_1}{dt} = \lambda_H \mathrm{HIV}_0 + \beta_H \mathrm{ART} - (\alpha_H + \mu_H)\mathrm{HIV}_1$$

$$\frac{d\ \mathrm{ART}}{dt} = \alpha_H \mathrm{HIV}_1 - (\beta_H + \delta_H)\mathrm{ART}$$

##### 1.2.4 Natural history equations incorporating HIV and ART

Let $p_{adj} = (1 + th1(\theta_{mul} - 1))$

| $Age\ j = 0$ | $Age\ j \neq 0$ |
|---|---|
| $\frac{dU_{N_j}}{dt} = B_k - (\lambda_j + \mu_{j,k})U_{N_j}$ | $\frac{dU_{N_j}}{dt} = -(\lambda_j + \mu_{j,k})U_{N_j}$ |

$$\frac{dU_{C_j}}{dt} = \phi_F I_{F_j} + \phi_S I_{S_j} - [(1 - p_C p_R)\lambda_j + \mu_{j,k}]U_{C_j}$$

$$\frac{dI_{F_j}}{dt} = \lambda_j U_{N_j} + (1 - p_C p_R)\lambda_j U_{C_j} + [(1 - p_R)\lambda_j]I_{S_j} - (\phi_F + \omega + p_{adj}\theta_j + \mu_{j,k})I_{F_j}$$

$$\frac{dI_{S_j}}{dt} = \omega I_{F_j} - (\phi_S + p_{adj}\sigma_j + (1 - p_R)\lambda_j + \mu_{j,k})I_{S_j}$$

$$\frac{dD_{S_j}}{dt} = p_{adj}\theta_j I_{F_j} + p_{adj}\sigma_j I_{S_j} + [p_{adj}\rho_j + (1 - p_R)\tfrac{p_{adj}\theta_j}{p_{adj}\theta_j + \omega}\lambda_j]R_j - (\chi + \zeta + \mu_{j,k})D_{S_j}$$

$$\frac{dD_{C_j}}{dt} = \zeta D_{S_j} + \tfrac{f_{j,k}}{\tau}T_j - (\chi + \eta_{j,k} + m_{adj}\mu_{DC_j} + \mu_{j,k})D_{C_j}$$

$$\frac{dT_j}{dt} = \eta_{j,k}D_{C_j} - \left(\tfrac{s_{j,k} + f_{j,k}}{\tau} + m_{adj}\mu_{T_{j,k}} + \mu_{j,k}\right)T_j$$

$$\frac{dR_j}{dt} = \tfrac{s_{j,k}}{\tau}T_j + (D_{S_j} + D_{C_j})\chi - [p_{adj}\rho_j + (1 - p_R)\tfrac{p_{adj}\theta_j}{p_{adj}\theta_j + \omega}\lambda_j + m_{adj}\mu_{R_j} + \mu_{j,k}]R_j$$

### 1.3 Access to Care Dimension

#### 1.3.1 Access to care description

The access to care dimension is incorporated to allow for the negative correlation between tuberculosis burden and health care access to prevent the overestimation of vaccine impact, as well as to facilitate future analyses of equity implications of vaccine introduction. The access to care dimension contains 2 classes: high-access-to-care, representing the top 3 quintiles (60% of the population) and low-access-to-care, representing the bottom 2 quintiles (40% of the population). We assumed that there was no transition between the high- and low-access-to-care classes, as well as assuming random mixing between the high-access-to-care and low-access-to-care classes.

To constrain relative burden between access-to-care classes, we calibrated the relative tuberculosis prevalence in the high-access-to-care class to the low-access-to-care class in 2019. The calibration target, 0.674, was calculated as a weighted average from ten studies, with lower and upper bounds (0.575–0.801) representing the 25th and 75th percentiles of the datasets.[12–21]

To incorporate access to care into our model, we assume that the differences in tuberculosis burden between classes are due to differences in the force of infection, the rate of care-seeking (i.e., tuberculosis treatment initiation), and the rate of tuberculosis progression. We assume relative to the low-access-to-care strata, the high-access-to-care strata has a reduced force of infection per contact, an increased rate of treatment initiation, and a reduced rate of tuberculosis progression. Differential burden was implemented by introducing a new parameter $p_E$, such that $p_E \in [0, 1]$ for the high-access-to-care and $p_E = 0$ and included within the model natural history structure as described in Table S1.1. This new parameter was fitted during calibration.

**Table S1.1** Implementing the access-to-care parameter

| | **Access-to-Care** |
|---|---|
| **Force of infection** | $p_T \times \sum_{y=1}^{n_{ygroups}} (1 - p_E) \times C[m, y] \times \left( \frac{(1-ep)(TD_{C_y} + rTD_{S_y})}{N_y} \right)$ |
| **Treatment Initiation Rate** | $\frac{\eta_{j,k}}{(1-p_E)}$ |
| **Rate of Tuberculosis Progression** | $(1 - p_E) \times \theta_j$<br>$(1 - p_E) \times \sigma_j$<br>$(1 - p_E) \times \rho_j$ |

### 2. Model Parameters and Data Sources

#### 2.1 Model Parameters and Data Sources

Parameters used in the natural history model structure and the HIV and ART model structure are provided in Table S2.1 below, along with their definitions, sources, and information on whether the parameter is fixed or varied (as well as whether they are varied by age or time) during calibration. Further details about how the age varying parameters are implemented are provided in section 2.2.

**Table S2.1** Demographic and tuberculosis natural history parameters and definitions

| Description | Units | Symbol | Prior | Fixed or Varying During Calibration | Age Varying | Time Varying | Source |
|---|---|---|---|---|---|---|---|
| ***Births and deaths (excluding on-treatment mortality)*** | | | | | | | |
| Birth rate | Per year | $B_k$ | United Nations World Population Prospects population estimates and projections | Fixed | No | Yes | 22 |
| Background mortality rate | Per year | $\mu_{j,k}$ | Calculated in the model from United Nations population estimates and projections | Fixed | Yes, age specific mortality rates from demographic dataset | Yes | 22 |
| Mortality rate for clinical tuberculosis disease | Per person per year | $\mu_{DC_j}$ | (0–0.178) | Varying | Yes, value for children is greater than value for adults | No | 23 |
| Mortality rate post-tuberculosis disease | Per person per year | $\mu_{R_j}$ | $0.22\mu_{j,k}$ | Fixed relationship | Yes, because $\mu_{j,k}$ varies | Yes, because $\mu_{j,k}$ varies | 24 |
| ***Natural history*** | | | | | | | |
| Force of infection | Per year | $\lambda_j$ | Fitted | Fixed equation | Yes, age specific contact rates[9] | No | *Calculated* |
| Probability of transmission per infectious contact | - | $p_T$ | (0–0.0068) | Varying | No | No | *Assumed* |

| | | | | | | | |
|---|---|---|---|---|---|---|---|
| Fraction of total tuberculosis disease that is extrapulmonary | - | $ep$ | Country-specific average of previous 3 years | Fixed | No | No | [25,26] |
| Infectiousness of subclinical relative to clinical tuberculosis | - | $r$ | 0.80 | Fixed | No | No | [27] |
| Rate of self-clearance from $I_F$ to $U_C$ | Per person per year | $\phi_F$ | 0.00000140 | Fixed | No | No | [2] |
| Rate of self-clearance from $I_S$ to $U_C$ | Per person per year | $\phi_S$ | (0.0254–0.0467) | Varying | No | No | [2] |
| Rate of fast progression to disease, by age | Per person per year | $\theta_j$ | (0.0696–0.111) | Varying | Yes, value for children is less than value for adults | No | [2] |
| Rate from $I_F$ to $I_S$ | Per person per year | $\omega$ | 0.5 | Fixed | No | No | *Defined* |
| Rate of reactivation from $I_S$, by age | Per person per year | $\sigma_j$ | (0.000135–0.00113) | Varying | Yes, value for children is less than value for adults | No | [2] |
| Rate of progression from $D_S$ to $D_C$ | Per person per year | $\zeta$ | (0–1) | Varying | No | No | *Assumed* |
| Rate of natural cure from $D_C$ and $D_S$ | Per person per year | $\chi$ | (0.1–0.25) | Varying | No | No | [28,29] |
| Rate of relapse from R, by age | Per person per year | $\rho_j$ | (0.0001–0.07) | Varying | Yes, value for children is less than value for adults | No | [30–32] |
| ***Treatment outcome parameters*** | | | | | | | |
| Treatment duration | Number of years | $\tau$ | 0.5 | Fixed | No | No | [3,4] |
| Rate of on-treatment mortality | Per person per year | $\mu_{T_j} = \frac{\kappa_j}{\tau}$ | Country-specific | Varying | Yes, value for children greater than value for adults | Yes | [33] |
| Rate of treatment completion | Per person per year | $\frac{s_j}{\tau}$ | Country-specific | Fixed equation | Yes, indirectly scaled by $s_{Age}$ | Yes | [33] |

| | | | | | | | |
|---|---|---|---|---|---|---|---|
| Rate of treatment non-completion | Per person per year | $\frac{f_j}{\tau}$ | Country-specific | Fixed equation | Yes, indirectly scaled by $s_{Age}$ | Yes | [33] |
| ***Protection parameters*** | | | | | | | |
| Protection from reinfection $I_S$, $I_F$, R | - | $p_R$ | (0.6–0.85) | Varying | No | No | [5,6,28,29,34] |
| Relative protection from reinfection for self-clearance compared to $p_R$ | - | $p_C$ | 0.50 | Fixed | No | No | *Assumed* |
| SES parameter | - | $p_E$ | (0–1) | Varying | No | No | *Assumed* |
| ***HIV parameters*** | | | | | | | |
| HIV incidence rate | Per year | $\lambda_H$ | (0–1) | Varying | No | No | *Assumed* |
| Rate of ART initiation | Per year | $\alpha_H$ | (0–10) | Varying | No | No | *Assumed* |
| Rate of ART discontinuation | Per year | $\beta_H$ | 0.074 | Fixed | No | No | [35,36] |
| Mortality rate from HIV not on ART | Per year | $\mu_H$ | 0.10 | Fixed | No | No | [37] |
| Mortality rate from HIV on ART | Per year | $\delta_H$ | 0.026 | Fixed | No | No | [38] |
| Relative increase in progression rate for $HIV_1$ | - | $\theta_{mul}$ | (3.94–14.45) | Varying | No | No | [39] |
| Relative reduction in $\theta_{mul}$ for HIV and ART compartments | - | $th1$ | $HIV_0$ = 0<br>$HIV_1$ = 1.00<br>ART = 0.35 | Fixed | No | No | [11] |
| Relative mortality rate adjustment for HIV and ART compartments | - | $m_{adj}$ | $HIV_0$ = 1.00<br>$HIV_1$ = 1.50<br>ART = 1.15 | Fixed | No | No | [11,23,40,41] |

### 2.2 Operationalizing Age Varying Parameters

We assume that aspects of tuberculosis natural history and mortality vary by age. This is implemented by stratifying certain natural history parameters by age and applying age-specific prior ranges and relative constraints during calibration.[42] The following table describes the method used to operationalise the age varying differences in parameters between adults, defined as all ages greater than and equal to 15, and children, defined as all ages less than 15. Age varying for the treatment initiation rate is described in the section 3.

**Table S2.2** How age varying parameters are operationalised

| Parameter | Parameter prior range | Age-specific constraints during calibration | Sample the corresponding age scaling parameter | Adults ($\theta_{A15}$) | Children ($\theta_{A0}$) |
|---|---|---|---|---|---|
| $\theta_j$ Rate per year of fast progression | Sample from $(0.0696, 0.111)$ | Retain if value for children is less than value for adults (but still within the prior range) | Sample $j_1$ from $(0, 1)$ | Sample $\theta_{A15}$ from $(0.0696, 0.111)$ | $\max(0.0696, \theta_{A15} \times j_1)$ |
| $\sigma_j$ Rate per year of reactivation | Sample from $(0.000135, 0.00113)$ | Retain if value for children is less than value for adults (but still within the prior range) | Sample $j_2$ from $(0, 1)$ | Sample $\sigma_{A15}$ from $(0.000135, 0.00113)$ | $\max(0.000135, \sigma_{A15} \times j_2)$ |
| $\rho_j$ Rate per year of relapse | Sample from $(0.0001, 0.07)$ | Retain if value for children is less than value for adults (but still within the prior range) | Sample $j_3$ from $(0, 1)$ | Sample $\rho_{A15}$ from $(0.0001, 0.07)$ | $\max(0.0001, \rho_{A15} \times j_3)$ |
| $\mu_{DC_j}$ Clinical tuberculosis mortality rate per year | Sample from $(0, 0.178)$ | Retain if value for children is greater than value for adults | Sample $S_{Age}$ from $(0, 1)$ | $\mu_{DC_{A0}} \times S_{Age}$ | Sample $\mu_{DC_{A0}}$ from $(0, 0.178)$ |
| $\mu_{T_j} = \frac{\kappa_j}{\tau}$ On-treatment mortality rate per year | Sample from $(0, \frac{\kappa_{max}}{\tau})$ | Retain if value for children is greater than value for adults | Sample $S_{Age}$ from $(0, 1)$ | $\frac{\kappa_{A0}}{\tau} \times S_{Age}$ | Sample $\kappa_{A0}$ from $(0, \kappa_{max})$ |

### 3. Tuberculosis treatment

#### 3.1 Tuberculosis treatment initiation

Tuberculosis treatment was assumed to start in 1960, aligned roughly with the discovery and widespread use of rifampicin, and increase following a sigmoid curve (Figure S3.1) to 2019. The treatment initiation rate parameter, $\eta_j$, represents the age specific rate of treatment initiation from the clinical disease compartment to the on-treatment compartment. During calibration, a country-specific value for $\eta_j$ was sampled between 0 and 1. $\eta_j$ was multiplied by an age scaling parameter for children, $j_4$, also sampled between 0 and 1, to ensure that the treatment initiation rate in children was less than in adults. This was then multiplied by the value of the sigmoid curve at each year. The treatment initiation rate was calibrated to the country-specific notification rate in 2019 overall and by age reported by the WHO.[25] Due to inconsistencies in the availability of private sector treatment notification data, the contribution of the private sector was not explicitly represented in our model aside from where it had already been incorporated in WHO estimates.

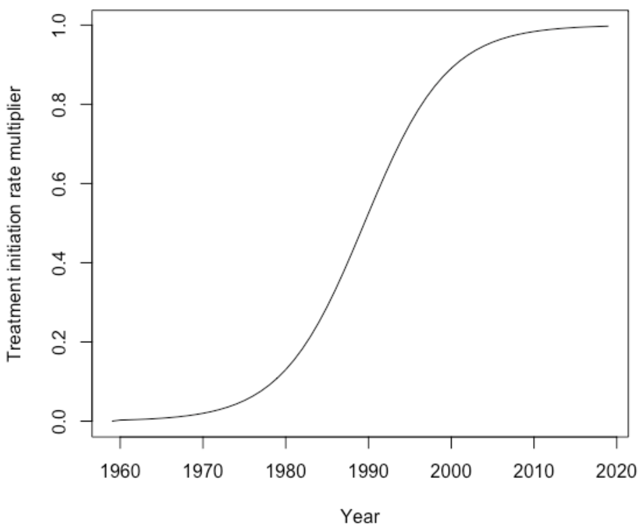


**Figure S3.1** Sigmoid curve representing the scale-up in tuberculosis treatment from 1960-2019

#### 3.2 Tuberculosis treatment outcomes

There are three possible exits from the on-treatment compartment: treatment completion, which progresses to the resolved compartment, treatment non-completion, which returns to the clinical disease compartment, and on-treatment mortality, which counts toward tuberculosis mortality. To account for the variability in tuberculosis treatment outcomes and possible underreporting of on-treatment mortality, we used the following country-specific process:

1. For each country separately, the proportion of treatment completions out of the sum of the number of treatment completions and non-completions (previously called "treatment failures") was calculated and averaged over the years of available data from WHO.

Let $s_R$ = Reported number of treatment completions, $f_R$ = Reported number of treatment non-completions

Note: reported number of treatment non-completions included 0.5 × (reported number lost to follow up)

$$\mathrm{SFR} = \frac{s_R}{s_R + f_R}$$

Ex. In India, averaged over 2012–2018, SFR = 0.96. This can be interpreted as of the sum of treatment completions and non-completions, on average, 96% are completions and 4% non-completions.

2. A value for child treatment mortality ($\kappa_{A0}$) was sampled between 0 and $(2 \times \text{Average Reported Treatment Mortality})$. The average reported treatment mortality is multiplied by 2 to give an upper bound in the case of unreported data.

   Ex. For India, $\kappa_{A0} \in (0, 0.135)$

3. The age multiplier, $S_{Age}$, was sampled from $(0, 1)$, and multiplied by $\kappa_{A0}$ to calculate the adult treatment mortality

$$\kappa_{A15} = \kappa_{A0} \times S_{Age}$$

4. The success and failure rates per year were calculated as in Table S3.1

**Table S3.1** Calculating treatment outcome parameter values for adults and children

| Parameter | Adults | Children |
|---|---|---|
| $\kappa_j$ On-treatment mortality fraction | $\kappa_{A0} \times S_{Age}$ | Sample $\kappa_{A0}$ from 0 to 2 x Average mortality on-treatment |
| $s_j$ On-treatment completion fraction | $(1 - \kappa_{A15})\mathrm{SFR}$ | $(1 - \kappa_{A0})\mathrm{SFR}$ |
| $f_j$ On-treatment non-completion fraction | $(1 - \kappa_{A15})(1 - \mathrm{SFR})$ | $(1 - \kappa_{A0})(1 - \mathrm{SFR})$ |

5. Each of the parameters in Table S3.1 were divided by $\tau$ to obtain the on-treatment mortality rate per year, on-treatment completion rate per year, and on-treatment non-completion rate per year.

### 4. Calibration methodology

The model was fitted to calibration targets using history matching with emulation, a relatively new calibration method that allows us to explore high-dimensional parameter spaces efficiently and robustly.[43–46] History matching progresses as a series of iterations, called waves, where implausible areas of the parameter space, i.e., areas that are unable to give a match between the model output and the empirical data, are found and discarded. In order to identify implausible parameter sets, emulators are used. Emulators are statistical approximations of model outputs that are built using a modest number of model runs. Emulators provide an estimate of the value of the model at any parameter set of interest, with the advantage that they are orders of magnitude faster than the model.

History matching with emulation, implemented through the *hmer* package in R,[47] considerably reduced the size of the parameter space to investigate. Rejection sampling was then performed on the reduced space to identify at least 1000 parameter sets that matched all targets for each country.

If countries were unable to find at least 1000 fully fitted parameter sets using history matching with emulation, they were subsequently assessed using an Approximate Bayesian Computation using Markov Chain Monte Carlo method (ABC-MCMC). ABC-MCMC was conducted using the *easyABC* package in R, modified by the Sebastian Funk, Gwenan Knight, and the Tuberculosis Modelling group at LSTHM for adaptive sampling and to accept seeded parameter values.[48,49] We used parameter sets with the maximum number of targets fitted using history matching with emulation as a starting seed, with the ABC-MCMC algorithm continuously adapting using the last 1000 points and the noise factor set to 0.0001.

### 5. Low- and middle-income countries

#### 5.1 Eligible countries

To decide which low and middle income countries (LMICs) to model on, we used the 135 LMICs indicated on the 2019 World Bank Income Level classifications.[50] The 135 countries were broken down into 29 low-income countries (LICs), 50 lower middle-income countries (LMICs), and 56 upper middle-income countries (UMICs). Distribution by World Bank Income and WHO region are shown below in Table S5.1.

**Table S5.1** LMICs by WHO Region and World Bank Income Level 2019

| **WHO Region** | **World Bank Income Level 2019** | | | ***Total*** |
|---|---|---|---|---|
| | LIC | LMIC | UMIC | |
| AFR | 21 | 19 | 5 | 45 |
| AMR | 1 | 4 | 20 | 25 |
| EMR | 5 | 6 | 5 | 16 |
| EUR | 1 | 4 | 15 | 20 |
| SEAR | 1 | 7 | 3 | 11 |
| WPR | 0 | 10 | 8 | 18 |
| ***Total*** | 29 | 50 | 56 | 135 |

*Abbreviations: AFR = WHO African Region, AMR = WHO Region of the Americas, EMR = WHO Eastern Mediterranean Region, EUR = WHO European Region, LIC = low-income countries, LMIC = lower-middle income countries, SEAR = WHO South-East Asian Region, UMIC = upper-middle income countries, WPR = WHO Western Pacific Region*

### 5.2 Countries excluded from attempting calibration

Of the 135 LMIC countries, 20 countries were excluded from attempting calibration due to the following reasons:

**Table S5.2** Countries excluded from attempting calibration and reasons for exclusion

| Country | Reason for Exclusion |
|---|---|
| American Samoa | Missing critical epidemiological data for calibration, no contact matrices available |
| Belize | No case notification or incidence data for children |
| Comoros | No case notification data |
| Democratic Republic of the Congo | No case notification data by age |
| Republic of the Congo | No population estimates |
| Democratic People's Republic of Korea | Missing critical epidemiological data for calibration |
| Djibouti | No case notification data by age |
| Dominica | Missing critical epidemiological data for calibration, no contact matrices available |
| Federated States of Micronesia | Missing critical epidemiological data for calibration, no contact matrices available |
| Grenada | Missing critical epidemiological data for calibration, no contact matrices available |
| Haiti | Missing 2020 contact matrix |
| Kiribati | Missing 2020 contact matrix |
| Kosovo | Missing critical epidemiological data for calibration |
| Lebanon | Missing 2020 contact matrix |
| Marshall Islands | Missing critical epidemiological data for calibration, no contact matrices available |
| Samoa | No case notification or incidence data for children |
| Somalia | No contact matrices available |
| St. Lucia | No case notification or incidence data for children |
| Tuvalu | No contact matrices available |
| West Bank and Gaza | Missing critical epidemiological data for calibration |

### 5.3 Countries incorporating the HIV structure

A separate stratum was included to dynamically model the tuberculosis-HIV co-epidemic if the proportion of tuberculosis cases among people living with HIV (PLHIV) was greater than or equal to 15%, and if the HIV prevalence in the country was greater than 1%, resulting in 23 of the 115 worth running countries incorporating the HIV structure (Table S5.3).

**Table S5.3** Countries incorporating the HIV structure with their corresponding HIV prevalence and proportion of tuberculosis cases among PLHIV.

| Country | HIV Prevalence (%) | Proportion of tuberculosis cases among PLHIV (%) |
|---|---|---|
| Botswana | 16.5 | 48.6 |
| Central African Republic | 2.1 | 25.4 |
| Côte d'Ivoire | 1.7 | 17.5 |
| Cameroon | 2.0 | 26.8 |
| Gabon | 2.3 | 32.8 |
| Ghana | 1.1 | 20.8 |
| The Gambia | 1.2 | 17.7 |
| Guinea-Bissau | 2.1 | 31.3 |
| Equatorial Guinea | 4.8 | 26.5 |
| Guyana | 1.1 | 19.0 |
| Kenya | 2.9 | 26.2 |
| Lesotho | 16.0 | 61.6 |
| Mozambique | 7.2 | 33.8 |
| Malawi | 5.9 | 46.6 |
| Namibia | 8.4 | 32.5 |
| Rwanda | 1.8 | 21.1 |
| Eswatini | 17.4 | 60.1 |
| Togo | 1.5 | 16.2 |
| Tanzania | 2.9 | 23.6 |
| Uganda | 3.4 | 39.0 |
| South Africa | 12.8 | 58.0 |
| Zambia | 6.7 | 46.2 |
| Zimbabwe | 9.6 | 59.8 |

### 6. No-New-Vaccine baselines

#### 6.1 *Status Quo No-New-Vaccine* baseline

The primary no-new-vaccine simulated was the "*Status Quo No-New-Vaccine*" baseline, which assumed non-vaccine tuberculosis interventions continue at current levels into the future. As reported country-level data includes the high coverage levels of neonatal BCG vaccination, this was not explicitly modelled. We assumed that BCG vaccination would not be discontinued over the model time horizon.

All countries were fitted to nine calibration targets in 2019:

- The tuberculosis incidence rate per 100,000 population (all ages, ages 0-14, and ages 15+)
- The tuberculosis case notification rate per 100,000 population (all ages, ages 0-14, and ages 15+)
- The tuberculosis mortality rate per 100,000 population (all ages)
- The ratio of the prevalence of subclinical tuberculosis with the prevalence of active tuberculosis (subclinical + clinical tuberculosis)
- The ratio of the prevalence of active tuberculosis in the low access-to-care class relative to the high access-to-care

In addition to the targets above, countries incorporating the HIV structure were fitted to four HIV specific calibration targets for all ages in 2019:

- HIV prevalence
- ART coverage
- Tuberculosis incidence rate in PLHIV per 100,000 population
- Tuberculosis mortality rate in PLHIV per 100,000 population

#### 6.2 *2025 End TB No-New-Vaccine* baseline

To provide conservative estimates on absolute vaccine impact, we simulated an alternative *No-New-Vaccine* baseline assuming scale-up between 2019 and 2025 in order to meet the End TB incidence target in 2025 of a reduction in 50% of the tuberculosis incidence rate in 2015 by 2025 for all ages.[51] To implement this, an additional parameter, $p_F$ (equal to 1 up to and including 2019 and sampled between 0 and 1 afterwards), was included in the model as a contact rate multiplier within the force of infection equation, and as a multiplier on the progression to disease flows. Using the fully fitted parameter sets for each country from the *Status Quo No-New-Vaccine baseline*, we then varied $p_F$ during calibration to hit the country-specific target of a reduction in 50% of the tuberculosis incidence rate in 2015 in 2025.

### 7. Vaccination

#### 7.1 Vaccine profile

The vaccine profile for an adult/adolescent vaccine and infant vaccine were based on the WHO Preferred Product Characteristics for New Tuberculosis vaccines,[52] and are outlined in Table S7.1 below.

**Table S7.1** WHO Preferred Product Characteristics for New Tuberculosis Vaccines

| Vaccine | Host infection status at time of vaccination required for efficacy | Effect type | Vaccine efficacy | Duration of protection |
|---|---|---|---|---|
| Adolescent / Adult | Pre- and post-infection | Prevention of disease | 50% | Lifelong |
| | | | | 10 years |
| Infant | Pre-infection | Prevention of disease | 80% | Lifelong |
| | | | | 10 years |

Vaccine efficacy was assumed to be the same in both PLHIV and HIV-naïve recipients in countries incorporating the HIV structure, and in both younger age groups and older adults. The vaccine was assumed to have the same impact on preventing drug-susceptible and drug-resistant tuberculosis as specified in the WHO PPCs,[52] and as we were modelling a prevention of disease vaccine, there was no direct impact on *Mtb* transmission or the force of infection.

#### 7.2 Vaccine delivery scenarios

The infant vaccine was implemented in two scenarios, and, separately, the adolescent/adult vaccine was implemented in three scenarios. The *Basecase* and *Accelerated Scale-up* scenarios included routine neonatal vaccination for the infant vaccine (85% coverage), and routine vaccination of 9-year-olds (80% coverage) with a one-time vaccination campaign for ages ten and older (70% coverage) for the adolescent/adult vaccine. The *Routine Only* scenario (adolescent/adult vaccine only) was introduced through routine 9-year-old vaccination only (i.e., no campaign). Specifics of the infant and adolescent/adult vaccine scenarios are provided in Table S7.5.

##### 7.2.1 Country-specific introduction years

In the *Basecase* and *Routine Only* scenarios, vaccines were introduced in country-specific introduction years between 2028 and 2047. To calculate the specific year of introduction, countries were divided into two general categories: those procuring with support from Gavi, the Vaccine Alliance, and those self-procuring. Determination of country status was based on eligibility information posted on Gavi's website.[53] Countries transitioning from Gavi support are able to benefit from Gavi pricing and incremental financing for a period of 5-10 years. For countries that have already initiated the period of transition by 2019, this window will have largely ended by the time of tuberculosis vaccine availability through Gavi. As such, these countries were categorised as self-procuring countries. Countries that have not yet commenced transition, including India and Nigeria, were categorised as Gavi supported countries, given the long grace period post-commencement of transition. For more information, please see Gavi, https://www.gavi.org/types-support/sustainability/transition (retrieved December 1, 2020).

Through a consultative process with experts from WHO, Gavi, PATH, PDVAC, CHAI, and industry partners, factors influencing likelihood of being an early or late adopter were identified for both Gavi and self-procuring countries. Identified factors include disease burden, immunization capacity, and early adopter status. Country-specific registration timelines and commercial prioritization were also deemed important determinants of introduction timing for self-procuring countries.

*Additional factors for Gavi countries:* For countries procuring through Gavi, timelines for introduction are also influenced by Gavi processes. Prior to offering a new vaccine, Gavi requires that products be licensed, included in Gavi's Vaccine Investment Strategy, reviewed by SAGE, recommended in a WHO position paper, WHO prequalified, and approved for procurement by Gavi (Table S7.2). In addition, time for country application processing, contracting, and delivery must be factored. Through consultations, it was determined that a baseline time of roughly two years post licensure would be needed for Gavi processes prior to first country introduction, assuming several steps advance in parallel.

**Table S7.2** Timelines for Gavi processes post licensure

| | Cumulative additional time (years) | | |
|---|---|---|---|
| **Activities post licensure** | **Low End** | **High End** | **Average** |
| WHO PQ | 0.25 | 1.00 | 0.63 |
| SAGE Policy Review & WHO Position Paper | 0.25 | 0.50 | 0.38 |
| Gavi Decision | 0.25 | 0.50 | 0.38 |
| National review & Country applications | 0.25 | 0.75 | 0.50 |
| Contracting & delivery | 0.25 | 0.50 | 0.38 |
| **Years** | **1.25** | **3.25** | **2.25** |

*Weight of criteria, indicators, and scoring*: Differential weight was assigned to criteria based on their relative impact on the order of country adoption. This weight varied for self-procuring and Gavi countries (Table S7.3).

**Table S7.3** Weight of criteria influencing order of country adoption

| Criteria | Self-procuring countries | Gavi countries |
|---|---|---|
| Disease burden | 30% | 45% |
| Immunization capacity | 15% | 30% |
| Early adopter/leader | 15% | 25% |
| Lack of regulatory barriers | 15% | NA |
| Commercial prioritization | 25% | NA |

The following indicators were used to measure each of the variables identified in Table S7.3.

**Table S7.4** Indicators of criteria influencing order of country adoption

| Criteria | Indicator |
|---|---|
| **Disease burden** | Tuberculosis incidence |
| **Immunization capacity** | % receiving 3 doses DPT3 among infants 1 years of age (The percent of infants receiving 3 doses DPT3 is commonly used as a proxy for assessing immunization infrastructure) |
| **Lack of regulatory barriers** | Signatories to WHO PQ or SRA collaborative registration scheme<br>Lack of requirements for additional local clinical trial data |
| **Early adopter/leader** | Time to policy adoption of universal Xpert MTB/RIF screening for presumed tuberculosis cases<br>Time to adoption of HPV |
| **Commercial prioritization** | |
| *Ability to finance vaccines* | GDP per capita |
| *Political will to address tuberculosis* | Spending per tuberculosis case |
| *Market potential* | Population |

To standardise across these varied metrics, a point value ranging from 1–5 per criteria was assigned, with a score of 1 correlating with an earlier adopter and score of 5 correlating with a later adopter.

*Continuous variable*s such as disease burden or population were divided into quintiles. Those in the highest quintile were assigned a score of 1, those in the second highest quintile received a score of 2, and so forth. *Categorical variables* such as registration or early adopter status were scored based on whether countries met fixed criteria. For instance, countries that are signatories of WHO PQ or SRA collaborative registration schemes were assigned a score of 1. Those that are not signatories and have requirements for additional clinical trial data in local populations received a score of 5.

Scores were then weighted as reflected in Table S7.3 and aggregated into a composite score to determine countries' relative position in the queue of introductions. Composite scores for each of the 135 countries are provided in the SupplementaryMaterial_CountryTimelines.xls.

Assumptions for the pace of introduction—i.e., how many countries per year would introduce the product and what the scale up curve might look like— was informed with data from pneumococcal vaccine (PCV) scale-up.[54] The percent of countries adopting each year (year 1 to year 12) for PCV was calculated. These annual percentages were then applied to tuberculosis vaccine scale up (based on a total n=135 countries: 78 self-procuring countries and 57 Gavi countries). The first year of tuberculosis vaccine scale up was estimated to be 2028, with Gavi countries following a similar scale up trajectory but delayed by two years due to required Gavi lead time for processing new vaccines (Table S7.2). Because PCV data is only available for 12 years, data was extrapolated for years 13 to 20 of tuberculosis vaccine roll out at a steady state. Country introduction timelines were adjusted—where applicable—to group countries with the same composite score in the same year of adoption. The cumulative number of countries introducing the vaccine by year is shown in Figure S7.1, and the country-specific introduction year for each country is in Table S8.1.

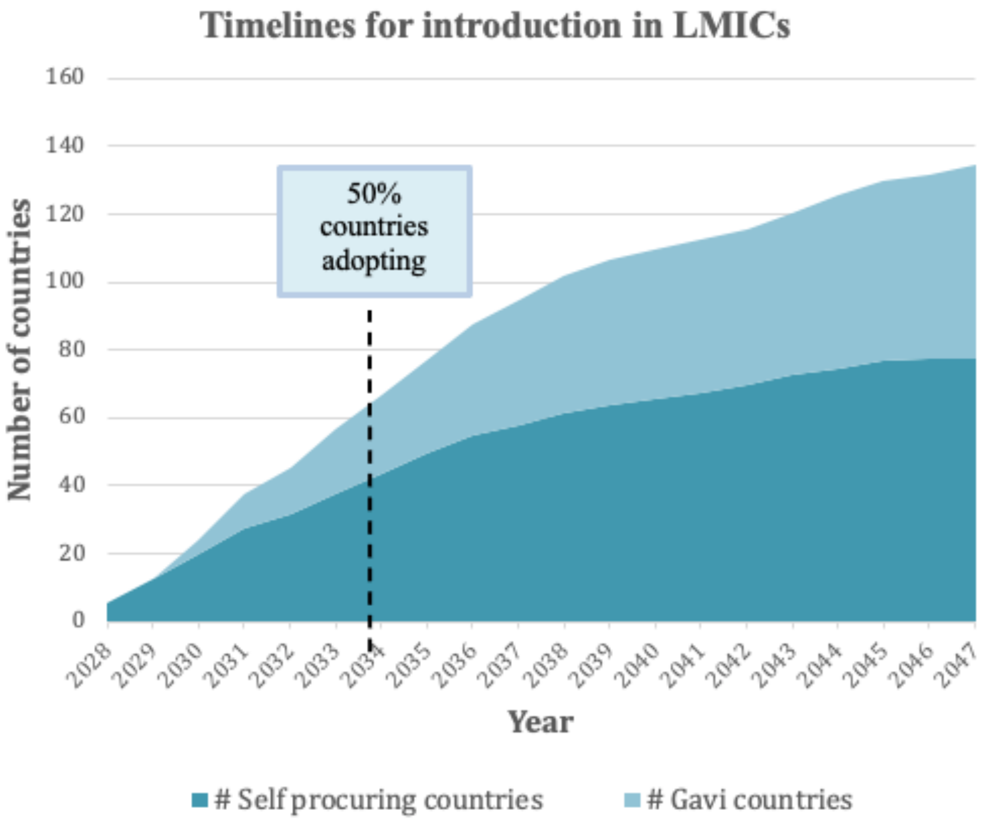


**Figure S7.1** Assumed cumulative number of countries introducing a novel vaccine per year

#### 7.2.2 Vaccine coverage targets

For each vaccine implementation scenario, low, medium, and high coverage targets for 5 years post-introduction were evaluated. The medium coverage target for the routine infant vaccination was 85%, based on the 2019 DTP3 (diphtheria, tetanus toxoid, and pertussis) average coverage level according to the WHO and UNICEF estimates of national immunisation coverage, with 10% uncertainty (low coverage = 75%, high coverage = 95%).[54] Routine adolescent vaccination assumed a medium coverage target of 80% aligning with HPV coverage in South Africa combined with aggregated secondary school enrolment in China and India as assumed in Harris 2020,[55] also with 10% uncertainty targets (low coverage = 70%, high coverage = 90%). The medium coverage target for the adolescent/adult campaign was 70% aligning with the lower bound of the MenAfriVac campaigns in sub-Saharan Africa as assumed in Harris 2020,[55] with a wider uncertainty of 20% (low coverage = 50%, high coverage = 90%). In the *Accelerated Scale-up* implementation, the 5-year coverage targets are achieved instantly in year 1, while in the *Basecase* and *Routine Only* implementations, the scale-up to coverage occurs linearly over 5 years.

**Table S7.5** Vaccine scenarios for the infant and adolescent/adult vaccines

| Characteristics | Infant Vaccine Scenarios | | Adolescent/Adult Vaccine Scenarios | | |
|---|---|---|---|---|---|
| | ***Basecase*** | ***Accelerated Scale-up*** | ***Basecase*** | ***Accelerated Scale-up*** | ***Routine Only*** |
| **Ages Targeted** | *Neonatal:* Routine | *Neonatal:* Routine | *Age 9:* Routine<br><br>*Ages 10+:* One-time vaccination campaign over 5 years | *Age 9:* Routine<br><br>*Ages 10+:* One-time vaccination campaign in 2025 | *Age 9:* Routine |
| **Introduction Year** | Country-specific | 2025 | Country-specific | 2025 | Country-specific |
| **Vaccine Rollout Trend** | 5-year linear scale-up to coverage | Instant scale-up to coverage | 5-year linear scale-up to coverage | Instant scale-up to coverage | 5-year linear scale-up to coverage |
| **Target Coverage *(Low/Med/High)*** | 75% / 85% / 95% | | Age 9: 70% / 80% / 90%<br>Ages 10+: 50% / 70% / 90% | | |

### 7.3 Vaccine implementation

#### 7.3.1 Vaccine structure

To simplify accounting for the number of vaccinees and vaccinations in the model, we included vaccines through an additional "vaccine structure" with three compartments (Figure S7.2) with the influences of vaccines on tuberculosis natural history parameters occurring separately in the natural history structure (Figure S7.3).

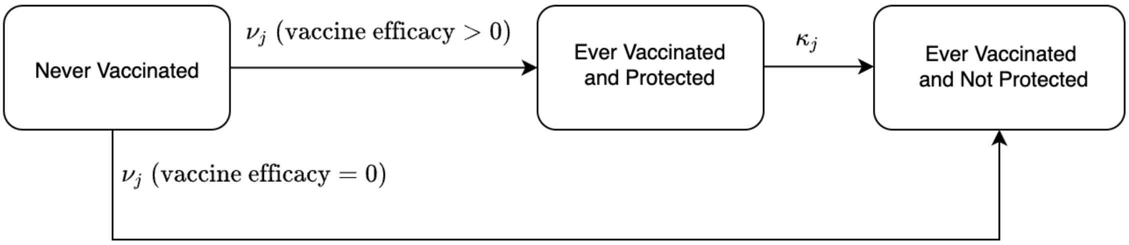


**Figure S7.2** Vaccine structure

Before vaccination, all individuals in the model begin in the *Never Vaccinated* compartment, with no vaccine protection. Upon vaccination, individuals either transition to the *Ever Vaccinated and Protected* compartment (with vaccine protection) or *Ever Vaccinated and Not Protected* compartment (with no vaccine protection), depending on vaccine specific host infection status at time of vaccination required for vaccine take and their infection status at the time of vaccination, summarised in Table S7.6.

In this work, we modelled the infant vaccine as a pre-infection (PRI) vaccine, meaning the individual must be uninfected at the time of vaccination for the vaccine to be efficacious ("take"). We modelled the adolescent/adult vaccine as a pre- and post-infection (PPI) vaccine, which means that it will be efficacious in any infection status at time of vaccination aside from active disease. We assumed that the effect of disease on the immune response is likely to be substantially larger than any additional benefit from the vaccine, and therefore would not be efficacious in those compartments. For example, the Phase 2b M72/AS01$_E$ trial saw a small number of cases in each arm within the first 6 months after ruling out those who were XPERT positive on the day they were tested. Assuming those cases were individuals who were subclinical but not XPERT positive on the day they were tested, the vaccine had no impact on their disease progression.[56]

The arrow directly from *Never Vaccinated* to *Ever Vaccinated and Not Protected* was included to account for individuals who may be accidentally administered a vaccine which would not be efficacious (i.e., would not "take" and therefore vaccine efficacy is zero) given their infection status at the time of vaccination. As individuals with subclinical disease present with no symptoms, it is possible that they may be accidentally vaccinated, as seen in the Phase 2b M72/AS01E trial. Similarly, with a PRI vaccine, if no pre-vaccination testing is available, it is possible that individuals who are not uninfected may be vaccinated. By including the flow directly to *Ever Vaccinated and Not Protected*, we could easily identify and track these individuals, and ensure they received no protection from the vaccine in the model.

**Table S7.6** Transitions within the vaccine structure following vaccination based on natural history state and host infection status at time of vaccination required for vaccine take

| | **Host infection status at the time of vaccination required for vaccine to take** | |
|---|---|---|
| **Natural History State (Infection Status) at time of vaccination** | **Pre-infection vaccine** (i.e., the infant vaccine) | **Pre- and post-infection vaccine** (i.e., the adolescent/adult vaccine) |
| Uninfected – Naïve | *Ever Vaccinated and Protected* | *Ever Vaccinated and Protected* |
| Uninfected – Cleared | *Ever Vaccinated and Not Protected* | *Ever Vaccinated and Protected* |
| Infection – Fast | *Ever Vaccinated and Not Protected* | *Ever Vaccinated and Protected* |
| Infection – Slow | *Ever Vaccinated and Not Protected* | *Ever Vaccinated and Protected* |
| Subclinical Disease | *Ever Vaccinated and Not Protected* | *Ever Vaccinated and Not Protected* |
| Clinical Disease | *NA* | *NA* |
| On-treatment | *NA* | *NA* |
| Resolved | *Ever Vaccinated and Not Protected* | *Ever Vaccinated and Protected* |

Waning, or loss of vaccine protection, moved individuals from the *Ever Vaccinated and Protected* compartment to the *Ever Vaccinated and Not Protected* compartment. We assumed duration of protection was 10 years on average, in addition to a sensitivity analysis with lifelong duration of protection. The shape of waning immunity was modelled as an exponential distribution, based on similar shapes for waning vaccine immunity of BCG[57] and other vaccines.[58,59]

#### 7.3.1 Vaccine implementation in the tuberculosis natural history model

Vaccines are incorporated in the tuberculosis natural history structure as indicated with the orange boxes in Figure S7.3 by reducing the rate of progression to disease parameters into the subclinical disease compartment from the infection-fast, infection-slow, and resolved compartments by (1-$p_V$), where $p_V$ is the vaccine efficacy. Vaccine efficacy was modelled as "degree", also known as "leaky". Degree vaccines assume that everyone who has been vaccinated receives some protection from the vaccine equivalent to the value of the vaccine efficacy.

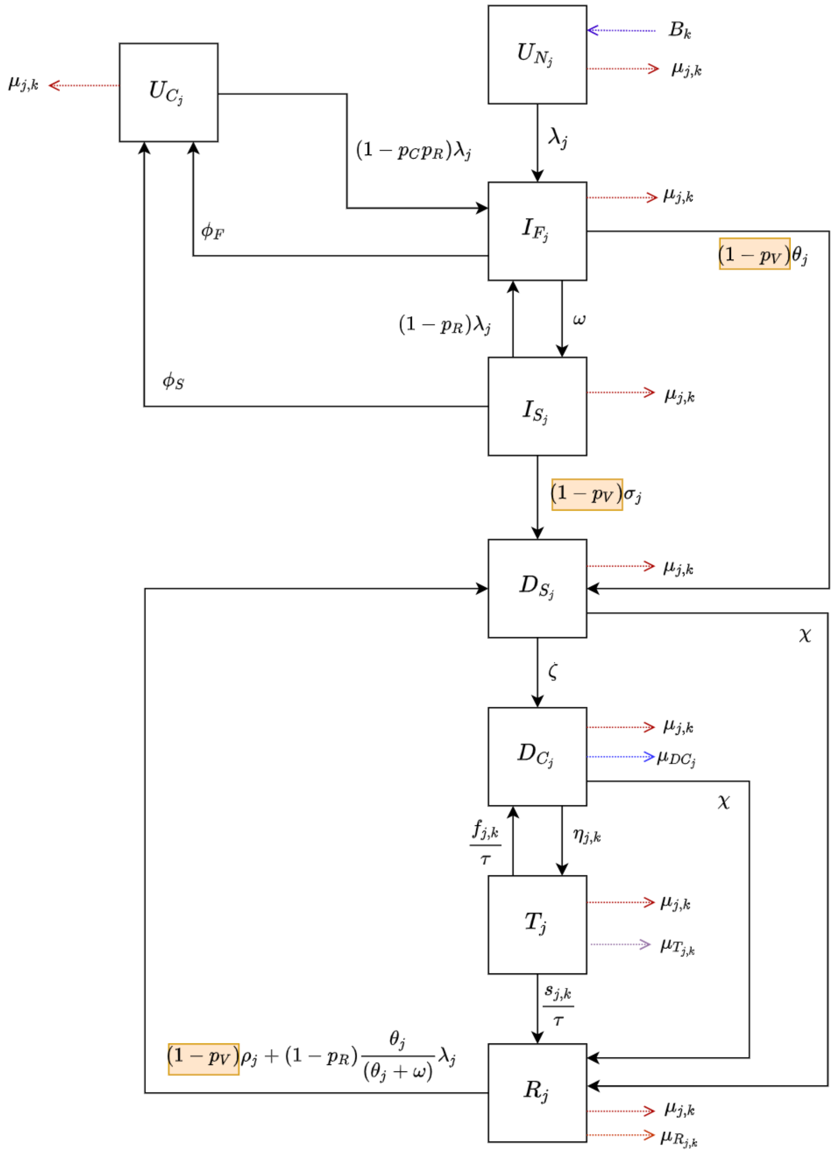


**Figure S7.3** Tuberculosis natural history model incorporating vaccination

*Abbreviations: $D_C$ = Clinical Disease; $D_S$ = Subclinical Disease; $I_F$ = Infection-Fast; $I_S$ = Infection-Slow; R = Resolved; T = On-Treatment; $U_C$ = Uninfected-Cleared; $U_N$ = Uninfected-Naive.*

### 8. Model outcomes

#### 8.1 Epidemiological impact measures

The following measures were calculated for each vaccine scenario as the median and 95% uncertainty range

- Percent incidence rate reduction in 2050 for each vaccine scenario compared to the *No-New-Vaccine* baseline
- Incidence rate per 100,000 population in 2035 for each vaccine scenario
- Percent mortality rate reduction in 2050 for each vaccine scenario compared to the *No-New-Vaccine* baseline
- Cumulative cases averted for each vaccine scenario between vaccine introduction (either 2025 or country-specific years) and 2050 compared to the *No-New-Vaccine* baseline
- Cumulative deaths averted for each vaccine scenario between vaccine introduction (either 2025 or country-specific years) and 2050 compared to the *No-New-Vaccine* baseline
- Cumulative treatments averted for each vaccine scenario between vaccine introduction (either 2025 or country-specific years) and 2050 compared to the *No-New-Vaccine* baseline

#### 8.2 Groupings for reporting model outcomes

The epidemiological impact measures were calculated and reported for the calibrated countries by WHO region, World Bank Income Group, for tuberculosis burden, and overall. Countries are divided into the six WHO regions, [African region (AFR), region of the Americas (AMR), Eastern-Mediterranean region (EMR), South-East Asian region (SEAR), and Western-Pacific region (WPR)], three income groups based on the 2021 World Bank Income Groups for low- and middle-income countries [low-income countries (LIC), lower-middle-income countries (LMIC) or upper-middle-income countries (UMIC)], and by whether they were or were not included on the WHO high TB burden list (*High TB Burden* vs *Other* respectively). Groups for each of the LMICs are in Table S8.1.

**Table S8.1** Country-specific introduction year, WHO region, World Bank income group 2021, and WHO TB burden level for LMICs

| Country | Gavi/Self Procuring | Introduction Year | WHO Region | World Bank Income Group 2021 | WHO High TB Burden |
|---|---|---|---|---|---|
| Afghanistan | Gavi | 2031 | EMR | LIC | Other |
| Albania | Self-Procuring | 2035 | EUR | UMIC | Other |
| Algeria | Self-Procuring | 2032 | AFR | LMIC | Other |
| American Samoa | Self-Procuring | 2044 | WPR | UMIC | Other |
| Angola | Self-Procuring | 2032 | AFR | LMIC | High TB Burden |
| Argentina | Self-Procuring | 2031 | AMR | UMIC | Other |
| Armenia | Self-Procuring | 2033 | EUR | UMIC | Other |
| Azerbaijan | Self-Procuring | 2028 | EUR | UMIC | Other |
| Bangladesh | Gavi | 2035 | SEAR | LMIC | High TB Burden |
| Belarus | Self-Procuring | 2028 | EUR | UMIC | Other |
| Belize | Self-Procuring | 2034 | AMR | UMIC | Other |
| Benin | Gavi | 2037 | AFR | LMIC | Other |
| Bhutan | Self-Procuring | 2034 | SEAR | LMIC | Other |
| Bolivia | Self-Procuring | 2037 | AMR | LMIC | Other |
| Bosnia and Herzegovina | Self-Procuring | 2039 | EUR | UMIC | Other |
| Botswana | Self-Procuring | 2028 | AFR | UMIC | Other |
| Brazil | Self-Procuring | 2030 | AMR | UMIC | High TB Burden |
| Bulgaria | Self-Procuring | 2029 | EUR | UMIC | Other |
| Burkina Faso | Gavi | 2039 | AFR | LIC | Other |
| Burundi | Gavi | 2044 | AFR | LIC | Other |
| Cabo Verde | Self-Procuring | 2043 | AFR | LMIC | Other |
| Cambodia | Gavi | 2036 | WPR | LMIC | Other |
| Cameroon | Gavi | 2031 | AFR | LMIC | Other |
| Central African Republic | Gavi | 2033 | AFR | LIC | High TB Burden |
| Chad | Gavi | 2033 | AFR | LIC | Other |
| China | Self-Procuring | 2029 | WPR | UMIC | High TB Burden |
| Colombia | Self-Procuring | 2030 | AMR | UMIC | Other |
| Comoros | Gavi | 2046 | AFR | LMIC | Other |
| Congo | Gavi | 2035 | AFR | LMIC | High TB Burden |
| Costa Rica | Self-Procuring | 2033 | AMR | UMIC | Other |
| Côte d'Ivoire | Gavi | 2034 | AFR | LMIC | Other |
| Cuba | Self-Procuring | 2035 | AMR | UMIC | Other |
| Democratic People’s Republic of Korea | Gavi | 2035 | SEAR | LIC | High TB Burden |

| | | | | | |
|---|---|---|---|---|---|
| Democratic Republic of the Congo | Gavi | 2030 | AFR | LIC | High TB Burden |
| Djibouti | Gavi | 2043 | EMR | LMIC | Other |
| Dominica | Self-Procuring | 2031 | AMR | UMIC | Other |
| Dominican Republic | Self-Procuring | 2031 | AMR | UMIC | Other |
| Ecuador | Self-Procuring | 2033 | AMR | UMIC | Other |
| Egypt | Self-Procuring | 2033 | EMR | LMIC | Other |
| El Salvador | Self-Procuring | 2039 | AMR | LMIC | Other |
| Equatorial Guinea | Self-Procuring | 2042 | AFR | UMIC | Other |
| Eritrea | Gavi | 2047 | AFR | LIC | Other |
| Ethiopia | Gavi | 2030 | AFR | LIC | High TB Burden |
| Fiji | Self-Procuring | 2031 | WPR | UMIC | Other |
| Gabon | Self-Procuring | 2038 | AFR | UMIC | High TB Burden |
| Gambia | Gavi | 2039 | AFR | LIC | Other |
| Georgia | Self-Procuring | 2029 | EUR | UMIC | Other |
| Ghana | Gavi | 2040 | AFR | LMIC | Other |
| Grenada | Self-Procuring | 2035 | AMR | UMIC | Other |
| Guatemala | Self-Procuring | 2036 | AMR | UMIC | Other |
| Guinea | Gavi | 2033 | AFR | LIC | Other |
| Guinea-Bissau | Gavi | 2043 | AFR | LIC | Other |
| Guyana | Self-Procuring | 2030 | AMR | UMIC | Other |
| Haiti | Gavi | 2033 | AMR | LIC | Other |
| Honduras | Self-Procuring | 2037 | AMR | LMIC | Other |
| India | Gavi | 2033 | SEAR | LMIC | High TB Burden |
| Indonesia | Self-Procuring | 2034 | SEAR | LMIC | High TB Burden |
| Iran | Self-Procuring | 2031 | EMR | LMIC | Other |
| Iraq | Self-Procuring | 2033 | EMR | UMIC | Other |
| Jamaica | Self-Procuring | 2036 | AMR | UMIC | Other |
| Jordan | Self-Procuring | 2037 | EMR | UMIC | Other |
| Kazakhstan | Self-Procuring | 2028 | EUR | UMIC | Other |
| Kenya | Gavi | 2032 | AFR | LMIC | High TB Burden |
| Kiribati | Self-Procuring | 2041 | WPR | LMIC | Other |
| Kosovo | Self-Procuring | 2045 | EUR | UMIC | Other |
| Kyrgyz Republic | Gavi | 2044 | EUR | LMIC | Other |
| Lao People's Democratic Republic | Gavi | 2035 | WPR | LMIC | Other |
| Lebanon | Self-Procuring | 2038 | EMR | UMIC | Other |
| Lesotho | Gavi | 2039 | AFR | LMIC | High TB Burden |
| Liberia | Gavi | 2037 | AFR | LIC | High TB Burden |
| Libya | Self-Procuring | 2035 | EMR | UMIC | Other |

| | | | | | |
|---|---|---|---|---|---|
| Madagascar | Gavi | 2031 | AFR | LIC | Other |
| Malawi | Gavi | 2038 | AFR | LIC | Other |
| Malaysia | Self-Procuring | 2028 | WPR | UMIC | Other |
| Maldives | Self-Procuring | 2034 | SEAR | UMIC | Other |
| Mali | Gavi | 2037 | AFR | LIC | Other |
| Marshall Islands | Self-Procuring | 2041 | WPR | UMIC | Other |
| Mauritania | Gavi | 2042 | AFR | LMIC | Other |
| Mexico | Self-Procuring | 2029 | AMR | UMIC | Other |
| Micronesia | Self-Procuring | 2045 | WPR | LMIC | Other |
| Mongolia | Self-Procuring | 2032 | WPR | LMIC | High TB Burden |
| Montenegro | Self-Procuring | 2044 | EUR | UMIC | Other |
| Morocco | Self-Procuring | 2029 | EMR | LMIC | Other |
| Mozambique | Gavi | 2032 | AFR | LIC | High TB Burden |
| Myanmar | Gavi | 2031 | SEAR | LMIC | High TB Burden |
| Namibia | Self-Procuring | 2030 | AFR | UMIC | High TB Burden |
| Nepal | Gavi | 2036 | SEAR | LMIC | Other |
| Nicaragua | Gavi | 2047 | AMR | LMIC | Other |
| Niger | Gavi | 2036 | AFR | LIC | Other |
| Nigeria | Gavi | 2030 | AFR | LMIC | High TB Burden |
| North Macedonia | Self-Procuring | 2038 | EUR | UMIC | Other |
| Pakistan | Gavi | 2031 | EMR | LMIC | High TB Burden |
| Papua New Guinea | Gavi | 2032 | WPR | LMIC | High TB Burden |
| Paraguay | Self-Procuring | 2035 | AMR | UMIC | Other |
| Peru | Self-Procuring | 2029 | AMR | UMIC | Other |
| Philippines | Self-Procuring | 2030 | WPR | LMIC | High TB Burden |
| Republic of Moldova | Self-Procuring | 2034 | EUR | UMIC | Other |
| Russian Federation | Self-Procuring | 2030 | EUR | UMIC | Other |
| Rwanda | Gavi | 2045 | AFR | LIC | Other |
| Samoa | Self-Procuring | 2046 | WPR | UMIC | Other |
| Sao Tome and Principe | Gavi | 2044 | AFR | LMIC | Other |
| Senegal | Gavi | 2038 | AFR | LMIC | Other |
| Serbia | Self-Procuring | 2036 | EUR | UMIC | Other |
| Sierra Leone | Gavi | 2037 | AFR | LIC | High TB Burden |
| Solomon Islands | Gavi | 2047 | WPR | LMIC | Other |
| Somalia | Gavi | 2030 | EMR | LIC | Other |
| South Africa | Self-Procuring | 2029 | AFR | UMIC | High TB Burden |
| South Sudan | Gavi | 2034 | AFR | LIC | Other |
| Sri Lanka | Self-Procuring | 2028 | SEAR | LMIC | Other |

| | | | | | |
|---|---|---|---|---|---|
| St. Lucia | Self-Procuring | 2031 | AMR | UMIC | Other |
| St. Vincent and the Grenadines | Self-Procuring | 2032 | AMR | UMIC | Other |
| Sudan | Gavi | 2036 | EMR | LIC | Other |
| Suriname | Self-Procuring | 2040 | AMR | UMIC | Other |
| Swaziland | Self-Procuring | 2036 | AFR | LMIC | Other |
| Syrian Arab Republic | Gavi | 2036 | EMR | LIC | Other |
| Tajikistan | Gavi | 2045 | EUR | LMIC | Other |
| Thailand | Self-Procuring | 2031 | SEAR | UMIC | High TB Burden |
| Timor-Leste | Self-Procuring | 2031 | SEAR | LMIC | Other |
| Togo | Gavi | 2041 | AFR | LIC | Other |
| Tonga | Self-Procuring | 2040 | WPR | UMIC | Other |
| Tunisia | Self-Procuring | 2036 | EMR | LMIC | Other |
| Turkey | Self-Procuring | 2030 | EUR | UMIC | Other |
| Turkmenistan | Self-Procuring | 2034 | EUR | UMIC | Other |
| Tuvalu | Self-Procuring | 2043 | WPR | UMIC | Other |
| Uganda | Gavi | 2034 | AFR | LIC | High TB Burden |
| Ukraine | Self-Procuring | 2033 | EUR | LMIC | Other |
| United Republic of Tanzania | Gavi | 2031 | AFR | LMIC | High TB Burden |
| Uzbekistan | Gavi | 2038 | EUR | LMIC | Other |
| Vanuatu | Self-Procuring | 2042 | WPR | LMIC | Other |
| Venezuela | Self-Procuring | 2035 | AMR | UMIC | Other |
| Vietnam | Self-Procuring | 2038 | WPR | LMIC | High TB Burden |
| West Bank and Gaza | Self-Procuring | 2043 | EMR | LMIC | Other |
| Yemen | Gavi | 2036 | EMR | LIC | Other |
| Zambia | Gavi | 2034 | AFR | LIC | High TB Burden |
| Zimbabwe | Gavi | 2032 | AFR | LMIC | Other |

*Abbreviations: AFR = WHO African Region, AMR = WHO Region of the Americas, EMR = WHO Eastern Mediterranean Region, EUR = WHO European Region, LIC = low-income countries, LMIC = lower-middle income countries, SEAR = WHO South-East Asian Region, UMIC = upper-middle income countries, WPR = WHO Western Pacific Region*

### 8.2 Calculating uncertainty

To appropriately represent the global uncertainty and remove inter-country variability in parameters that are likely to be the same across countries when generating impact estimates (e.g., those governing the underlying biology of *Mtb*), we used the following process:

1. We obtained 1000 fitted parameter sets for each country by thinning the total number of fitted parameter sets per country to 1000.

2. Within each country, the 1000 parameter sets were ordered and ranked from smallest to largest by 2019 tuberculosis incidence rate.

3. The parameter sets for all countries were then pairwise grouped on their rank value. For example, the rank 1 parameter sets were grouped together for all countries, the rank 2 parameter sets were grouped together for all countries, etc.

4. Within each pairwise rank group, we calculated the measure of interest by combining all information. For example, to calculate the incidence rate, we summed the number of cases from all countries with rank 1 and divided by the sum of the population for all countries with rank 1. This was continued for all ranks until there were 1000 estimates of the measure of interest.

5. We combined the 1000 estimates for the measure of interest, generated the distribution and calculated all country and group-level estimates.

**SUPPLEMENTAL RESULTS:**

### 9. Model Calibration

#### 9.1 LMIC countries calibration

Of the 135 LMICs, there were 20 countries which were excluded from calibration due to missing crucial data for calibration (as described in Table S5.3). The 10 countries that did not calibrate out of the 115 worth running were: Algeria, Bosnia and Herzegovina, Cabo Verde, Guinea-Bissau, Guyana, Jamaica, North Macedonia, St. Vincent and the Grenadines, Tonga, and Turkmenistan.

#### 9.2 WHO Region and World Bank Income Group for Calibrated LMICs

**Table S9.1** WHO regions and World bank income groups for calibrated LMICs

| WHO Region | World Bank Income Level 2019 | | | *Total* |
|---|---|---|---|---|
| | LIC | LMIC | UMIC | |
| AFR | 19 | 15 | 5 | 39 |
| AMR | 0 | 4 | 13 | 17 |
| EMR | 4 | 5 | 3 | 12 |
| EUR | 0 | 4 | 12 | 16 |
| SEAR | 0 | 8 | 2 | 10 |
| WPR | 0 | 8 | 3 | 11 |
| ***Total*** | 23 | 44 | 38 | 105 |

*Abbreviations: AFR = WHO African Region, AMR = WHO Region of the Americas, EMR = WHO Eastern Mediterranean Region, EUR = WHO European Region, LIC = low-income countries, LMIC = lower-middle income countries, SEAR = WHO South-East Asian Region, UMIC = upper-middle income countries, WPR = WHO Western Pacific Region*

### 9.3 *Status Quo No-New-Vaccine* baseline calibration target values

Table S9.2 presents the common country-specific calibration targets for the 105 calibrated countries, and Table S9.3 highlights the HIV specific calibration targets for the 21 countries incorporating the HIV structure. Two additional calibration targets were assumed consistent across countries: the fraction of subclinical tuberculosis among active tuberculosis in 2019 [50.4% (36.1%–79.7%)],[1] and the risk ratio of active tuberculosis in the low-access-to-care group relative to high-access-to-care in 2019 [67.4 (57.5, 80.1)].[12–21]

**Table S9.2** Values for the seven country specific calibration targets for all calibrated countries in 2019. Point values represent the mean with 95% confidence intervals in brackets.

| Country | Tuberculosis Incidence Rate (per 100,000 population per year) | | | Tuberculosis Notification Rate (per 100,000 population per year) | | | Tuberculosis Mortality Rate (per 100,000 population per year) |
|---|---|---|---|---|---|---|---|
| | Ages 0–14 | Ages 15+ | Ages 0–99 | Ages 0–14 | Ages 15+ | Ages 0–99 | Ages 0–99 |
| Afghanistan | 92.8 (48.9–136.2) | 260.5 (137.1–383.8) | 189.3 (115.7–262.9) | 71.0 (56.8–85.2) | 187.2 (149.7–224.6) | 137.8 (110.3–165.4) | 26 (16–39) |
| Albania | 2.6 (2.2–3.0) | 19.3 (16.4–22.3) | 16.3 (13.9–18.7) | 2.2 (1.8–2.6) | 16.9 (13.5–20.2) | 14.3 (11.4–17.2) | 0.3 (0.2–0.5) |
| Angola | 107.8 (58.6–155) | 565.1 (312.0–818.2) | 351.9 (213.7–487.0) | 59.4 (47.5–71.3) | 384.3 (307.5–461.2) | 232.8 (186.3–279.4) | 62 (39–89) |
| Argentina | 9.1 (7.9–10.9) | 35.5 (29.6–41.5) | 29.0 (24.6–33.5) | 8.1 (6.5–9.8) | 31.2 (24.9–37.4) | 25.6 (20.4–30.7) | 1.6 (1.4–1.8) |
| Armenia | 7.7 (5.7–9.8) | 31.1 (23.0–39.3) | 26.4 (19.6–32.8) | 6.2 (4.9–7.4) | 24.9 (19.9–29.9) | 21.0 (16.8–25.2) | 0.6 (0.4–0.8) |
| Azerbaijan | 19.5 (14–24.6) | 72.8 (53.3–92.3) | 59.7 (44.8–74.6) | 7.6 (6.1–9.1) | 44.6 (35.7–53.6) | 48.0 (38.4–57.6) | 6.1 (5.6–6.6) |
| Bangladesh | 74.4 (49.6–96.9) | 276.4 (187.9–364.9) | 221.4 (156.4–285.8) | 27.8 (22.2–33.3) | 235.3 (188.3–282.4) | 178.8 (143.1–214.6) | 24 (15–35) |
| Belarus | 4.3 (3.2–5.3) | 34.4 (25.5–43.4) | 29.6 (22.2–36.0) | 0.5 (0.4–0.6) | 28 (22.4–33.6) | 23.3 (18.7–28.0) | 3.3 (3.1–3.5) |
| Benin | 11.4 (6.6–16.3) | 86.5 (49.9–124.6) | 55.1 (33.9–77.1) | 4.6 (3.7–5.6) | 59.2 (47.3–71.0) | 36.1 (28.9–43.4) | 11 (7.2–15) |
| Bhutan | 32.1 (23.8–40.9) | 210.6 (154.5–263.3) | 170.4 (123.2–209.7) | 14.5 (11.6–17.4) | 171.5 (137.2–205.8) | 131.7 (105.4–158.0) | 18 (12–26) |

| | | | | | | | |
|---|---|---|---|---|---|---|---|
| Bolivia | 25.8<br>(15.3–36.9) | 137.7<br>(82.6–200.3) | 104.2<br>(65.1–147.7) | 6.9<br>(5.6–8.3) | 90.3<br>(72.3–108.4) | 64.8<br>(51.8–77.8) | 11<br>(8.1–15) |
| Botswana | 74.6<br>(55.3–93.9) | 340.8<br>(249.0–432.5) | 251.8<br>(191.0–312.5) | 29.8<br>(23.9–35.8) | 187.2<br>(149.8–224.7) | 134.1<br>(107.3–160.9) | 78<br>(61–96) |
| Brazil | 7.0<br>(5.9–8.1) | 55.8<br>(47.4–64.2) | 45.5<br>(38.9–52.1) | 6.0<br>(4.8–7.3) | 48.5<br>(38.8–58.2) | 39.6<br>(31.7–47.5) | 3.2<br>(2.9–3.4) |
| Bulgaria | 8.8<br>(6.4–10.7) | 21.8<br>(16.6–28.5) | 20.0<br>(15.7–25.7) | 5.3<br>(4.2–6.3) | 20.7<br>(16.5–24.8) | 18.4<br>(14.7–22.1) | 1.6<br>(1.6–1.7) |
| Burkina Faso | 9.8<br>(5.6–14.3) | 77.4<br>(44.5–106.7) | 47.2<br>(28.5–64.0) | 1.5<br>(1.2–1.8) | 42.0<br>(33.6–50.4) | 24.2<br>(19.4–29) | 11<br>(7.2–16) |
| Burundi | 24.8<br>(13.8–34.4) | 174.7<br>(100.1–254.1) | 104.1<br>(65.0–147.4) | 6.2<br>(5.0–7.4) | 102.1<br>(81.7–122.5) | 58.6<br>(46.9–70.3) | 24<br>(15–35) |
| Cambodia | 68.3<br>(39.0–97.5) | 387.3<br>(220.1–545.8) | 285.1<br>(175.9–400.3) | 36.1<br>(28.9–43.3) | 247.0<br>(197.6–296.4) | 181.4<br>(145.1–217.7) | 20<br>(13–28) |
| Cameroon | 43.8<br>(24.6–62.0) | 274.9<br>(154.2–395.6) | 177.8<br>(108.2–247.3) | 11.4<br>(9.2–13.7) | 154.6<br>(123.7–185.6) | 94.0<br>(75.2–112.8) | 48<br>(34–65) |
| Central African Republic | 182.3<br>(100.8–263.9) | 826.7<br>(450.9–1202.5) | 547.9<br>(337.2–758.7) | 79.1<br>(63.3–94.9) | 391.2<br>(313.0–469.5) | 253.9<br>(203.1–304.6) | 158<br>(111–215) |
| Chad | 36.2<br>(20.1–52.2) | 235.8<br>(129.7–342) | 144.2<br>(87.8–194.4) | 14<br>(11.2–16.8) | 148.5<br>(118.8–178.2) | 85.5<br>(68.4–102.6) | 31<br>(21–42) |
| China | 14.9<br>(12.5–16.8) | 67.5<br>(57.4–77.6) | 58.1<br>(49.7–66.5) | 2.6<br>(2.1–3.1) | 61.2<br>(49.0–73.5) | 50.8<br>(40.6–61) | 2.3<br>(2.1–2.6) |
| Colombia | 7.2<br>(5.4–8.8) | 43.6<br>(33.4–53.9) | 35.8<br>(25.8–43.7) | 3.5<br>(2.8–4.2) | 35.7<br>(28.5–42.8) | 28.4<br>(22.7–34.1) | 3.4<br>(3–3.9) |
| Costa Rica | 1.7<br>(1.2–2.1) | 12.3<br>(9–15.6) | 10.1<br>(7.5–12.5) | 1.3<br>(1.1–1.6) | 9.8<br>(7.9–11.8) | 8.0<br>(6.4–9.6) | 0.8<br>(0.7–0.9) |
| Cote d'Ivoire | 30.8<br>(17.7–43.8) | 213.5<br>(120.1–306.9) | 136.1<br>(81.7–190.5) | 9.5<br>(7.6–11.4) | 134.7<br>(107.8–161.6) | 82.5<br>(66–99) | 30<br>(20–41) |
| Cuba | 0.4<br>(0.3–0.5) | 7.6<br>(6.4–8.7) | 6.4<br>(5.5–7.4) | 0.3<br>(0.2–0.4) | 6.6<br>(5.3–7.9) | 5.6<br>(4.5–6.7) | 0.4<br>(0.3–0.5) |
| Dominican Republic | 6.7<br>(5.0–8.4) | 55.4<br>(41.2–69.6) | 41.9<br>(31.7–52.1) | 2.7<br>(2.2–3.2) | 45.2<br>(36.2–54.3) | 33.4<br>(26.8–40.1) | 4<br>(2.4–6) |
| Ecuador | 8.7<br>(6.4–11.0) | 59.7<br>(43.8–75.6) | 45.5<br>(34.5–57.0) | 3.9<br>(3.1–4.7) | 49.1<br>(39.2–58.9) | 36.5<br>(29.2–43.8) | 4.6<br>(3.9–5.3) |
| Egypt | 2.6<br>(2.3–2.9) | 16.6<br>(14.2–18.1) | 12.0<br>(10.0–12.9) | 1.4<br>(1.1–1.7) | 11.4<br>(9.1–13.7) | 8<br>(6.4–9.6) | 0.5<br>(0.4–0.5) |

| | | | | | | | |
|---|---|---|---|---|---|---|---|
| El Salvador | 18.5 (13.3–23.7) | 72 (53–93.2) | 58.9 (43.4–72.8) | 7.8 (6.2–9.3) | 53.7 (43–64.5) | 46.6 (37.3–56) | 1.7 (1.4–2) |
| Equatorial Guinea | 45.9 (39.9–53.9) | 257.3 (222.3–292.4) | 184.4 (154.9–206.5) | 19.2 (15.3–23) | 169.0 (135.2–202.8) | 114.3 (91.4–137.2) | 39 (31–48) |
| Eritrea | 29.6 (7.6–51.6) | 127.3 (32.8–220.3) | 85.8 (31.5–143) | 15.8 (12.7–19) | 78.2 (62.6–93.9) | 52.3 (41.8–62.7) | 17 (8–29) |
| Ethiopia | 37.6 (24.3–53.1) | 207.9 (133.1–282.6) | 140.1 (94.6–184.7) | 24.4 (19.5–29.3) | 149.6 (119.7–179.5) | 99.1 (79.3–118.9) | 22 (14–30) |
| Fiji | 57.5 (38.4–76.7) | 69.9 (47.7–92.2) | 66.3 (49.4–83.2) | 47.2 (37.7–56.6) | 55.5 (44.4–66.6) | 53 (42.4–63.6) | 5.3 (4.9–5.7) |
| Gabon | 160.9 (92.8–235.2) | 732.7 (410.3–1025.8) | 506.3 (317.6–736.5) | 45.4 (36.3–54.5) | 368.7 (295–442.4) | 248.5 (198.8–298.2) | 110 (66–165) |
| Gambia | 37.6 (26.1–50.2) | 251.6 (175.3–327.8) | 157.6 (115.0–200.2) | 12.5 (10–15.1) | 189.8 (151.8–227.7) | 111.6 (89.2–133.9) | 27 (20–35) |
| Georgia | 20 (16.2–23.7) | 87.6 (72–103.2) | 75.1 (62.6–87.6) | 9 (7.2–10.8) | 65.6 (52.5–78.7) | 54.3 (43.4–65.1) | 4.2 (3.8–4.6) |
| Ghana | 55.4 (16.7–96.8) | 194.2 (57.7–335.9) | 144.7 (55.9–230.1) | 7.2 (5.8–8.7) | 72.8 (58.2–87.3) | 48.3 (38.6–58) | 50 (29–77) |
| Guatemala | 10.9 (7.9–14.1) | 34.4 (24.1–43.9) | 26.2 (19.9–33) | 7.4 (5.9–8.9) | 28 (22.4–33.6) | 21.1 (16.9–25.4) | 2.4 (2.1–2.6) |
| Guinea | 37.8 (21.6–54.1) | 276.9 (166.2–401.5) | 172.3 (109.6–242.7) | 20.9 (16.7–25) | 210.9 (168.7–253.1) | 128.3 (102.7–154) | 29 (20–39) |
| Honduras | 4.3 (3.2–5.3) | 43.2 (31.3–55.1) | 30.8 (23.6–39) | 1.6 (1.3–1.9) | 35.4 (28.3–42.5) | 24.9 (19.9–29.8) | 5 (4.2–5.8) |
| India | 91.6 (55.8–127.6) | 229.4 (139.6–320.1) | 193.2 (125.9–259.8) | 40 (32–48) | 201.1 (160.9–241.4) | 158.2 (126.6–189.9) | 33 (30–35) |
| Indonesia | 200.2 (179–221.3) | 352.1 (314.5–389.6) | 312.2 (284.2–340.3) | 98.8 (79–118.6) | 245.5 (196.4–294.6) | 207.7 (166.1–249.2) | 36 (33–38) |
| Iran | 3.0 (2.3–3.9) | 16 (11.8–20.8) | 13.3 (9.6–15.7) | 1.6 (1.2–1.9) | 13.1 (10.5–15.8) | 10.3 (8.2–12.3) | 1.2 (1.1–1.2) |
| Iraq | 12.7 (10.7–14.7) | 57.5 (49.3–65.7) | 40.7 (35.6–45.8) | 3.8 (3–4.6) | 24.8 (19.9–29.8) | 16.8 (13.5–20.2) | 2.1 (1.9–2.3) |
| Jordan | 1.1 (0.8–1.4) | 7.7 (5.7–9.8) | 5.5 (4.2–6.9) | 0.9 (0.7–1) | 6.2 (5–7.5) | 4.4 (3.5–5.3) | 0.1 (0.1–0.2) |
| Kazakhstan | 6.7 (3.9–9.3) | 91 (56.1–128.9) | 70.1 (41.5–97) | 6.6 (5.2–7.9) | 92.1 (73.7–110.5) | 67.4 (53.9–80.9) | 1.9 (1.4–2.4) |

| | | | | | | | |
|---|---|---|---|---|---|---|---|
| Kenya | 87.4 (43.2–126.2) | 384.8 (193.9–572.4) | 266.3 (150.3–382.3) | 40.3 (32.2–48.3) | 237.9 (190.3–285.5) | 160.4 (128.3–192.5) | 62 (42–85) |
| Kyrgyz Republic | 16.8 (14.4–19.7) | 154.7 (131.6–180.1) | 110.7 (93.5–126.2) | 14.6 (11.7–17.6) | 134.7 (107.8–161.6) | 95.7 (76.5–114.8) | 5.9 (5.4–6.3) |
| Lao PDR | 47.5 (25.9–64.8) | 206 (119.5–288.5) | 153.4 (94.8–209.2) | 4.1 (3.3–4.9) | 138.5 (110.8–166.2) | 95.1 (76.1–114.1) | 30 (19–44) |
| Lesotho | 137.7 (75.4–202.9) | 905.8 (494.7–1323.8) | 658.7 (376.4–941.1) | 41.2 (32.9–49.4) | 472 (377.6–566.4) | 332.1 (265.7–398.6) | 224 (153–309) |
| Liberia | 129.2 (69.6–188.9) | 444.4 (232.5–615.3) | 303.8 (188.4–425.3) | 70.2 (56.2–84.3) | 234.8 (187.8–281.7) | 167.9 (134.3–201.4) | 74 (49–103) |
| Libya | 15.8 (8.4–23.1) | 75.9 (41–110.8) | 59 (33.9–84.1) | 5.4 (4.3–6.5) | 43.2 (34.6–51.8) | 32.7 (26.1–39.2) | 12 (7.1–19) |
| Madagascar | 69.8 (38.6–101) | 342 (192.8–497.4) | 233.6 (140.9–322.6) | 28 (22.4–33.6) | 212.4 (169.9–254.9) | 138 (110.4–165.6) | 46 (27–68) |
| Malawi | 48.2 (18.5–77.8) | 218.4 (85.5–360.8) | 144.9 (69.8–225.5) | 18.9 (15.1–22.6) | 146 (116.8–175.2) | 90.7 (72.6–108.9) | 37 (24–53) |
| Malaysia | 12.9 (11–14.5) | 114.9 (98.4–135.4) | 90.8 (78.2–106.4) | 11.2 (9–13.5) | 101.4 (81.1–121.7) | 80 (64–96) | 4.8 (4.3–5.3) |
| Maldives | 3.8 (2.8–4.7) | 44.7 (32.9–56.4) | 35.8 (26.4–45.2) | 2.8 (2.3–3.4) | 35.5 (28.4–42.6) | 29 (23.2–34.8) | 2.1 (1.9–2.3) |
| Mali | 9.8 (5.7–14) | 90.7 (53.1–125.5) | 50.9 (32.6–71.2) | 3.7 (3–4.4) | 63.4 (50.7–76.1) | 35.2 (28.1–42.2) | 9.1 (6–13) |
| Mauritania | 22.2 (12.7–31.6) | 132.3 (77.2–191.1) | 88.4 (55.2–123.7) | 8.6 (6.9–10.4) | 85.4 (68.3–102.5) | 55.1 (44.1–66.1) | 17 (10–25) |
| Mexico | 3.9 (3–5.1) | 29.7 (22.3–38.2) | 23.5 (17.2–29) | 2.0 (1.6–2.4) | 24.5 (19.6–29.4) | 18.6 (14.9–22.3) | 2 (1.9–2.2) |
| Mongolia | 201.3 (70.5–332.2) | 537.7 (183.7–851.3) | 434.1 (186–682.1) | 40.9 (32.7–49) | 173.4 (138.8–208.1) | 132.6 (106.1–159.1) | 10 (9.2–12) |
| Montenegro | 0.9 (0.7–1.1) | 17.7 (15–19.5) | 14.6 (12.4–17.5) | 0.9 (0.7–1.1) | 15.4 (12.3–18.4) | 12.7 (10.2–15.3) | 0.2 (0.2–0.2) |
| Morocco | 24.4 (20.3–28.5) | 123.9 (105.1–142.7) | 96 (82.3–112.4) | 21.2 (17–25.5) | 107.6 (86.1–129.2) | 84.3 (67.5–101.2) | 8.1 (5–12) |
| Mozambique | 259.8 (111.3–400.8) | 444 (189.4–692.6) | 362.2 (207.5–517) | 95.4 (76.3–114.5) | 492.3 (393.9–590.8) | 316.2 (253–379.5) | 37 (25–52) |
| Myanmar | 271.3 (142.8–392.7) | 339.7 (182.3–497.0) | 322 (199.8–442.2) | 169.2 (135.4–203.1) | 276.7 (221.3–332) | 248.9 (199.1–298.6) | 41 (27–59) |

| | | | | | | | |
|---|---|---|---|---|---|---|---|
| Namibia | 152.1 (99.9–206.4) | 698.9 (444.8–889.5) | 481.1 (336.7–641.4) | 79.6 (63.7–95.5) | 449.1 (359.3–538.9) | 312.7 (250.2–375.3) | 107 (80–137) |
| Nepal | 60.3 (30.7–89.9) | 312.6 (158.8–466.4) | 237.7 (129.3–346) | 20.4 (16.3–24.5) | 147.2 (117.8–176.7) | 110.1 (88.1–132.1) | 59 (32–92) |
| Nicaragua | 13.8 (10.2–17.9) | 54.5 (39.2–69.7) | 42.8 (32.1–53.5) | 8.9 (7.1–10.6) | 45.2 (36.2–54.2) | 34.3 (27.5–41.2) | 2.3 (1.9–2.6) |
| Niger | 17.2 (9.5–24.1) | 153.9 (85.5–213.8) | 85.8 (51.5–115.8) | 4.6 (3.6–5.5) | 93.9 (75.1–112.7) | 49.3 (39.4–59.1) | 17 (10–25) |
| Nigeria | 94.5 (51.3–137.8) | 316.3 (170.5–461.3) | 218.9 (134.9–303) | 10.8 (8.6–12.9) | 95.2 (76.1–114.2) | 58.4 (46.7–70.1) | 77 (49–110) |
| Pakistan | 100.1 (64.5–137) | 351.2 (224.7–477.8) | 263.2 (180.1–346.3) | 59.9 (47.9–71.8) | 200.9 (160.7–241) | 151.6 (121.3–181.9) | 20 (16–25) |
| Papua New Guinea | 353.2 (256.9–417.4) | 476.9 (353.2–600.5) | 433.0 (353.2–512.8) | 220.2 (176.2–264.3) | 402.2 (321.8–482.7) | 342.6 (274.1–411.1) | 50 (34–70) |
| Paraguay | 11.2 (9.2–12.6) | 60.1 (50.1–70.2) | 46.8 (39.7–52.5) | 9.6 (7.7–11.5) | 52.9 (42.3–63.4) | 40.2 (32.2–48.3) | 4.5 (3.8–5.2) |
| Peru | 26.8 (19.5–34.1) | 148.1 (111.1–189.3) | 120.0 (89.2–147.6) | 16.6 (13.3–19.9) | 123.4 (98.7–148.0) | 97.7 (78.2–117.2) | 8.8 (7.2–11) |
| Philippines | 209.4 (91.0–324.7) | 705.1 (310.0–1101.5) | 554.0 (276.6–831.5) | 129.5 (103.6–155.4) | 487.5 (390.0–585.1) | 378.4 (302.8–454.1) | 26 (22–29) |
| Republic of Moldova | 18.7 (15.3–20.2) | 91.2 (76.5–105.9) | 79.1 (66.8–91.5) | 15.7 (12.6–18.9) | 79.6 (63.7–95.6) | 69.5 (55.6–83.4) | 6.2 (5.4–7.1) |
| Russian Federation | 7.6 (4.5–10.6) | 59.5 (35.2–82.9) | 50.0 (30.2–69.2) | 7.7 (6.1–9.2) | 59.2 (47.4–71.1) | 50.3 (40.2–60.3) | 6.7 (6–7.4) |
| Rwanda | 12.3 (9.0–15.7) | 86.8 (63.1–110.4) | 57.0 (42.8–71.3) | 8.6 (6.9–10.4) | 72.0 (57.6–86.4) | 45.7 (36.5–54.8) | 7.5 (5.5–9.7) |
| Sao Tome and Principe | 21.0 (4.4–38.6) | 184.9 (36.2–329.5) | 116.3 (29.8–200.0) | 12.1 (9.7–14.6) | 103.7 (82.9–124.4) | 65.1 (52.1–78.1) | 26 (13–44) |
| Senegal | 21.5 (14.3–30.1) | 182.5 (128.8–246.9) | 116.6 (79.8–153.4) | 8.5 (6.8–10.2) | 137.0 (109.6–164.4) | 82.0 (65.6–98.4) | 18 (12–25) |
| Serbia | 1.3 (1.1–1.5) | 17.5 (14.8–18.9) | 14.8 (12.5–17.1) | 1.1 (0.9–1.3) | 14.4 (11.5–17.2) | 12.6 (10.1–15.1) | 0.5 (0.5–0.6) |
| Sierra Leone | 100.6 (56.6–147.7) | 431.8 (237.5–626.2) | 294.4 (179.2–409.6) | 73.9 (59.1–88.6) | 333.4 (266.7–400.0) | 227.7 (182.2–273.3) | 40 (26–56) |
| Solomon Islands | 24.6 (17.5–31.6) | 94.8 (67.3–122.2) | 65.7 (49.3–82.1) | 19.7 (15.8–23.7) | 75.1 (60.0–90.1) | 52.8 (42.3–63.4) | 7.3 (4.7–10) |

| | | | | | | | |
|---|---|---|---|---|---|---|---|
| South Africa | 224.0 (141.5–306.5) | 774.1 (488.0–1060.2) | 614.8 (409.8–818) | 97.0 (77.6–116.4) | 464.2 (371.4–557.0) | 357.8 (286.3–429.4) | 99 (59–150) |
| South Sudan | 106.6 (56.5–158.8) | 309.4 (170.2–464.1) | 226.0 (135.6–316.4) | 72.8 (58.2–87.3) | 200.8 (160.6–240.9) | 147.6 (118.0–177.1) | 42 (28–60) |
| Sri Lanka | 15.5 (10.8–19.6) | 80.2 (54.9–104.8) | 65.7 (45.5–79.7) | 4.6 (3.7–5.6) | 49.1 (39.3–59.0) | 38.5 (30.8–46.2) | 3.6 (2.9–4.4) |
| Sudan | 19.8 (12.2–27.3) | 97.6 (58.5–136.6) | 67.7 (44.4–88.8) | 11.3 (9.1–13.6) | 68.3 (54.6–81.9) | 46.2 (37.0–55.5) | 10 (6.6–15) |
| Suriname | 4.5 (3.8–5.8) | 37.7 (28.2–47.1) | 29.2 (20.6–36.1) | 3.8 (3.1–4.6) | 29.9 (23.9–35.9) | 22.9 (18.3–27.5) | 4.5 (3.7–5.4) |
| Swaziland | 80.6 (43.7–117.4) | 532.4 (294.2–770.6) | 365.8 (209.0–513.9) | 40.5 (32.4–48.6) | 378.3 (302.6–453.9) | 250.5 (200.4–300.6) | 84 (55–118) |
| Syrian Arab Republic | 6.6 (4.7–8.3) | 24.6 (17.8–31.4) | 18.7 (14.1–23.4) | 4.2 (3.4–5.1) | 20.1 (16.1–24.1) | 15.2 (12.1–18.2) | 0.1 (0.1–0.1) |
| Tajikistan | 18.2 (13.3–22.9) | 121.0 (88.7–153.4) | 82.6 (62.2–103.0) | 11.7 (9.4–14.0) | 91.2 (73–109.5) | 61.7 (49.4–74.1) | 8.5 (7.6–9.4) |
| Thailand | 28.2 (20.5–35.9) | 174.4 (127.8–221.0) | 150.8 (112.0–188.1) | 7.5 (6.0–9.0) | 147.5 (118.0–177.0) | 126.1 (100.9–151.3) | 16 (13–21) |
| Timor-Leste | 157.6 (89.2–228.1) | 702.9 (394.6–1011.2) | 494.9 (301.6–696.0) | 75.5 (60.4–90.6) | 454.6 (363.6–545.5) | 313.2 (250.6–375.8) | 90 (54–134) |
| Togo | 5.7 (4.5–6.6) | 58.7 (46.1–71.3) | 37.1 (29.7–44.5) | 2.6 (2.1–3.1) | 52.2 (41.8–62.7) | 31.9 (25.5–38.2) | 3.6 (2.5–5) |
| Tunisia | 11.3 (8.1–14.1) | 42.9 (31.6–54.2) | 35.1 (26.5–44.5) | 7.5 (6.0–9.0) | 34.8 (27.8–41.8) | 28.2 (22.6–33.8) | 1.3 (0.9–1.6) |
| Turkey | 2.8 (2.4–3.3) | 19.0 (15.8–22.2) | 15.6 (13.2–18.0) | 2.4 (2.0–2.9) | 17.0 (13.6–20.4) | 13.5 (10.8–16.2) | 0.4 (0.3–0.4) |
| Uganda | 77.7 (35.0–121.4) | 304 (135.1–472.9) | 198.8 (106.2–293.7) | 39.9 (31.9–47.9) | 238.1 (190.5–285.7) | 148.9 (119.1–178.6) | 35 (24–48) |
| Ukraine | 18.5 (10.7–25.7) | 89.2 (51.4–124.4) | 77.3 (47.7–106.8) | 8.3 (6.7–10.0) | 67.0 (53.6–80.5) | 57.7 (46.2–69.2) | 12 (9.9–13) |
| United Republic of Tanzania | 82.6 (22.8–145.5) | 356.1 (98.2–617.1) | 236.2 (89.6–384.4) | 48.1 (38.5–57.8) | 211.7 (169.4–254.1) | 140.0 (112.0–168.0) | 55 (32–84) |
| Uzbekistan | 33.7 (21.1–46.3) | 80.9 (51.1–110.7) | 66.7 (45.5–91) | 23.1 (18.4–27.7) | 60.0 (48.0–72.0) | 49.3 (39.5–59.2) | 5.4 (5.0–5.8) |
| Vanuatu | 16.4 (11.2–20.7) | 59.8 (40.3–76.2) | 40.0 (30.7–53.4) | 12.1 (9.6–14.5) | 43.0 (34.4–51.6) | 31.0 (24.8–37.2) | 5.4 (3.5–7.7) |

| | | | | | | | |
|---|---|---|---|---|---|---|---|
| Venezuela | 11.5<br>(8.4–14.1) | 58.0<br>(42.5–72.5) | 45.6<br>(34.0–56.1) | 7.2<br>(5.8–8.7) | 47.0<br>(37.6–56.4) | 36.1<br>(28.9–43.3) | 3.4<br>(2.7–4.2) |
| Vietnam | 35.7<br>(20.5–49.1) | 218.7<br>(126.9–311.9) | 176.2<br>(105.7–247.8) | 7.6<br>(6.1–9.1) | 136.0<br>(108.8–163.2) | 106.3<br>(85.0–127.5) | 12<br>(8.0–16) |
| Yemen | 15.7<br>(14.0–18.4) | 67.7<br>(56.4–79.0) | 48.0<br>(41.1–54.9) | 10.8<br>(8.7–13) | 50.7<br>(40.5–60.8) | 35.1<br>(28.0–42.1) | 6.6<br>(4.7–8.9) |
| Zambia | 80.6<br>(45.3–114.6) | 534.3<br>(302.4–766.2) | 330.3<br>(201.6–459.1) | 31.1<br>(24.9–37.4) | 339.5<br>(271.6–407.4) | 202.4<br>(161.9–242.9) | 86<br>(62–114) |
| Zimbabwe | 43.7<br>(30.8–56.7) | 306.9<br>(212.5–413.2) | 198.0<br>(143.4–252.6) | 19.0<br>(15.2–22.8) | 234.2<br>(187.3–281) | 143.4<br>(114.8–172.1) | 43<br>(33–54) |

**Table S9.3** HIV specific calibration targets for countries including the HIV structure for 2019 and for ages 0–99. Point values represent the mean with 95% confidence intervals in brackets.

| Country | HIV Prevalence (%) | ART Coverage (%) | Tuberculosis Incidence Rate in PLHIV (per 100,000 population per year) | Tuberculosis Mortality Rate in PLHIV (per 100,000 population per year) |
|---|---|---|---|---|
| Botswana | 16.5 (14.8–17.8) | 82 (74–89) | 123 (95–155) | 49 (23–81) |
| Central African Republic | 2.1 (1.8–2.7) | 46 (37–59) | 137 (88–195) | 61 (17–115) |
| Côte d'Ivoire | 1.7 (1.4–1.9) | 63 (54–74) | 24 (16–35) | 8.5 (2.3–15.5) |
| Cameroon | 2 (1.7–2.2) | 62 (54–68) | 48 (31–69) | 19 (5–37) |
| Gabon | 2.3 (1.7–3) | 51 (38–66) | 171 (65–327) | 26 (0–194) |
| Ghana | 1.1 (0.8–1.5) | 45 (32–61) | 30 (15–52) | 16 (0–40) |
| Gambia | 1.2 (0.9–1.5) | 29 (23–37) | 28 (21–37) | 7.9 (3.3–12.1) |
| Equatorial Guinea | 4.8 (3.5–6.5) | 35 (27–48) | 48 (41–55) | 16 (8–24) |
| Kenya | 2.9 (2.5–3.2) | 74 (65–86) | 70 (43–104) | 24 (6–48) |
| Lesotho | 16 (15.1–16.9) | 65 (61–70) | 403 (250–591) | 168 (38–328) |
| Mozambique | 7.2 (5.9–9.2) | 60 (48–74) | 122 (75–180) | 18 (4–38) |
| Malawi | 5.9 (5.2–5.9) | 79 (71–84) | 68 (36–109) | 23 (1–49) |
| Namibia | 8.4 (7.6–8.8) | 85 (79–91) | 158 (113–210) | 50 (20–88) |
| Rwanda | 1.8 (1.6–2) | 87 (77–95) | 12 (9.2–15) | 2.5 (1.1–4.3) |
| Swaziland | 17.4 (16.5–19.2) | 96 (88–100) | 218 (129–329) | 61 (11–125) |

| | | | | |
|---|---|---|---|---|
| Togo | 1.5<br>(1.2–1.7) | 64<br>(54–79) | 6<br>(4.8–7.4) | 1<br>(0.3–1.8) |
| United Republic of Tanzania | 2.9<br>(2.6–3.1) | 75<br>(67–81) | 56<br>(27–97) | 20<br>(0–48) |
| Uganda | 3.4<br>(3.2–3.6) | 84<br>(78–92) | 78<br>(46–119) | 19<br>(3–39) |
| South Africa | 12.8<br>(11.8–13.7) | 70<br>(64–74) | 357<br>(248–486) | 62<br>(0–168) |
| Zambia | 6.7<br>(5.4–7.3) | 85<br>(80–92) | 154<br>(100–220) | 53<br>(15–99) |
| Zimbabwe | 9.6<br>(8.2–10.9) | 85<br>(74–97) | 119<br>(88–154) | 31<br>(13–53) |

### 9.4 Calibrated *Status-Quo No-New-Vaccine* baseline trends

Each country was calibrated individually to either the nine or fourteen calibration targets as in section 9.3. Here we show the tuberculosis incidence and mortality rate targets for the selected grouping for reporting model outcomes. In Figure S9.1, looking by WHO region, we see the incidence rates are highest in AFR and SEAR, and lowest in AMR and EUR. In Figure S9.2, we see that correspondingly, the mortality rates are highest in AFR and SEAR, and lowest in AMR, EUR, and WPR. The estimated model medians for all WHO regions demonstrate decreasing trends from 2019 to 2050. In Figure S9.3 and Figure S9.4, we show the incidence and mortality rate trends by income group. Both incidence and mortality rates follow a trend with the highest estimated medians in lower-middle-income countries, followed by low-income countries and high-income countries, which aligns with the expectation of burden within each region. In Figure S9.5 and S9.6, we compare incidence and mortality rates between countries included on the WHO high TB burden list and all other countries modelled, and as expected, higher values are predicted for countries on the high TB burden list.

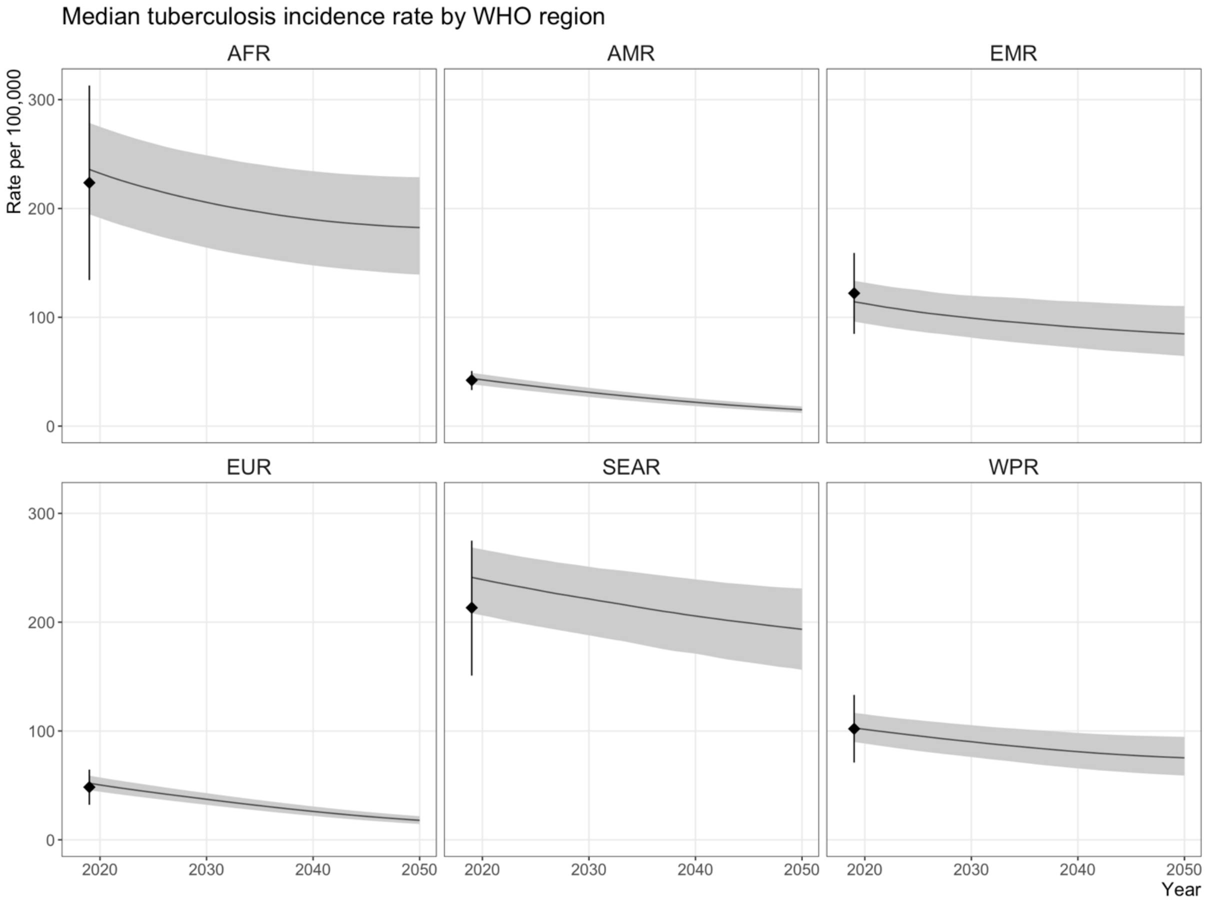


**Figure S9.1** Tuberculosis incidence rates for the *Status Quo No-New-Vaccine* baseline by WHO region. The black diamond is the WHO median estimate of the incidence in 2019 for the 105 modelled LMICs by WHO region with 95% uncertainty range. The black line is the model estimated median incidence rate, with shaded 95% uncertainty ranges.

*AFR = WHO African Region, AMR = WHO Region of the Americas, EMR = WHO Eastern Mediterranean Region, EUR = WHO European Region, SEAR = WHO South-East Asian Region, WPR = WHO Western Pacific Region*

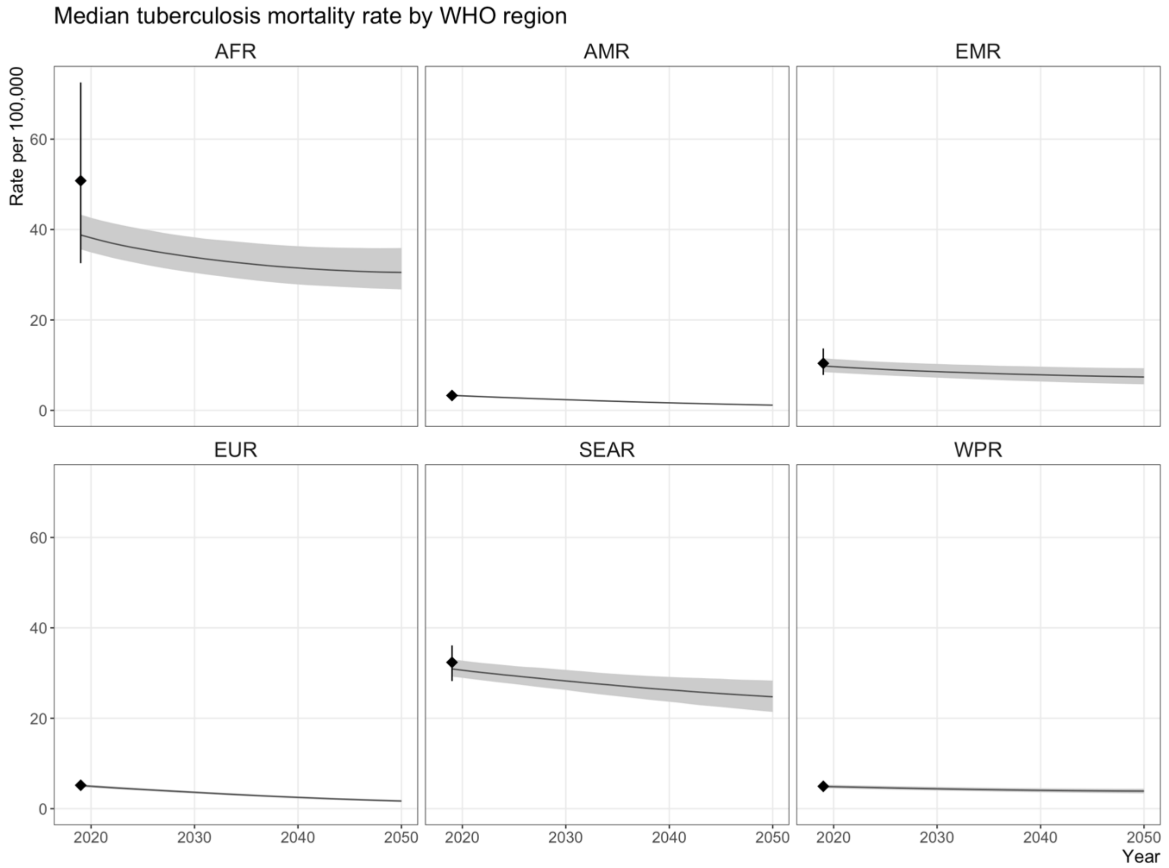


**Figure S9.2** Tuberculosis mortality rates for the *Status Quo No-New-Vaccine* baseline by WHO region. The black diamond is the WHO median estimate of the mortality rate in 2019 for the 105 modelled LMICs by WHO region with 95% uncertainty range. The black line is the model estimated median mortality rate, with shaded 95% uncertainty ranges.

*AFR = WHO African Region, AMR = WHO Region of the Americas, EMR = WHO Eastern Mediterranean Region, EUR = WHO European Region, SEAR = WHO South-East Asian Region, WPR = WHO Western Pacific Region*

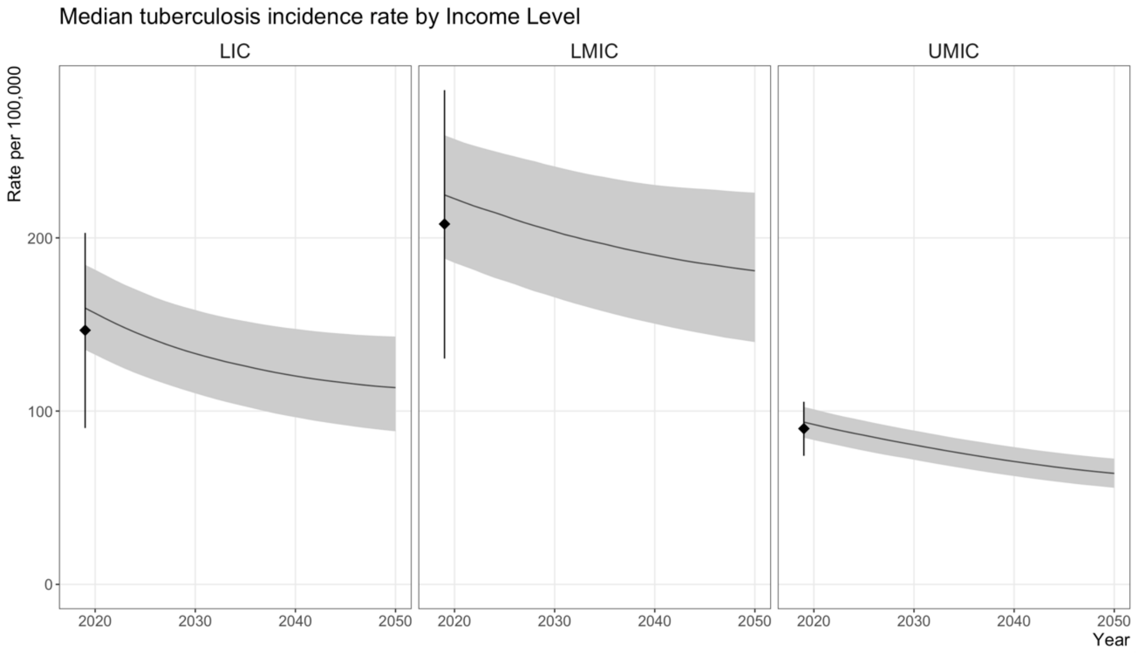


**Figure S9.3** Tuberculosis incidence rates for the *Status Quo No-New-Vaccine* baseline by income group. The black diamond is the WHO median estimate of the incidence rate in 2019 for the 105 modelled LMICs by income group with 95% uncertainty range. The black line is the model estimated median incidence rate, with shaded 95% uncertainty ranges.

*LIC = low-income countries, LMIC = lower-middle income countries, UMIC = upper-middle income countries*

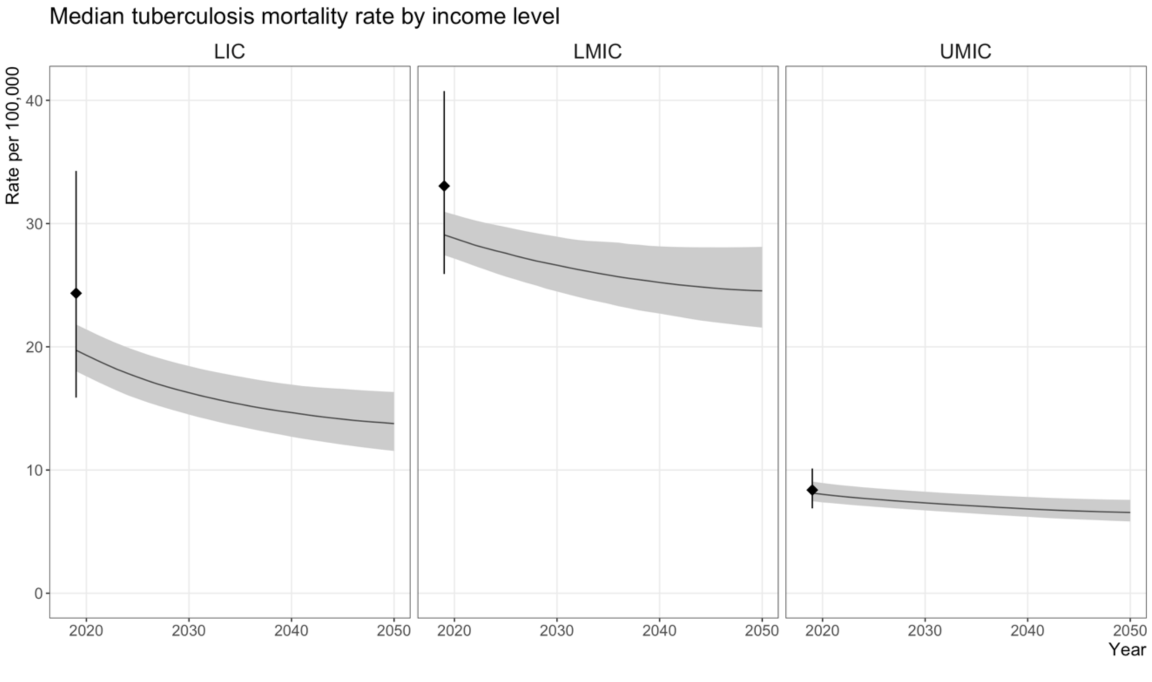


**Figure S9.4** Tuberculosis mortality rates for the *Status Quo No-New-Vaccine* baseline by income group. The black diamond is the WHO median estimate of the mortality rate in 2019 for the 105 modelled LMICs by income group with 95% uncertainty range. The black line is the model estimated median mortality rate, with shaded 95% uncertainty ranges.

*LIC = low-income countries, LMIC = lower-middle income countries, UMIC = upper-middle income countries*

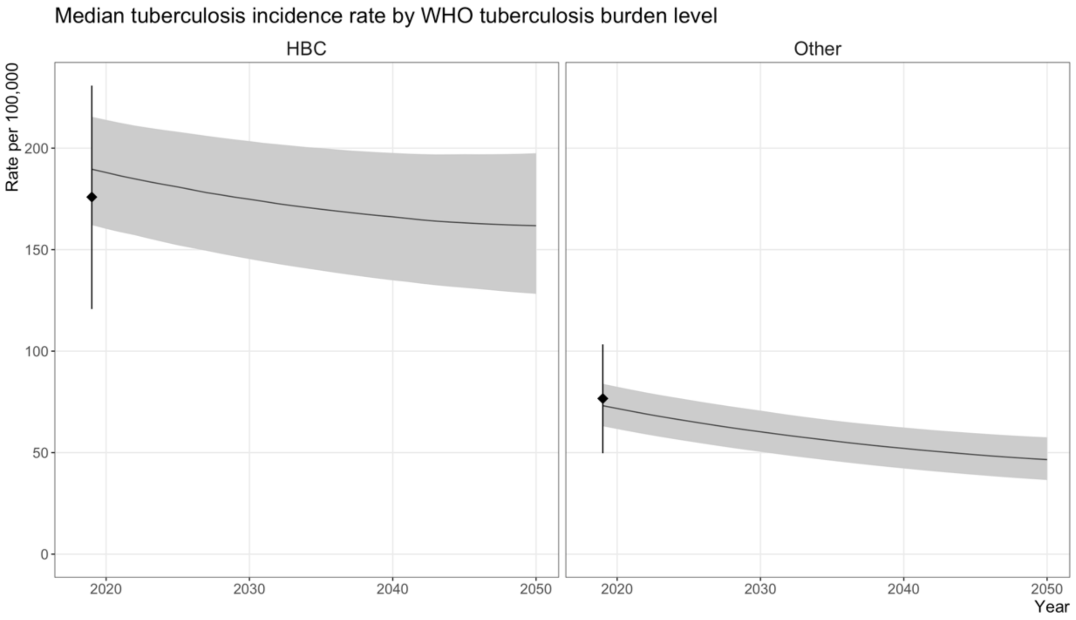


**Figure S9.5** Tuberculosis incidence rates for the *Status Quo No-New-Vaccine* baseline for the countries included on the WHO high-TB-burden list and for all other countries modelled. The black diamond is the WHO median estimate of the incidence rate in 2019 for the 105 modelled LMICs by burden level with 95% uncertainty range. The black line is the model estimated median incidence rate, with shaded 95% uncertainty ranges.

*HBC = high burden countries*

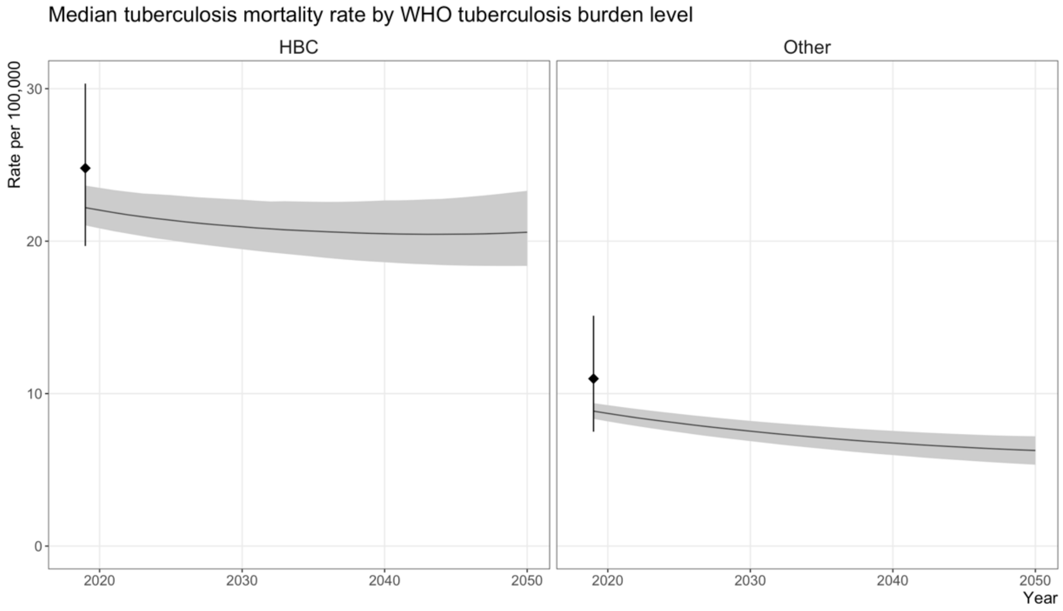


**Figure S9.6** Tuberculosis mortality rates for the *Status Quo No-New-Vaccine* baseline for the countries included on the WHO high-TB-burden list and for all other countries modelled. The black diamond is the WHO median estimate of the mortality rate in 2019 for the 105 modelled LMICs by burden level with 95% uncertainty range. The black line is the model estimated median mortality rate, with shaded 95% uncertainty ranges.

*HBC = high burden countries*

### 10. Vaccine Health Impact Results

#### 10.1 Incidence and mortality rate reductions and cumulative cases, treatments, and deaths averted

As stated in the main text, delivery of a 50% efficacy vaccine with an average of 10-years protection and medium coverage will have a substantial impact, which varies based on delivery and vaccine characteristics. For the adolescent/adult vaccine, compared to the *Basecase* implementation, the *Accelerated Scale-up* scenario averted approximately double the number of cases, treatments, and deaths by 2050, and almost eight times as many as the *Routine Only* scenario, demonstrating the benefits of instantly introducing and scaling-up to coverage, as well as including a campaign for ages ten and over. We performed scenario analyses by varying certain vaccine and delivery characteristics, the results of which are presented in Table S10.1 (adolescent/adult vaccine) and Table S10.2 (infant vaccine) below, as the median estimate and 95% uncertainty range. Decreasing the target vaccine coverage correspondingly decreased the health impact estimates, and increasing the target vaccine coverage, increasing the duration of protection to lifelong, or increasing the vaccine efficacy increases the health impact estimates.

The order of vaccine scenario health impact results within each table is as follows:

Primary scenarios (as in main text)

- *Basecase*, medium coverage, 50% efficacy, 10-years protection
- *Accelerated Scale-up*, medium coverage, 50% efficacy, 10-years protection
- *Routine Only*, medium coverage, 50% efficacy, 10-years protection (adolescent/adult vaccine only)

Low and high coverage targets

- *Basecase*, low coverage, 50% efficacy, 10-years protection
- *Basecase*, high coverage, 50% efficacy, 10-years protection
- *Accelerated Scale-up*, low coverage, 50% efficacy, 10-years protection
- *Accelerated Scale-up*, high coverage, 50% efficacy, 10-years protection
- *Routine Only*, low coverage, 50% efficacy, 10-years protection (adolescent/adult vaccine only)
- *Routine Only*, high coverage, 50% efficacy, 10-years protection (adolescent/adult vaccine only)

Lifelong duration of protection

- *Basecase*, medium coverage, 50% efficacy, lifelong protection
- *Accelerated Scale-up*, medium coverage, 50% efficacy, lifelong protection
- *Routine Only*, medium coverage, 50% efficacy, lifelong protection (adolescent/adult vaccine only)

75% efficacy (adolescent/adult vaccine only)

- *Basecase*, medium coverage, 75% efficacy, 10-years protection
- *Accelerated Scale-up*, medium coverage, 75% efficacy, 10-years protection

**Table S10.1** Estimated health impact in 2050 by WHO region, income level, and tuberculosis burden level for the adolescent/adult vaccine scenarios

| | Health Impact Measure | All modelled countries | WHO Region | | | | | | TB Burden Level | | World Bank Income Group | | |
|---|---|---|---|---|---|---|---|---|---|---|---|---|---|
| **Vaccine Scenario** | | | **AFR** | **AMR** | **EMR** | **EUR** | **SEAR** | **WPR** | **HBC** | **All other countries** | **LIC** | **LMIC** | **UMIC** |
| **Primary scenarios** | | | | | | | | | | | | | |
| *Basecase*<br><br>Medium coverage, 50% efficacy, 10 years protection | Averted cases before 2050 | 44.0m (37.2–51.6) | 13.9m (11.7–16.7) | 0.5m (0.5–0.6) | 3.9m (3.1–4.8) | 0.3m (0.3–0.4) | 19.5m (15.9–23.1) | 5.9m (5.0–6.9) | 39.8m (33.7–46.7) | 4.1m (3.4–4.9) | 5.0m (4.1–6.0) | 34.3m (28.6–40.3) | 4.7m (4.1–5.4) |
| | Averted tx before 2050 | 24.9m (21.9–27.3) | 6.3m (5.7–6.8) | 0.4m (0.3–0.4) | 2.4m (2.0–2.8) | 0.2m (0.2–0.3) | 11.7m (10.1–13.4) | 3.8m (3.3–4.2) | 22.6m (19.9–24.8) | 2.3m (2.0–2.6) | 2.9m (2.5–3.2) | 19.0m (16.6–21.2) | 2.9m (2.7–3.2) |
| | Averted deaths before 2050 | 5.0m (4.6–5.4) | 2.1m (1.9–2.3) | 0.04m (0.03–0.04) | 0.3m (0.2–0.4) | 0.028m (0.026–0.031) | 2.2m (2.0–2.6) | 0.3m (0.2–0.3) | 4.5m (4.2–4.9) | 0.5m (0.4–0.5) | 0.6m (0.5–0.6) | 4.1m (3.7–4.4) | 0.4m (0.3–0.5) |
| | IRR in 2050 (%) | 25.4% (23.9–27.7) | 27.0% (25.7–31.3) | 15.9% (15.2–16.9) | 26.7% (23.7–31.6) | 20.2% (18.6–22.6) | 25.4% (23.3–28.2) | 19.8% (18.3–22.2) | 25.4% (23.8–27.9) | 25.1% (24.1–26.6) | 27.3% (26.0–29.1) | 26.1% (24.3–28.9) | 16.7% (15.8–18.0) |
| | MRR in 2050 (%) | 27.1% (25.6–30.1) | 27.7% (26.3–33.3) | 17.7% (16.8–18.7) | 28.1% (25.0–32.8) | 19.9% (18.6–21.6) | 26.5% (24.3–29.4) | 23.1% (21.2–25.8) | 27.3% (25.5–30.6) | 25.9% (25.0–27.1) | 27.8% (26.6–29.4) | 27.6% (25.8–31.3) | 19.4% (18.1–21.3) |
| *Accelerated Scale–up*<br><br>Medium coverage, 50% efficacy, 10 years protection | Averted cases before 2050 | 65.5m (55.6–76.0) | 19.5m (16.7–23.1) | 0.8m (0.7–1.0) | 5.4m (4.3–6.7) | 0.6m (0.5–0.7) | 31.0m (25.8–36.4) | 8.1m (6.9–9.5) | 58.6m (49.9–67.9) | 7.0m (5.8–8.2) | 7.5m (6.2–9.0) | 51.7m (43.6–60.2) | 6.4m (5.6–7.2) |
| | Averted tx before 2050 | 38.6m (34.4–42.3) | 9.2m (8.5–9.9) | 0.6m (0.5–0.7) | 3.4m (2.9–4.0) | 0.4m (0.4–0.5) | 19.5m (16.8–22.2) | 5.3m (4.8–5.9) | 34.6m (30.7–37.9) | 4.0m (3.5–4.4) | 4.5m (4.0–5.0) | 30.0m (26.5–33.3) | 4.1m (3.7–4.4) |
| | Averted deaths before 2050 | 7.9m (7.3–8.5) | 3.1m (2.9–3.4) | 0.06m (0.05–0.06) | 0.5m (0.4–0.6) | 0.06m (0.05–0.06) | 3.8m (3.3–4.3) | 0.40m (0.36–0.45) | 7.0m (6.4–7.6) | 0.8m (0.8–0.9) | 0.9m (0.8–1.0) | 6.5m (5.9–7.0) | 0.5m (0.4–0.6) |
| | IRR in 2050 (%) | 25.2% (23.9–27.5) | 27.6% (26.3–32.1) | 15.2% (14.4–16.2) | 27.1% (24.5–31.4) | 18.4% (16.4–21.6) | 24.7% (22.8–27.3) | 19.4% (18.1–21.3) | 25.2% (23.8–27.6) | 25.3% (24.5–26.8) | 27.5% (26.3–29.2) | 25.9% (24.3–28.6) | 16.3% (15.5–17.3) |
| | MRR in 2050 (%) | 26.7% (25.2–29.9) | 28.2% (26.8–34.6) | 16.2% (15.3–17.3) | 27.9% (25.2–32.3) | 18.1% (16.5–20.7) | 25.3% (23.2–28.2) | 21.8% (20.2–24.3) | 26.8% (25.1–30.4) | 26.1% (25.3–27.2) | 27.7% (26.6–29.2) | 27.2% (25.5–31.0) | 18.4% (17.3–20.0) |
| *Routine Only*<br><br>Medium coverage, 50% efficacy, | Averted cases before 2050 | 8.8m (7.6–10.1) | 3.5m (3.0–3.9) | 0.04m (0.03–0.05) | 0.9m (0.7–1.2) | 0.02m (0.02–0.03) | 3.4m (2.6–4.4) | 1.0m (0.8–1.2) | 8.1m (7.0–9.3) | 0.7m (0.6–0.8) | 1.1m (0.9–1.3) | 7.2m (6.2–8.3) | 0.5m (0.4–0.7) |
| | Averted tx before 2050 | 4.1m (3.7–4.6) | 1.2m (1.1–1.4) | 0.03m (0.02–0.03) | 0.5m (0.4–0.6) | 0.01m (0.01–0.02) | 1.8m (1.4–2.2) | 0.6m (0.5–0.7) | 3.8m (3.4–4.2) | 0.3m (0.3–0.4) | 0.6m (0.5–0.6) | 3.3m (2.9–3.8) | 0.3m (0.2–0.3) |

| | | | | | | | | | | | | | |
|---|---|---|---|---|---|---|---|---|---|---|---|---|---|
| 10 years protection | Averted deaths before 2050 | 1.1m (0.9–1.2) | 0.5m (0.4–0.6) | 0.003m (0.003–0.004) | 0.08m (0.06–0.10) | 0.002m (0.002–0.003) | 0.4m (0.3–0.5) | 0.1m (0.0–0.1) | 1.0m (0.8–1.1) | 0.08m (0.07–0.09) | 0.12m (0.10–0.14) | 0.9m (0.7–1.0) | 0.1m (0.0–0.1) |
| | IRR in 2050 (%) | 9.9% (9.0–11.6) | 11.2% (10.3–14.7) | 3.4% (3.1–3.9) | 11.9% (9.9–15.3) | 4.1% (3.4–5.2) | 9.1% (7.8–11.1) | 7.7% (6.5–9.5) | 10.2% (9.1–12.0) | 8.0% (7.3–9.2) | 10.5% (9.6–11.9) | 10.4% (9.2–12.5) | 5.2% (4.4–6.3) |
| | MRR in 2050 (%) | 9.9% (8.9–12.3) | 10.7% (9.7–15.2) | 3.7% (3.3–4.2) | 11.9% (9.9–15.1) | 3.8% (3.3–4.5) | 8.7% (7.3–10.7) | 9.2% (7.5–11.7) | 10.2% (9.1–12.9) | 7.2% (6.5–8.1) | 9.6% (8.8–10.7) | 10.2% (9.0–13.1) | 6.2% (5.2–7.8) |
| **Low and high coverage** | | | | | | | | | | | | | |
| *Basecase* Low coverage, 50% efficacy, 10 years protection | Averted cases before 2050 | 33.5m (28.5–39.2) | 10.7m (9.1–12.8) | 0.4m (0.3–0.5) | 3.0m (2.4–3.7) | 0.3m (0.2–0.3) | 14.7m (12.1–17.5) | 4.4m (3.8–5.2) | 30.4m (25.7–35.5) | 3.1m (2.6–3.7) | 3.9m (3.2–4.6) | 26.1m (22.0–30.7) | 3.5m (3.0–4.0) |
| | Averted tx before 2050 | 18.8m (16.6–20.6) | 4.8m (4.4–5.1) | 0.3m (0.2–0.3) | 1.8m (1.5–2.2) | 0.2m (0.1–0.2) | 8.8m (7.6–10.1) | 2.8m (2.5–3.2) | 17.0m (15.1–18.8) | 1.7m (1.5–1.9) | 2.2m (1.9–2.5) | 14.4m (12.6–16.0) | 2.2m (2.0–2.3) |
| | Averted deaths before 2050 | 3.8m (3.5–4.1) | 1.6m (1.5–1.8) | 0.03m (0.02–0.03) | 0.2m (0.2–0.3) | 0.021m (0.019–0.023) | 1.7m (1.5–1.9) | 0.21m (0.18–0.23) | 3.5m (3.2–3.8) | 0.3m (0.3–0.4) | 0.4m (0.4–0.5) | 3.1m (2.8–3.4) | 0.3m (0.2–0.4) |
| | IRR in 2050 (%) | 20.2% (19.0–22.2) | 21.6% (20.5–25.4) | 12.2% (11.6–13.0) | 21.5% (19.0–25.8) | 15.5% (14.2–17.5) | 20.1% (18.3–22.6) | 15.7% (14.4–17.7) | 20.3% (18.9–22.5) | 19.7% (18.9–21.0) | 21.7% (20.6–23.4) | 20.8% (19.3–23.3) | 13.0% (12.2–14.1) |
| | MRR in 2050 (%) | 21.5% (20.2–24.2) | 22.0% (20.9–27.0) | 13.5% (12.8–14.4) | 22.5% (19.9–26.6) | 15.2% (14.1–16.6) | 20.9% (19.0–23.4) | 18.3% (16.6–20.7) | 21.7% (20.2–24.6) | 20.1% (19.3–21.2) | 22.0% (20.9–23.4) | 22.0% (20.4–25.2) | 15.1% (14.0–16.8) |
| *Basecase* High coverage, 50% efficacy, 10 years protection | Averted cases before 2050 | 54.2m (45.7–63.6) | 17.0m (14.2–20.5) | 0.7m (0.6–0.8) | 4.8m (3.8–5.9) | 0.4m (0.4–0.5) | 24.1m (19.7–28.5) | 7.3m (6.2–8.6) | 49.1m (41.4–57.6) | 5.1m (4.2–6.1) | 6.2m (5.0–7.4) | 42.1m (35.1–49.7) | 6.0m (5.2–6.8) |
| | Averted tx before 2050 | 30.8m (27.1–33.9) | 7.7m (7.0–8.4) | 0.5m (0.4–0.5) | 2.9m (2.4–3.4) | 0.29m (0.26–0.33) | 14.5m (12.5–16.6) | 4.7m (4.2–5.3) | 27.9m (24.6–30.7) | 2.9m (2.5–3.2) | 3.5m (3.1–4.0) | 23.5m (20.5–26.2) | 3.7m (3.4–4.0) |
| | Averted deaths before 2050 | 6.1m (5.7–6.7) | 2.6m (2.3–2.9) | 0.0m (0.0–0.1) | 0.4m (0.3–0.5) | 0.04m (0.03–0.04) | 2.8m (2.4–3.2) | 0.3m (0.3–0.4) | 5.6m (5.1–6.1) | 0.6m (0.5–0.6) | 0.7m (0.6–0.8) | 5.0m (4.6–5.4) | 0.5m (0.4–0.6) |
| | IRR in 2050 (%) | 30.3% (28.6–32.8) | 32.1% (30.6–36.7) | 19.5% (18.6–20.6) | 31.6% (28.2–37.0) | 24.6% (22.9–27.3) | 30.3% (28.0–33.5) | 23.9% (22.2–26.4) | 30.3% (28.5–33.0) | 30.2% (29.1–31.7) | 32.5% (31.0–34.5) | 31.1% (29.0–34.1) | 20.2% (19.2–21.8) |
| | MRR in 2050 (%) | 32.4% (30.7–35.7) | 33.1% (31.6–39.1) | 21.7% (20.7–22.9) | 33.3% (29.9–38.5) | 24.4% (22.9–26.4) | 31.8% (29.3–35.1) | 27.8% (25.6–30.8) | 32.6% (30.6–36.2) | 31.4% (30.4–32.7) | 33.3% (32.0–35.1) | 33.0% (31.0–37.0) | 23.5% (22.0–25.7) |
| *Accelerated Scale–up* Low coverage, 50% efficacy, | Averted cases before 2050 | 50.9m (43.4–59.0) | 15.3m (13.3–18.1) | 0.6m (0.5–0.7) | 4.3m (3.4–5.2) | 0.5m (0.4–0.6) | 24.0m (19.9–28.3) | 6.2m (5.3–7.2) | 45.6m (38.9–52.8) | 5.4m (4.5–6.3) | 5.9m (4.9–7.1) | 40.2m (34.1–46.8) | 4.8m (4.2–5.4) |
| | Averted tx before 2050 | 29.7m (26.6–32.6) | 7.2m (6.6–7.7) | 0.4m (0.4–0.5) | 2.6m (2.2–3.1) | 0.3m (0.3–0.4) | 15.0m (13.0–17.2) | 4.0m (3.6–4.5) | 26.6m (23.8–29.3) | 3.1m (2.7–3.4) | 3.5m (3.1–3.9) | 23.2m (20.6–25.8) | 3.0m (2.8–3.3) |

| | | | | | | | | | | | | | |
|---|---|---|---|---|---|---|---|---|---|---|---|---|---|
| 10 years protection | Averted deaths before 2050 | 6.1m (5.7–6.6) | 2.5m (2.2–2.7) | 0.04m (0.04–0.05) | 0.4m (0.3–0.4) | 0.04m (0.04–0.05) | 2.9m (2.5–3.4) | 0.3m (0.3–0.3) | 5.5m (5.0–6.0) | 0.7m (0.6–0.7) | 0.7m (0.6–0.8) | 5.0m (4.6–5.5) | 0.4m (0.3–0.5) |
| | IRR in 2050 (%) | 20.8% (19.5–22.8) | 22.7% (21.6–26.8) | 12.0% (11.3–12.8) | 22.4% (20.2–26.3) | 14.9% (13.2–17.8) | 20.3% (18.6–22.7) | 15.7% (14.6–17.5) | 20.8% (19.5–22.9) | 20.7% (20.0–22.1) | 22.7% (21.6–24.2) | 21.4% (19.9–23.8) | 13.0% (12.3–13.8) |
| | MRR in 2050 (%) | 21.9% (20.6–24.8) | 23.2% (21.9–28.9) | 12.8% (12.0–13.7) | 23.1% (20.7–27.0) | 14.6% (13.2–16.9) | 20.7% (18.9–23.4) | 17.8% (16.4–20.0) | 22.0% (20.6–25.2) | 21.2% (20.5–22.3) | 22.7% (21.8–24.2) | 22.4% (20.9–25.9) | 14.7% (13.8–16.1) |
| *Accelerated Scale–up* High coverage, 50% efficacy, 10 years protection | Averted cases before 2050 | 79.6m (67.5–92.6) | 23.5m (20.0–28.0) | 1.0m (0.9–1.2) | 6.5m (5.2–8.1) | 0.8m (0.7–0.9) | 37.7m (31.4–44.1) | 10.0m (8.5–11.7) | 71.1m (60.4–82.7) | 8.5m (7.1–10.1) | 9.1m (7.5–10.8) | 62.6m (52.7–73.1) | 8.0m (7.0–9.0) |
| | Averted tx before 2050 | 47.0m (41.9–51.7) | 11.2m (10.2–12.0) | 0.7m (0.7–0.8) | 4.1m (3.5–4.8) | 0.6m (0.5–0.6) | 23.7m (20.5–27.0) | 6.6m (5.9–7.3) | 42.2m (37.4–46.4) | 4.9m (4.3–5.4) | 5.4m (4.8–6.0) | 36.5m (32.3–40.6) | 5.1m (4.7–5.5) |
| | Averted deaths before 2050 | 9.5m (8.8–10.3) | 3.8m (3.5–4.2) | 0.1m (0.1–0.1) | 0.5m (0.4–0.7) | 0.07m (0.06–0.07) | 4.6m (4.0–5.2) | 0.5m (0.4–0.5) | 8.5m (7.8–9.2) | 1.0m (0.9–1.1) | 1.1m (1.0–1.2) | 7.8m (7.2–8.5) | 0.6m (0.5–0.8) |
| | IRR in 2050 (%) | 29.4% (28.0–31.9) | 32.1% (30.7–37.0) | 18.3% (17.2–19.4) | 31.4% (28.6–36.1) | 21.5% (19.5–25.0) | 28.8% (26.7–31.7) | 22.9% (21.5–25.0) | 29.4% (27.9–32.0) | 29.6% (28.7–31.2) | 32.0% (30.7–33.9) | 30.2% (28.4–33.1) | 19.5% (18.6–20.6) |
| | MRR in 2050 (%) | 31.2% (29.6–34.7) | 33.0% (31.4–39.8) | 19.4% (18.3–20.7) | 32.4% (29.4–37.1) | 21.4% (19.6–24.2) | 29.6% (27.3–32.7) | 25.7% (23.8–28.3) | 31.3% (29.5–35.1) | 30.6% (29.8–31.8) | 32.3% (31.2–34.0) | 31.8% (29.9–35.9) | 21.9% (20.7–23.7) |
| *Routine Only* Low coverage, 50% efficacy, 10 years protection | Averted cases before 2050 | 7.8m (6.8–8.9) | 3.1m (2.6–3.5) | 0.04m (0.03–0.04) | 0.8m (0.6–1.1) | 0.02m (0.02–0.03) | 3.0m (2.3–3.8) | 0.9m (0.7–1.1) | 7.2m (6.2–8.2) | 0.6m (0.5–0.7) | 1.0m (0.8–1.2) | 6.3m (5.5–7.3) | 0.5m (0.4–0.6) |
| | Averted tx before 2050 | 3.6m (3.3–4.0) | 1.1m (1.0–1.2) | 0.022m (0.019–0.025) | 0.4m (0.4–0.6) | 0.013m (0.011–0.015) | 1.5m (1.3–1.9) | 0.5m (0.4–0.6) | 3.3m (3.0–3.7) | 0.29m (0.26–0.33) | 0.5m (0.4–0.6) | 2.9m (2.6–3.3) | 0.2m (0.2–0.3) |
| | Averted deaths before 2050 | 0.9m (0.8–1.1) | 0.5m (0.4–0.6) | 0.003m (0.002–0.003) | 0.1m (0.0–0.1) | 0.0019m (0.0017–0.0022) | 0.3m (0.3–0.4) | 0.1m (0.0–0.1) | 0.9m (0.7–1.0) | 0.07m (0.06–0.08) | 0.1m (0.1–0.1) | 0.8m (0.7–0.9) | 0.0m (0.0–0.1) |
| | IRR in 2050 (%) | 8.8% (7.9–10.3) | 9.9% (9.2–13.1) | 3.0% (2.7–3.5) | 10.6% (8.7–13.6) | 3.6% (3.0–4.6) | 8.1% (6.9–9.9) | 6.8% (5.8–8.5) | 9.0% (8.1–10.6) | 7.1% (6.4–8.1) | 9.3% (8.5–10.5) | 9.2% (8.1–11.1) | 4.6% (3.9–5.6) |
| | MRR in 2050 (%) | 8.8% (7.8–10.9) | 9.5% (8.6–13.5) | 3.3% (2.9–3.7) | 10.6% (8.7–13.4) | 3.3% (2.9–4.0) | 7.7% (6.5–9.5) | 8.1% (6.6–10.3) | 9.1% (8.0–11.4) | 6.3% (5.7–7.2) | 8.5% (7.8–9.5) | 9.1% (8.0–11.6) | 5.5% (4.5–6.9) |
| *Routine Only* High coverage, 50% efficacy, 10 years protection | Averted cases before 2050 | 9.8m (8.5–11.3) | 3.9m (3.3–4.4) | 0.0m (0.0–0.1) | 1.0m (0.8–1.3) | 0.03m (0.02–0.03) | 3.8m (2.9–4.9) | 1.1m (0.9–1.4) | 9.1m (7.8–10.4) | 0.8m (0.6–0.9) | 1.2m (1.0–1.5) | 8.0m (6.9–9.3) | 0.6m (0.5–0.7) |
| | Averted tx before 2050 | 4.6m (4.2–5.1) | 1.4m (1.2–1.5) | 0.03m (0.02–0.03) | 0.6m (0.4–0.7) | 0.02m (0.01–0.02) | 2.0m (1.6–2.4) | 0.6m (0.5–0.8) | 4.2m (3.8–4.7) | 0.4m (0.3–0.4) | 0.6m (0.5–0.7) | 3.6m (3.2–4.2) | 0.3m (0.3–0.4) |
| | Averted deaths before 2050 | 1.2m (1.0–1.3) | 0.6m (0.5–0.7) | 0.004m (0.003–0.004) | 0.08m (0.06–0.11) | 0.002m (0.002–0.003) | 0.4m (0.3–0.6) | 0.07m (0.05–0.09) | 1.1m (0.9–1.3) | 0.09m (0.07–0.10) | 0.1m (0.1–0.2) | 1.0m (0.8–1.2) | 0.1m (0.0–0.1) |

| | | | | | | | | | | | | | |
|---|---|---|---|---|---|---|---|---|---|---|---|---|---|
| | IRR in 2050 (%) | 11.0% (10.0–12.9) | 12.5% (11.5–16.3) | 3.8% (3.4–4.4) | 13.2% (11.0–17.0) | 4.6% (3.8–5.8) | 10.1% (8.6–12.3) | 8.6% (7.3–10.6) | 11.3% (10.1–13.3) | 8.9% (8.1–10.2) | 11.7% (10.7–13.2) | 11.5% (10.2–13.8) | 5.8% (4.9–7.0) |
| | MRR in 2050 (%) | 11.0% (9.9–13.6) | 12.0% (10.9–16.9) | 4.1% (3.7–4.7) | 13.3% (11.0–16.8) | 4.2% (3.6–5.0) | 9.7% (8.2–11.9) | 10.3% (8.4–13.0) | 11.4% (10.1–14.3) | 8.0% (7.3–9.0) | 10.7% (9.8–11.9) | 11.4% (10.0–14.5) | 6.9% (5.8–8.7) |
| **Lifelong protection** | | | | | | | | | | | | | |
| *Basecase*<br><br>Medium coverage, 50% efficacy, lifelong protection | Averted cases before 2050 | 69.7m (58.3–82.0) | 21.9m (18.1–26.6) | 0.9m (0.8–1.0) | 6.2m (4.9–7.7) | 0.6m (0.5–0.6) | 30.3m (24.7–35.8) | 9.8m (8.3–11.7) | 63.2m (52.9–74.5) | 6.5m (5.3–7.7) | 7.8m (6.4–9.4) | 53.7m (44.5–63.7) | 8.2m (7.1–9.4) |
| | Averted tx before 2050 | 38.4m (33.9–42.4) | 9.7m (8.7–10.5) | 0.6m (0.5–0.7) | 3.7m (3.1–4.4) | 0.4m (0.3–0.4) | 17.8m (15.3–20.3) | 6.2m (5.5–6.9) | 34.9m (30.7–38.5) | 3.5m (3.1–3.9) | 4.4m (3.8–4.9) | 29.1m (25.3–32.5) | 5.0m (4.5–5.3) |
| | Averted deaths before 2050 | 7.5m (6.9–8.2) | 3.2m (2.9–3.5) | 0.06m (0.05–0.06) | 0.5m (0.4–0.6) | 0.04m (0.04–0.05) | 3.3m (2.9–3.8) | 0.4m (0.4–0.5) | 6.8m (6.3–7.5) | 0.7m (0.6–0.8) | 0.8m (0.7–0.9) | 6.1m (5.6–6.6) | 0.6m (0.5–0.8) |
| | IRR in 2050 (%) | 50.2% (48.1–53.2) | 51.4% (49.4–56.5) | 40.7% (39.2–42.4) | 52.8% (48.4–59.2) | 42.7% (40.8–45.2) | 50.2% (47.1–53.8) | 44.8% (41.9–48.7) | 50.6% (48.2–53.8) | 47.5% (46.0–49.4) | 51.2% (49.2–53.6) | 51.0% (48.4–54.7) | 42.0% (40.0–44.9) |
| | MRR in 2050 (%) | 49.5% (47.3–53.2) | 49.5% (47.5–55.8) | 40.8% (39.4–42.5) | 52.2% (48.1–58.2) | 40.8% (39.1–42.8) | 49.3% (46.3–52.9) | 45.7% (42.6–49.7) | 50.1% (47.7–54.1) | 45.1% (43.4–46.8) | 48.9% (47.0–51.1) | 50.2% (47.6–54.5) | 42.9% (40.1–46.6) |
| *Accelerated Scale–up*<br><br>Medium coverage, 50% efficacy, lifelong protection | Averted cases before 2050 | 116.7m (98.4–136.4) | 34.1m (28.4–41.2) | 1.6m (1.3–1.8) | 9.5m (7.6–11.7) | 1.2m (1.0–1.3) | 55.3m (45.6–64.8) | 15.0m (12.7–17.7) | 104.2m (88.0–121.9) | 12.4m (10.2–14.7) | 13.2m (10.8–15.9) | 91.4m (76.3–107.3) | 12.1m (10.6–13.7) |
| | Averted tx before 2050 | 67.9m (60.1–74.7) | 16.1m (14.5–17.3) | 1.1m (1.0–1.2) | 5.9m (5.0–6.9) | 0.8m (0.7–0.9) | 34.2m (29.5–38.9) | 9.7m (8.6–10.8) | 60.9m (53.8–67.2) | 7.0m (6.2–7.7) | 7.8m (6.8–8.6) | 52.5m (46.1–58.5) | 7.6m (6.9–8.1) |
| | Averted deaths before 2050 | 13.4m (12.4–14.5) | 5.2m (4.8–5.8) | 0.11m (0.10–0.12) | 0.8m (0.6–0.9) | 0.10m (0.09–0.11) | 6.5m (5.7–7.4) | 0.7m (0.6–0.8) | 11.9m (11.0–13.0) | 1.5m (1.3–1.6) | 1.5m (1.4–1.7) | 10.9m (10.1–11.9) | 0.9m (0.8–1.2) |
| | IRR in 2050 (%) | 55.6% (53.5–58.7) | 57.7% (55.8–63.0) | 43.4% (41.9–45.1) | 57.4% (52.8–64.1) | 47.1% (44.6–50.6) | 55.7% (52.4–59.6) | 47.9% (45.1–51.9) | 55.7% (53.3–59.1) | 54.9% (53.6–56.8) | 58.0% (56.1–60.4) | 56.6% (53.9–60.3) | 44.4% (42.6–47.2) |
| | MRR in 2050 (%) | 55.7% (53.4–59.8) | 56.5% (54.5–63.5) | 43.7% (42.2–45.4) | 57.1% (52.7–63.3) | 46.1% (43.7–49.0) | 55.2% (51.8–59.3) | 49.4% (46.3–53.5) | 55.9% (53.4–60.4) | 54.4% (53.1–56.0) | 56.7% (54.9–58.9) | 56.5% (53.7–61.2) | 45.9% (43.3–49.5) |
| *Routine Only*<br><br>Medium coverage, 50% efficacy, lifelong protection | Averted cases before 2050 | 13.4m (11.6–15.5) | 5.3m (4.6–6.1) | 0.06m (0.05–0.08) | 1.4m (1.1–1.8) | 0.04m (0.03–0.05) | 5.0m (4.0–6.5) | 1.6m (1.2–1.9) | 12.4m (10.7–14.4) | 1.1m (0.9–1.2) | 1.7m (1.4–2.0) | 10.9m (9.4–12.7) | 0.9m (0.7–1.1) |
| | Averted tx before 2050 | 6.2m (5.6–6.9) | 1.9m (1.7–2.1) | 0.04m (0.03–0.05) | 0.8m (0.6–1.0) | 0.02m (0.02–0.03) | 2.6m (2.1–3.2) | 0.9m (0.7–1.1) | 5.7m (5.1–6.4) | 0.5m (0.4–0.6) | 0.8m (0.7–1.0) | 4.9m (4.4–5.6) | 0.4m (0.4–0.5) |
| | Averted deaths before 2050 | 1.5m (1.3–1.7) | 0.8m (0.6–0.9) | 0.00m (0.00–0.01) | 0.11m (0.08–0.15) | 0.003m (0.003–0.004) | 0.5m (0.4–0.7) | 0.08m (0.07–0.11) | 1.4m (1.2–1.6) | 0.1m (0.1–0.1) | 0.2m (0.1–0.2) | 1.3m (1.1–1.5) | 0.08m (0.06–0.12) |

| | | | | | | | | | | | | | |
|---|---|---|---|---|---|---|---|---|---|---|---|---|---|
| | IRR in 2050 (%) | 16.8% (15.3–19.2) | 19.2% (18.0–24.0) | 6.5% (5.8–7.4) | 20.4% (17.2–25.9) | 6.8% (5.8–8.3) | 15.1% (13.0–18.0) | 13.5% (11.6–16.5) | 17.2% (15.6–19.8) | 13.3% (12.2–15.1) | 17.6% (16.2–19.6) | 17.5% (15.7–20.4) | 9.9% (8.5–11.7) |
| | MRR in 2050 (%) | 15.8% (14.2–19.2) | 17.3% (15.9–23.7) | 6.3% (5.7–7.1) | 19.3% (16.2–24.0) | 5.9% (5.2–6.9) | 13.7% (11.7–16.5) | 14.4% (12.0–17.6) | 16.3% (14.7–20.0) | 11.3% (10.2–12.7) | 15.2% (14.0–16.8) | 16.3% (14.5–20.3) | 10.9% (9.1–13.5) |
| **75% efficacy vaccine** | | | | | | | | | | | | | |
| *Basecase* Medium coverage, 75% efficacy, 10 years protection | Averted cases before 2050 | 64.3m (54.0–75.6) | 20.3m (17.0–24.6) | 0.8m (0.7–0.9) | 5.6m (4.5–7.0) | 0.5m (0.4–0.6) | 28.4m (23.2–33.5) | 8.7m (7.3–10.3) | 58.2m (49.0–68.5) | 6.1m (5.0–7.2) | 7.3m (6.0–8.9) | 49.8m (41.4–58.9) | 7.1m (6.2–8.1) |
| | Averted tx before 2050 | 36.4m (32.0–40.1) | 9.2m (8.3–10.0) | 0.6m (0.5–0.6) | 3.4m (2.9–4.0) | 0.3m (0.3–0.4) | 17.1m (14.7–19.6) | 5.6m (4.9–6.3) | 33.0m (29.0–36.4) | 3.4m (2.9–3.8) | 4.2m (3.7–4.7) | 27.7m (24.1–30.9) | 4.4m (4.0–4.8) |
| | Averted deaths before 2050 | 7.3m (6.7–7.9) | 3.1m (2.8–3.4) | 0.1m (0.0–0.1) | 0.4m (0.4–0.6) | 0.04m (0.04–0.05) | 3.3m (2.9–3.7) | 0.04m (0.04–0.05) | 6.6m (6.1–7.2) | 0.7m (0.6–0.7) | 0.8m (0.7–0.9) | 5.9m (5.4–6.4) | 0.6m (0.5–0.7) |
| | IRR in 2050 (%) | 36.4% (34.5–39.1) | 38.7% (37.1–44.0) | 23.4% (22.4–24.7) | 37.7% (34.1–43.8) | 29.1% (27.1–32.1) | 36.2% (33.6–39.7) | 29.1% (27.2–32.1) | 36.4% (34.4–39.4) | 36.0% (34.9–37.8) | 39.0% (37.4–41.2) | 37.3% (35.0–40.7) | 24.8% (23.7–26.6) |
| | MRR in 2050 (%) | 38.8% (36.9–42.5) | 39.8% (38.2–46.6) | 26.0% (24.8–27.3) | 39.7% (35.9–45.4) | 28.9% (27.3–31.1) | 37.9% (35.1–41.4) | 33.7% (31.3–37.1) | 39.0% (36.9–43.1) | 37.4% (36.3–38.9) | 39.9% (38.5–41.9) | 39.5% (37.2–43.8) | 28.9% (27.2–31.6) |
| *Accelerated Scale–up* Medium coverage, 75% efficacy, 10 years protection | Averted cases before 2050 | 95.0m (80.5–110.9) | 28.2m (23.9–33.9) | 1.2m (1.1–1.4) | 7.8m (6.2–9.6) | 0.9m (0.8–1.1) | 44.8m (37.1–52.5) | 12.0m (10.2–14.1) | 84.8m (71.8–98.9) | 10.1m (8.4–12.0) | 10.9m (8.9–13.0) | 74.5m (62.6–87.3) | 9.6m (8.4–10.9) |
| | Averted tx before 2050 | 56.0m (49.7–61.6) | 13.4m (12.2–14.5) | 0.9m (0.8–1.0) | 4.9m (4.1–5.7) | 0.7m (0.6–0.7) | 28.2m (24.3–32.1) | 7.9m (7.0–8.8) | 50.2m (44.5–55.3) | 5.8m (5.1–6.4) | 6.5m (5.7–7.2) | 43.4m (38.2–48.2) | 6.1m (5.6–6.6) |
| | Averted deaths before 2050 | 11.4m (10.5–12.3) | 4.5m (4.1–5.0) | 0.09m (0.08–0.10) | 0.6m (0.5–0.8) | 0.08m (0.07–0.09) | 5.4m (4.8–6.2) | 0.6m (0.5–0.7) | 10.1m (9.3–11.0) | 1.2m (1.1–1.4) | 1.3m (1.2–1.5) | 9.3m (8.5–10.1) | 0.8m (0.6–1.0) |
| | IRR in 2050 (%) | 35.8% (34.2–38.5) | 39.2% (37.6–44.6) | 22.0% (20.8–23.2) | 37.9% (34.6–43.5) | 25.6% (23.3–29.6) | 34.9% (32.5–38.2) | 28.2% (26.6–30.7) | 35.9% (34.1–38.7) | 35.8% (34.7–37.4) | 38.9% (37.5–40.9) | 36.7% (34.7–40.0) | 24.1% (23.1–25.3) |
| | MRR in 2050 (%) | 37.8% (36.0–41.7) | 40.1% (38.4–47.7) | 23.4% (22.1–24.9) | 39.1% (35.6–44.5) | 25.5% (23.6–28.6) | 35.8% (33.2–39.3) | 31.6% (29.5–34.8) | 37.9% (35.9–42.3) | 36.9% (36.0–38.3) | 39.2% (37.9–41.1) | 38.5% (36.3–43.1) | 27.3% (25.9–29.4) |

*Abbreviations: AFR = WHO African Region, AMR = WHO Region of the Americas, EMR = WHO Eastern Mediterranean Region, EUR = WHO European Region, HBC = high burden countries, IRR = incidence rate reduction, LIC = low-income countries, LMIC = lower-middle income countries, MRR = mortality rate reduction, SEAR = WHO South-East Asian Region, tx = treatments, UMIC = upper-middle income countries, WPR = WHO Western Pacific Region*

**Table S10.2** Estimated health impact in 2050 by WHO region, income level, and tuberculosis burden level for the infant vaccine scenarios

| | Health Impact Measure | All modelled countries | WHO Region | | | | | | TB Burden Level | | World Bank Income Group | | |
|---|---|---|---|---|---|---|---|---|---|---|---|---|---|
| **Vaccine Scenario** | | | **AFR** | **AMR** | **EMR** | **EUR** | **SEAR** | **WPR** | **HBC** | **All other countries** | **LIC** | **LMIC** | **UMIC** |
| *Basecase* Medium coverage, 80% efficacy, 10 years protection | Averted cases before 2050 | 6.7m (5.8–7.7) | 2.9m (2.5–3.4) | 0.03m (0.02–0.03) | 0.8m (0.6–1.1) | 0.02m (0.01–0.02) | 2.2m (1.6–2.8) | 0.8m (0.6–1.0) | 6.2m (5.3–7.1) | 0.5m (0.4–0.6) | 0.9m (0.7–1.1) | 5.4m (4.7–6.2) | 0.4m (0.3–0.5) |
| | Averted tx before 2050 | 2.7m (2.4–2.9) | 0.9m (0.8–0.9) | 0.02m (0.01–0.02) | 0.4m (0.3–0.5) | 0.009m (0.008–0.01) | 1.0m (0.8–1.2) | 0.4m (0.3–0.5) | 2.4m (2.2–2.7) | 0.2m (0.2–0.3) | 0.4m (0.4–0.5) | 2.1m (1.9–2.3) | 0.2m (0.1–0.2) |
| | Averted deaths before 2050 | 0.9m (0.8–1.0) | 0.5m (0.4–0.6) | 0.003m (0.002–0.003) | 0.08m (0.05–0.11) | 0.0018m (0.0015–0.0021) | 0.3m (0.2–0.4) | 0.1m (0.0–0.1) | 0.8m (0.7–1.0) | 0.07m (0.06–0.08) | 0.11m (0.09–0.13) | 0.7m (0.6–0.9) | 0.1m (0.0–0.1) |
| | IRR in 2050 (%) | 8.8% (7.9–10.4) | 11.0% (10.0–14.5) | 2.7% (2.4–3.1) | 12.0% (9.7–15.6) | 2.9% (2.5–3.4) | 6.9% (5.8–8.6) | 7.2% (5.9–9.2) | 9.0% (8.1–10.7) | 7.1% (6.4–8.2) | 9.8% (8.9–11.1) | 9.1% (8.1–11.1) | 4.7% (3.9–5.9) |
| | MRR in 2050 (%) | 9.8% (8.7–12.0) | 11.3% (10.1–15.7) | 3.7% (3.2–4.3) | 13.4% (10.5–18.1) | 3.3% (2.9–3.9) | 7.2% (5.9–9.6) | 11.2% (8.5–15.4) | 10.1% (8.9–12.5) | 7.1% (6.4–8.0) | 9.9% (9.0–11.2) | 10.0% (8.7–12.5) | 6.6% (5.4–8.5) |
| *Accelerated Scale–up* Medium coverage, 80% efficacy, 10 years protection | Averted cases before 2050 | 16.3m (14.0–18.8) | 6.3m (5.4–7.2) | 0.07m (0.06–0.08) | 1.7m (1.3–2.2) | 0.07m (0.06–0.09) | 6.6m (5.1–8.6) | 1.5m (1.2–1.9) | 14.7m (12.6–17.1) | 1.6m (1.3–1.9) | 2.2m (1.8–2.8) | 13.3m (11.4–15.5) | 0.8m (0.6–0.9) |
| | Averted tx before 2050 | 7.7m (6.9–8.6) | 2.2m (2.0–2.4) | 0.04m (0.04–0.05) | 0.9m (0.8–1.2) | 0.04m (0.04–0.05) | 3.6m (2.9–4.3) | 0.9m (0.7–1.0) | 6.9m (6.2–7.8) | 0.7m (0.7–0.8) | 1.1m (1.0–1.3) | 6.2m (5.5–7.0) | 0.4m (0.3–0.4) |
| | Averted deaths before 2050 | 2.3m (2.0–2.6) | 1.1m (0.9–1.2) | 0.007m (0.006–0.008) | 0.2m (0.1–0.2) | 0.007m (0.006–0.009) | 0.9m (0.7–1.2) | 0.1m (0.1–0.2) | 2.0m (1.8–2.3) | 0.2m (0.2–0.3) | 0.3m (0.2–0.3) | 1.9m (1.6–2.2) | 0.1m (0.1–0.1) |
| | IRR in 2050 (%) | 14.3% (13.0–16.7) | 16.7% (15.4–21.6) | 4.6% (4.2–5.2) | 17.6% (14.5–22.3) | 7.5% (6.2–9.8) | 12.9% (11.0–15.8) | 10.3% (8.7–12.6) | 14.4% (13.0–17.0) | 13.4% (12.4–14.9) | 16.3% (15.1–18.1) | 14.9% (13.3–17.9) | 6.5% (5.6–7.8) |
| | MRR in 2050 (%) | 15.9% (14.2–19.3) | 17.5% (15.9–24.1) | 5.8% (5.2–6.6) | 19.2% (15.5–24.7) | 7.7% (6.5–9.5) | 13.4% (11.2–17.0) | 15.0% (12.0–19.3) | 16.1% (14.3–19.9) | 14.1% (13.1–15.3) | 16.8% (15.6–18.5) | 16.3% (14.4–20.3) | 8.9% (7.5–11.1) |
| **Low and high coverage** | | | | | | | | | | | | | |
| *Basecase* Low coverage, 80% efficacy, 10 years protection | Averted cases before 2050 | 6.0m (5.2–6.8) | 2.6m (2.2–3.0) | 0.02m (0.02–0.03) | 0.7m (0.5–1.0) | 0.01m (0.01–0.02) | 1.9m (1.5–2.5) | 0.7m (0.5–0.9) | 5.5m (4.7–6.3) | 0.5m (0.4–0.6) | 0.8m (0.6–1.0) | 4.8m (4.2–5.5) | 0.4m (0.3–0.5) |
| | Averted tx before 2050 | 2.4m (2.2–2.6) | 0.8m (0.7–0.8) | 0.01m (0.01–0.02) | 0.4m (0.3–0.5) | 0.008m (0.007–0.009) | 0.9m (0.7–1.0) | 0.3m (0.3–0.4) | 2.2m (2.0–2.4) | 0.20m (0.18–0.23) | 0.4m (0.3–0.4) | 1.8m (1.7–2.1) | 0.2m (0.1–0.2) |
| | Averted deaths before 2050 | 0.8m (0.7–0.9) | 0.4m (0.4–0.5) | 0.003m (0.002–0.003) | 0.1m (0.0–0.1) | 0.002m (0.001–0.002) | 0.2m (0.2–0.3) | 0.1m (0.0–0.1) | 0.7m (0.6–0.8) | 0.06m (0.05–0.07) | 0.09m (0.08–0.12) | 0.7m (0.6–0.8) | 0.0m (0.0–0.1) |

| | | | | | | | | | | | | | |
|---|---|---|---|---|---|---|---|---|---|---|---|---|---|
| | IRR in 2050 (%) | 7.9% (7.1–9.3) | 9.8% (9.0–13.1) | 2.4% (2.2–2.8) | 10.8% (8.6–14.0) | 2.6% (2.3–3.0) | 6.2% (5.2–7.7) | 6.5% (5.3–8.2) | 8.1% (7.2–9.6) | 6.3% (5.7–7.3) | 8.7% (7.9–9.9) | 8.2% (7.2–9.9) | 4.2% (3.5–5.2) |
| | MRR in 2050 (%) | 8.7% (7.8–10.7) | 10.1% (9.0–14.1) | 3.3% (2.9–3.9) | 12.0% (9.4–16.2) | 2.9% (2.5–3.4) | 6.4% (5.2–8.6) | 10.0% (7.6–13.8) | 9.0% (8.0–11.2) | 6.3% (5.7–7.2) | 8.8% (8.0–10.0) | 8.9% (7.7–11.2) | 5.9% (4.8–7.5) |
| *Basecase* High coverage, 80% efficacy, 10 years protection | Averted cases before 2050 | 7.4m (6.5–8.6) | 3.3m (2.8–3.7) | 0.03m (0.03–0.04) | 0.9m (0.7–1.2) | 0.018m (0.015–0.022) | 2.4m (1.8–3.1) | 0.9m (0.7–1.1) | 6.9m (5.9–7.9) | 0.6m (0.5–0.7) | 1.0m (0.8–1.2) | 6.0m (5.2–6.9) | 0.5m (0.4–0.6) |
| | Averted tx before 2050 | 3.0m (2.7–3.3) | 1.0m (0.9–1.0) | 0.017m (0.015–0.019) | 0.5m (0.4–0.6) | 0.010m (0.009–0.011) | 1.1m (0.9–1.3) | 0.4m (0.4–0.5) | 2.7m (2.5–3.0) | 0.3m (0.2–0.3) | 0.5m (0.4–0.5) | 2.3m (2.1–2.6) | 0.2m (0.2–0.2) |
| | Averted deaths before 2050 | 1.0m (0.9–1.1) | 0.5m (0.4–0.6) | 0.003m (0.003–0.004) | 0.08m (0.06–0.12) | 0.0020m (0.0017–0.0024) | 0.3m (0.2–0.4) | 0.1m (0.0–0.1) | 0.9m (0.8–1.1) | 0.08m (0.06–0.09) | 0.12m (0.10–0.14) | 0.8m (0.7–1.0) | 0.1m (0.0–0.1) |
| | IRR in 2050 (%) | 9.7% (8.8–11.4) | 12.1% (11.1–16.0) | 3.0% (2.7–3.5) | 13.3% (10.7–17.1) | 3.2% (2.8–3.7) | 7.7% (6.4–9.5) | 8.0% (6.5–10.1) | 9.9% (8.9–11.8) | 7.8% (7.0–9.1) | 10.8% (9.8–12.2) | 10.1% (8.9–12.2) | 5.2% (4.4–6.5) |
| | MRR in 2050 (%) | 10.8% (9.6–13.2) | 12.5% (11.2–17.3) | 4.1% (3.6–4.8) | 14.8% (11.6–19.9) | 3.7% (3.2–4.3) | 8.0% (6.5–10.6) | 12.3% (9.4–16.9) | 11.2% (9.9–13.8) | 7.9% (7.2–8.9) | 10.9% (9.9–12.3) | 11.1% (9.6–13.8) | 7.4% (6.0–9.3) |
| *Accelerated Scale–up* Low coverage, 80% efficacy, 10 years protection | Averted cases before 2050 | 14.6m (12.6–16.8) | 5.6m (4.8–6.4) | 0.06m (0.05–0.07) | 1.5m (1.1–2.0) | 0.06m (0.05–0.08) | 5.9m (4.6–7.7) | 1.4m (1.1–1.7) | 13.1m (11.3–15.3) | 1.4m (1.2–1.7) | 2.0m (1.6–2.5) | 11.9m (10.2–13.9) | 0.7m (0.5–0.8) |
| | Averted tx before 2050 | 6.9m (6.2–7.7) | 2.0m (1.8–2.2) | 0.04m (0.03–0.04) | 0.8m (0.7–1.1) | 0.04m (0.03–0.05) | 3.2m (2.6–3.9) | 0.8m (0.6–0.9) | 6.2m (5.5–7.0) | 0.7m (0.6–0.7) | 1.0m (0.9–1.2) | 5.5m (4.9–6.3) | 0.3m (0.3–0.4) |
| | Averted deaths before 2050 | 2.0m (1.8–2.3) | 1.0m (0.8–1.1) | 0.006m (0.005–0.008) | 0.1m (0.1–0.2) | 0.006m (0.005–0.008) | 0.8m (0.6–1.0) | 0.1m (0.1–0.2) | 1.8m (1.6–2.1) | 0.20m (0.17–0.22) | 0.3m (0.2–0.3) | 1.7m (1.4–1.9) | 0.1m (0.1–0.1) |
| | IRR in 2050 (%) | 12.9% (11.7–15.1) | 15.0% (13.9–19.5) | 4.1% (3.8–4.7) | 15.8% (13.0–20.1) | 6.7% (5.5–8.8) | 11.6% (9.9–14.2) | 9.2% (7.8–11.3) | 13.0% (11.6–15.4) | 12.0% (11.1–13.5) | 14.6% (13.5–16.3) | 13.4% (11.9–16.1) | 5.8% (5.0–7.0) |
| | MRR in 2050 (%) | 14.3% (12.8–17.4) | 15.7% (14.2–21.8) | 5.2% (4.7–5.9) | 17.3% (13.9–22.3) | 6.9% (5.8–8.5) | 12.1% (10.1–15.4) | 13.5% (10.8–17.4) | 14.5% (12.8–18.0) | 12.6% (11.7–13.8) | 15.1% (14.0–16.7) | 14.6% (12.9–18.3) | 8.0% (6.7–9.9) |
| *Accelerated Scale–up* High coverage, 80% efficacy, 10 years protection | Averted cases before 2050 | 18.0m (15.5–20.7) | 6.9m (5.9–7.9) | 0.08m (0.07–0.09) | 1.9m (1.4–2.4) | 0.08m (0.06–0.10) | 7.3m (5.7–9.4) | 1.7m (1.3–2.1) | 16.2m (13.9–18.9) | 1.8m (1.5–2.1) | 2.5m (2.0–3.1) | 14.6m (12.5–17.1) | 0.9m (0.7–1.0) |
| | Averted tx before 2050 | 8.5m (7.6–9.5) | 2.5m (2.2–2.7) | 0.05m (0.04–0.05) | 1.0m (0.8–1.3) | 0.05m (0.04–0.06) | 3.9m (3.3–4.8) | 0.9m (0.8–1.1) | 7.7m (6.9–8.6) | 0.8m (0.7–0.9) | 1.3m (1.1–1.4) | 6.8m (6.1–7.8) | 0.4m (0.3–0.5) |
| | Averted deaths before 2050 | 2.5m (2.2–2.8) | 1.2m (1.0–1.4) | 0.008m (0.007–0.009) | 0.2m (0.1–0.2) | 0.008m (0.007–0.010) | 1.0m (0.7–1.3) | 0.1m (0.1–0.2) | 2.2m (1.9–2.6) | 0.2m (0.2–0.3) | 0.3m (0.3–0.4) | 2.1m (1.8–2.4) | 0.1m (0.1–0.1) |
| | IRR in 2050 (%) | 15.7% (14.3–18.3) | 18.3% (17.0–23.5) | 5.1% (4.7–5.7) | 19.3% (15.9–24.3) | 8.2% (6.8–10.7) | 14.1% (12.1–17.3) | 11.3% (9.6–13.8) | 15.8% (14.2–18.6) | 14.7% (13.6–16.3) | 17.8% (16.5–19.8) | 16.3% (14.6–19.5) | 7.2% (6.1–8.6) |

| | | | | | | | | | | | | | |
|---|---|---|---|---|---|---|---|---|---|---|---|---|---|
| | MRR in 2050 (%) | 17.4% (15.7–21.1) | 19.2% (17.5–26.2) | 6.4% (5.8–7.3) | 21.0% (17.1–27.0) | 8.4% (7.2–10.4) | 14.7% (12.4–18.6) | 16.5% (13.3–21.1) | 17.7% (15.7–21.7) | 15.5% (14.4–16.8) | 18.5% (17.1–20.3) | 17.9% (15.8–22.2) | 9.8% (8.3–12.2) |
| **Lifelong protection** | | | | | | | | | | | | | |
| *Basecase*<br>Medium coverage, 80% efficacy, lifelong protection | Averted cases before 2050 | 11.8m (10.2–13.7) | 5.2m (4.4–6.0) | 0.1m (0.0–0.1) | 1.4m (1.1–1.8) | 0.029m (0.025–0.034)c | 3.8m (2.9–4.9) | 1.4m (1.1–1.8) | 10.9m (9.4–12.6) | 0.9m (0.8–1.1) | 1.5m (1.2–1.9) | 9.5m (8.2–11.0) | 0.8m (0.6–1.0) |
| | Averted tx before 2050 | 4.6m (4.2–5.0) | 1.5m (1.3–1.6) | 0.029m (0.025–0.032) | 0.7m (0.6–0.9) | 0.02m (0.01–0.02) | 1.7m (1.4–2.0) | 0.7m (0.6–0.8) | 4.2m (3.8–4.6) | 0.4m (0.3–0.4) | 0.7m (0.6–0.8) | 3.6m (3.2–4.0) | 0.3m (0.3–0.4) |
| | Averted deaths before 2050 | 1.5m (1.3–1.7) | 0.8m (0.7–0.9) | 0.01m (0.00–0.01) | 0.1m (0.1–0.2) | 0.003m (0.003–0.004) | 0.5m (0.3–0.6) | 0.1m (0.1–0.2) | 1.4m (1.2–1.6) | 0.11m (0.09–0.13) | 0.2m (0.1–0.2) | 1.2m (1.1–1.4) | 0.1m (0.1–0.1) |
| | IRR in 2050 (%) | 17.4% (15.9–20.1) | 21.6% (20.0–27.5) | 6.1% (5.5–7.0) | 23.4% (19.2–29.6) | 5.8% (5.2–6.6) | 13.7% (11.7–16.7) | 15.1% (12.5–18.8) | 17.8% (16.2–20.9) | 13.6% (12.3–15.6) | 18.8% (17.2–21.2) | 18.0% (16.1–21.4) | 10.5% (8.8–12.9) |
| | MRR in 2050 (%) | 18.1% (16.2–21.7) | 20.9% (18.9–28.2) | 7.5% (6.5–8.6) | 24.6% (19.7–32.0) | 6.4% (5.6–7.3) | 13.5% (11.1–17.5) | 20.8% (16.1–27.7) | 18.7% (16.7–22.7) | 12.8% (11.6–14.4) | 17.9% (16.4–20.1) | 18.5% (16.2–22.7) | 13.4% (11.1–16.7) |
| *Accelerated Scale–up*<br>Medium coverage, 80% efficacy, lifelong protection | Averted cases before 2050 | 32.2m (27.5–37.5) | 12.0m (10.4–14.1) | 0.1m (0.1–0.2) | 3.2m (2.4–4.2) | 0.1m (0.1–0.2) | 13.5m (10.6–17.0) | 3.1m (2.5–3.9) | 29.1m (24.6–34.0) | 3.2m (2.6–3.7) | 4.4m (3.6–5.4) | 26.2m (22.3–30.9) | 1.6m (1.3–2.0) |
| | Averted tx before 2050 | 15.3m (13.7–17.1) | 4.4m (4.0–4.7) | 0.09m (0.08–0.10) | 1.8m (1.4–2.2) | 0.09m (0.07–0.10) | 7.2m (6.0–8.7) | 1.7m (1.4–2.0) | 13.8m (12.3–15.6) | 1.5m (1.3–1.6) | 2.2m (1.9–2.5) | 12.3m (10.9–14.0) | 0.8m (0.7–0.9) |
| | Averted deaths before 2050 | 4.2m (3.7–4.8) | 2.0m (1.7–2.2) | 0.01m (0.01–0.02) | 0.3m (0.2–0.4) | 0.01m (0.01–0.02) | 1.7m (1.3–2.2) | 0.2m (0.2–0.3) | 3.8m (3.3–4.3) | 0.4m (0.4–0.5) | 0.5m (0.5–0.6) | 3.5m (3.0–4.0) | 0.2m (0.1–0.3) |
| | IRR in 2050 (%) | 31.8% (29.4–35.5) | 36.2% (34.2–43.3) | 11.8% (10.9–13.0) | 37.1% (31.7–45.4) | 17.1% (14.6–21.3) | 29.6% (26.1–34.7) | 23.7% (20.4–28.1) | 32.1% (29.5–36.2) | 29.5% (27.9–32.0) | 35.5% (33.4–38.5) | 33.1% (30.1–37.7) | 15.9% (13.7–18.8) |
| | MRR in 2050 (%) | 33.0% (30.2–38.3) | 35.5% (33.0–45.5) | 13.4% (12.2–14.9) | 38.4% (32.2–47.3) | 16.7% (14.6–19.8) | 29.2% (25.3–35.1) | 30.5% (25.3–37.4) | 33.5% (30.3–39.3) | 29.3% (27.7–31.4) | 34.7% (32.7–37.4) | 33.7% (30.4–40.1) | 19.8% (16.9–24.0) |

*Abbreviations: AFR = WHO African Region, AMR = WHO Region of the Americas, EMR = WHO Eastern Mediterranean Region, EUR = WHO European Region, HBC = high burden countries, IRR = incidence rate reduction, LIC = low-income countries, LMIC = lower-middle income countries, MRR = mortality rate reduction, SEAR = WHO South-East Asian Region, tx = treatments, UMIC = upper-middle income countries, WPR = WHO Western Pacific Region*

### 10.2 Comparing to the 2035 End TB incidence target

We calculated the incidence rate in 2035 for each *No-New-Vaccine* baseline and for each vaccine scenario to compare with the 2035 End TB incidence target of a 90% reduction in the tuberculosis incidence rate compared to the 2015 incidence rate. Results for all modelled countries, and for each of the select model groupings for outcome reporting are provided in Table S10.3, as the median estimate of the incidence rate per 100,000 population and 95% uncertainty range, with all vaccine scenarios assuming medium coverage and 10-years protection.

For all modelled countries, the estimated incidence rate in 2015 was approximately 164.2 per 100,000 population. A 90% reduction is equivalent to an incidence rate of 16.4 per 100,000 population. With the *Status Quo No-New-Vaccine* baseline, the closest vaccine scenario to reaching this target reduction is the *Accelerated Scale-up* scenario of an adolescent/adult vaccine with vaccine efficacy increased to 75%, which has an estimated incidence rate of 88.1 (70.9–104.7) per 100,000 population or meeting approximately 52% of the goal. With the *2025 End TB No-New-Vaccine* baseline, progress is increased, with the standard *Basecase* scenario of the adolescent/adult vaccine achieving an incidence rate of 42.5 (37.1–50.6) per 100,000 population and the standard *Accelerated Scale-up* scenario achieving an incidence rate of 43.4 (37.7–51.6) per 100,000 population, or 82% of the target.

**Table S10.3** Estimated incidence rate (per 100,000 population) for each vaccine scenario in 2035 to compare to meeting the 2035 End TB target.

| | All modelled countries | WHO Region | | | | | | TB Burden Level | | World Bank Income Group | | |
|---|---|---|---|---|---|---|---|---|---|---|---|---|
| **Scenario** | | **AFR** | **AMR** | **EMR** | **EUR** | **SEAR** | **WPR** | **HBC** | **All other countries** | **LIC** | **LMIC** | **UMIC** |
| *Status Quo No-New-Vaccine* baseline | 138.8 (114.2–163.6) | 196.8 (154.9–240.4) | 26.2 (22.3–30.0) | 94.8 (76.2–117.4) | 31.4 (26.8–36.4) | 213.2 (179.1–245.2) | 85.3 (70.6–101.5) | 169.9 (139.7–200.0) | 55.9 (46.0–65.9) | 132.3 (108.2–158.7) | 199.6 (163.1–236.4) | 50.6 (43.6–57.5) |
| Adolescent/adult vaccine: *Basecase*, 50% efficacy | 113.5 (93.1–133.9) | 155.4 (121.6–191.1) | 20.6 (17.4–23.7) | 70.9 (56.8–88.3) | 26.0 (22.1–30.3) | 183.9 (154.4–211.4) | 65.2 (53.5–78.3) | 138.0 (113.1–162.5) | 48.3 (39.6–57.4) | 110.6 (90.2–133.0) | 165.0 (134.6–195.3) | 38.1 (32.6–43.6) |
| Adolescent/adult vaccine: *Accelerated Scale-up*, 50% efficacy | 103.8 (84.1–123.0) | 145.4 (110.6–179.3) | 21.2 (18.0–24.4) | 69.3 (54.9–86.7) | 25.0 (21.3–29.3) | 157.2 (130.8–180.5) | 67.2 (54.8–80.4) | 126.9 (102.8–150.2) | 42.3 (34.7–50.6) | 96.9 (78.3–116.9) | 147.6 (118.4–175.3) | 41.0 (35.0–46.8) |
| Adolescent/adult vaccine: *Routine Only*, 50% efficacy | 137.3 (112.9–162.0) | 193.6 (151.8–236.6) | 26.1 (22.2–29.8) | 93.2 (74.8–115.4) | 31.3 (26.7–36.3) | 212.2 (178.3–243.8) | 84.1 (69.6–100.4) | 168.1 (138.0–197.9) | 55.5 (45.7–65.5) | 130.8 (107.0–156.8) | 197.5 (161.3–233.9) | 50.1 (43.2–56.9) |
| Adolescent/adult vaccine: *Basecase*, 75% efficacy | 100.9 (82.7–119.1) | 135.1 (105.4–166.1) | 17.8 (15.0–20.5) | 59.3 (47.4–74.1) | 23.2 (19.7–27.2) | 169.1 (141.8–194.1) | 55.3 (45.1–66.8) | 122.2 (100.0–143.6) | 44.5 (36.4–53.1) | 99.9 (81.3–120.5) | 148.0 (120.7–174.7) | 31.8 (26.9–36.4) |

| | | | | | | | | | | | | |
|---|---|---|---|---|---|---|---|---|---|---|---|---|
| Adolescent/adult vaccine: *Accelerated Scale-up*, 75% efficacy | 88.1 (70.9–104.7) | 122.0 (91.2–150.7) | 18.8 (15.9–21.7) | 58.4 (45.7–72.9) | 22.1 (18.7–25.8) | 132.4 (110.2–153.1) | 58.4 (47.4–70.0) | 107.5 (86.4–127.8) | 36.1 (29.6–43.4) | 80.9 (65.1–98.1) | 124.3 (99.0–148.4) | 36.2 (30.8–41.3) |
| Infant vaccine: *Basecase*, 80% efficacy | 138.0 (113.5–162.7) | 194.8 (153.1–237.9) | 26.1 (22.2–29.9) | 93.8 (75.3–116.2) | 31.3 (26.8–36.4) | 212.8 (178.8–244.6) | 84.6 (70.1–100.9) | 169.0 (138.8–198.9) | 55.7 (45.8–65.7) | 131.5 (107.5–157.7) | 198.5 (162.2–235.1) | 50.3 (43.4–57.2) |
| Infant vaccine: *Accelerated Scale-up*, 80% efficacy | 133.1 (109.1–157.0) | 186.6 (145.6–228.0) | 25.8 (22.0–29.6) | 89.0 (71.5–110.6) | 30.8 (26.3–35.8) | 205.8 (172.8–236.1) | 82.8 (68.6–98.9) | 163.0 (133.4–192.0) | 53.8 (44.2–63.7) | 125.3 (102.3–150.1) | 191.0 (155.9–226.0) | 49.7 (42.9–56.5) |
| *2025 End TB No-New-Vaccine* baseline | 51.5 (45.0–61.2) | 70.2 (58.8–85.5) | 12.3 (11.1–13.4) | 36.4 (28.3–58.2) | 17.4 (14.7–21.2) | 73.0 (56.8-100.1) | 37.1 (30.2–49.2) | 62.0 (53.2–75.2) | 23.3 (20.7–26.2) | 46.9 (40.3–56.1) | 69.0 (57.5–87.5) | 26.4 (23.5–31.5) |
| Adolescent/adult vaccine: *Basecase*, 50% efficacy | 42.5 (37.1–50.6) | 56.6 (47.5–68.0) | 9.8 ( 8.9–10.7) | 28.0 (21.9–44.0) | 14.3 (12.2–17.2) | 63.1 (49.3–85.9) | 29.3 (24.0–38.2) | 50.6 (43.4–61.8) | 20.4 (18.1–23.0) | 39.7 (34.0–46.7) | 57.7 (48.4–73.2) | 20.3 (18.1–24.2) |
| Adolescent/adult vaccine: *Accelerated Scale-up*, 50% efficacy | 43.4 (37.7–51.6) | 58.5 (48.5–70.9) | 10.6 ( 9.6–11.6) | 30.5 (23.7–45.7) | 15.0 (12.6–18.3) | 61.7 (47.9–83.2) | 31.8 (26.0–41.4) | 52.2 (44.7–63.3) | 19.9 (17.7–22.2) | 39.4 (33.9–46.4) | 58.0 (48.2–72.8) | 22.7 (20.3–27.0) |

*Abbreviations: AFR = WHO African Region, AMR = WHO Region of the Americas, EMR = WHO Eastern Mediterranean Region, EUR = WHO European Region, HBC = high burden countries, IRR = incidence rate reduction, LIC = low-income countries, LMIC = lower-middle income countries, MRR = mortality rate reduction, SEAR = WHO South-East Asian Region, UMIC = upper-middle income countries, WPR = WHO Western Pacific Region*

3 World Health Organization. Guidelines for treatment of drug-susceptible tuberculosis and patient care: 2017 update. 2017.

4 World Health Organization. Treatment of drug-susceptible tuberculosis: rapid communication. Geneva: World Health Organization, 2021 https://apps.who.int/iris/handle/10665/341729 (accessed Jan 7, 2022).

5 Sutherland I, Svandová E, Radhakrishna S. The development of clinical tuberculosis following infection with tubercle bacilli. 1. A theoretical model for the development of clinical tuberculosis following infection, linking from data on the risk of tuberculous infection and the incidence of clinical tuberculosis in the Netherlands. *Tubercle* 1982; **63**: 255–68.

6 Vynnycky E, Fine PE. The natural history of tuberculosis: the implications of age-dependent risks of disease and the role of reinfection. *Epidemiol Infect* 1997; **119**: 183–201.

7 World Health Organization. Global tuberculosis report 2021. Geneva: World Health Organization, 2021 https://apps.who.int/iris/handle/10665/346387 (accessed Jan 7, 2022).

8 Lee JY. Diagnosis and Treatment of Extrapulmonary Tuberculosis. *Tuberc Respir Dis* 2015; **78**: 47–55.

9 Prem K, Zandvoort K van, Klepac P, *et al.* Projecting contact matrices in 177 geographical regions: An update and comparison with empirical data for the COVID-19 era. *PLOS Comput Biol* 2021; **17**: e1009098.

10 Kwan CK, Ernst JD. HIV and Tuberculosis: a Deadly Human Syndemic. *Clin Microbiol Rev* 2011; **24**: 351–76.

11 Suthar AB, Lawn SD, del Amo J, *et al.* Antiretroviral Therapy for Prevention of Tuberculosis in Adults with HIV: A Systematic Review and Meta-Analysis. *PLoS Med* 2012; **9**: e1001270.

12 Harling G, Ehrlich R, Myer L. The social epidemiology of tuberculosis in South Africa: A multilevel analysis. *Soc Sci Med* 2008; **66**: 492–505.

13 Hoa NB, Tiemersma EW, Sy DN, *et al.* Household expenditure and tuberculosis prevalence in Viet Nam: prediction by a set of household indicators. ; : 7.

14 Hossain S, Quaiyum MA, Zaman K, *et al.* Socio Economic Position in TB Prevalence and Access to Services: Results from a Population Prevalence Survey and a Facility-Based Survey in Bangladesh. *PLOS ONE* 2012; **7**: e44980.

15 Kapata N, Chanda-Kapata P, Ngosa W, *et al.* The Prevalence of Tuberculosis in Zambia: Results from the First National TB Prevalence Survey, 2013–2014. *PLOS ONE* 2016; **11**: e0146392.

16 Migambi P, Gasana M, Uwizeye CB, *et al.* Prevalence of tuberculosis in Rwanda: Results of the first nationwide survey in 2012 yielded important lessons for TB control. *PLOS ONE* 2020; **15**: e0231372.

17 Oxlade O, Murray M. Tuberculosis and Poverty: Why Are the Poor at Greater Risk in India? *PLOS ONE* 2012; **7**: e47533.

18 Singh SK, Kashyap GC, Puri P. Potential effect of household environment on prevalence of tuberculosis in India: evidence from the recent round of a cross-sectional survey. *BMC Pulm Med* 2018; **18**: 66.

19 Siroka A, Law I, Macinko J, *et al.* The effect of household poverty on tuberculosis. *Int J Tuberc Lung Dis Off J Int Union Tuberc Lung Dis* 2016; **20**: 1603–8.

20 van Leth F, Guilatco RS, Hossain S, *et al.* Measuring socio-economic data in tuberculosis prevalence surveys. *Int J Tuberc Lung Dis Off J Int Union Tuberc Lung Dis* 2011; **15 Suppl 2**: 58–63.

21 Yates TA, Ayles H, Leacy FP, *et al.* Socio-economic gradients in prevalent tuberculosis in Zambia and the Western Cape of South Africa. *Trop Med Int Health* 2018; **23**: 375–90.

22 United Nations Department of Economic and Social Affairs. UN Population Projections, 2019 Revision. https://population.un.org/wpp/Download/Standard/Population/.

23 Tiemersma EW, Werf MJ van der, Borgdorff MW, Williams BG, Nagelkerke NJD. Natural History of Tuberculosis: Duration and Fatality of Untreated Pulmonary Tuberculosis in HIV Negative Patients: A Systematic Review. *PLOS ONE* 2011; **6**: e17601.

24 Quaife M, Houben RMGJ, Allwood B, *et al.* Post-tuberculosis mortality and morbidity: valuing the hidden epidemic. *Lancet Respir Med* 2020; **8**: 332–3.

25 World Health Organization. WHO TB Burden Estimates. 2021; published online Dec 17. https://www.who.int/tb/country/data/download/en/.

26 World Health Organization. WHO Case Notifications. 2020. https://www.who.int/tb/country/data/download/en/.

27 Emery J. Subclinical TB disease: lessons and questions from field to lab. 2020.

28 Abu-Raddad L, Sabatelli L, Achterberg JT, *et al.* Epidemiological benefits of more-effective tuberculosis vaccines, drugs, and diagnostics. *Proc Natl Acad Sci U S A* 2009; **106**: 13980–5.

29 Dye C, Williams BG. Eliminating human tuberculosis in the twenty-first century. *J R Soc Interface* 2008; **5**: 653–62.

30 Marx FM, Dunbar R, Enarson DA, *et al.* The Temporal Dynamics of Relapse and Reinfection Tuberculosis After Successful Treatment: A Retrospective Cohort Study. *Clin Infect Dis* 2014; **58**: 1676–83.

31 Gomes MGM, Franco AO, Gomes MC, Medley GF. The reinfection threshold promotes variability in tuberculosis epidemiology and vaccine efficacy. *Proc R Soc B Biol Sci* 2004; **271**: 617–23.

32 Dangisso MH, Woldesemayat EM, Datiko DG, Lindtjørn B. Long-term outcome of smear-positive tuberculosis patients after initiation and completion of treatment: A ten-year retrospective cohort study. *PloS One* 2018; **13**: e0193396.

33 World Health Organization. WHO Treatment Outcomes. 2020. https://www.who.int/tb/country/data/download/en/.

34 Gabriela M. Gomes M, Rodrigues P, Hilker FM, *et al.* Implications of partial immunity on the prospects for tuberculosis control by post-exposure interventions. *J Theor Biol* 2007; **248**: 608–17.

35 Mberi MN, Kuonza LR, Dube NM, Nattey C, Manda S, Summers R. Determinants of loss to follow-up in patients on antiretroviral treatment, South Africa, 2004–2012: a cohort study. *BMC Health Serv Res* 2015; **15**: 259.

36 Cutsem GV, Ford N, Hildebrand K, *et al.* Correcting for Mortality Among Patients Lost to Follow Up on Antiretroviral Therapy in South Africa: A Cohort Analysis. *PLOS ONE* 2011; **6**: e14684.

37 Johansson KA, Robberstad B, Norheim OF. Further benefits by early start of HIV treatment in low income countries: Survival estimates of early versus deferred antiretroviral therapy. *AIDS Res Ther* 2010; **7**: 3.

38 Fatti G, Mothibi E, Meintjes G, Grimwood A. Antiretroviral treatment outcomes amongst older adults in a large multicentre cohort in South Africa. *PloS One* 2014; **9**: e100273.

39 Vazquez F. A systematic review and meta-analysis of the effect of HIV status on the incidence of tuberculosis disease among individuals with latent Mycobacterium tuberculosis infection. 2019.

40 Ackah AN, Coulibaly D, Digbeu H, *et al.* Response to treatment, mortality, and CD4 lymphocyte counts in HIV-infected persons with tuberculosis in Abidjan, Côte d'Ivoire. *Lancet Lond Engl* 1995; **345**: 607–10.

41 Mukadi YD, Maher D, Harries A. Tuberculosis case fatality rates in high HIV prevalence populations in sub-Saharan Africa. *AIDS* 2001; **15**: 143–52.

42 Rajagopalan S. Tuberculosis and aging: a global health problem. *Clin Infect Dis Off Publ Infect Dis Soc Am* 2001; **33**: 1034–9.

43 Andrianakis I, Vernon I, McCreesh N, *et al.* History matching of a complex epidemiological model of human immunodeficiency virus transmission by using variance emulation. *J R Stat Soc Ser C Appl Stat* 2017; **66**: 717–40.

44 Andrianakis I, Vernon IR, McCreesh N, *et al.* Bayesian History Matching of Complex Infectious Disease Models Using Emulation: A Tutorial and a Case Study on HIV in Uganda. *PLOS Comput Biol* 2015; **11**: e1003968.

45 Goldstein M. Bayes Linear Analysis for Complex Physical Systems Modeled by Computer Simulators. In: Dienstfrey AM, Boisvert RF, eds. Uncertainty Quantification in Scientific Computing. Berlin, Heidelberg: Springer Berlin Heidelberg, 2012: 78–94.

46 Williamson D, Goldstein M, Allison L, *et al.* History matching for exploring and reducing climate model parameter space using observations and a large perturbed physics ensemble. *Clim Dyn* 2013; **41**: 1703–29.

47 Iskauskas A. hmer: History Matching and Emulation Package. 2022 https://CRAN.R-project.org/package=hmer (accessed April 14, 2022).

48 Jabot F, Faure T, Dumoulin N, Albert C. EasyABC: Efficient Approximate Bayesian Computation Sampling Schemes. 2015 https://CRAN.R-project.org/package=EasyABC (accessed April 12, 2022).

49 Roberts GO, Rosenthal JS. Examples of Adaptive MCMC. *J Comput Graph Stat* 2009; **18**: 349–67.

50 New country classifications by income level: 2019-2020. https://blogs.worldbank.org/opendata/new-country-classifications-income-level-2019-2020 (accessed Sept 26, 2021).

51 World Health Organization. WHO | WHO End TB Strategy. 2015.

52 World Health Organization. WHO Preferred Product Characteristics for New Tuberculosis Vaccines. 2018 DOI:CC BY-NC-SA 3.0 IGO.

53 Country hub. https://www.gavi.org/programmes-impact/country-hub (accessed April 15, 2022).

54 Vaccination and Immunization Statistics. UNICEF DATA. https://data.unicef.org/topic/child-health/immunization/ (accessed March 7, 2022).

55 Harris RC, Sumner T, Knight GM, Zhang H, White RG. Potential impact of tuberculosis vaccines in China, South Africa, and India. *Sci Transl Med* 2020; **12**: eaax4607.

56 Tait DR, Hatherill M, Van Der Meeren O, *et al.* Final Analysis of a Trial of M72/AS01E Vaccine to Prevent Tuberculosis. *N Engl J Med* 2019; **381**: 2429–39.

57 Abubakar I, Pimpin L, Ariti C, *et al.* Systematic review and meta-analysis of the current evidence on the duration of protection by bacillus Calmette–Guérin vaccination against tuberculosis. *Health Technol Assess* 2013; **17**. DOI:10.3310/hta17370.

58 Lewnard JA, Grad YH. Vaccine waning and mumps re-emergence in the United States. *Sci Transl Med* 2018; **10**: eaao5945.

59 Blackwood JC, Cummings DAT, Broutin H, Iamsirithaworn S, Rohani P. Deciphering the impacts of vaccination and immunity on pertussis epidemiology in Thailand. *Proc Natl Acad Sci U S A* 2013; **110**: 9595–600.
